# Small-molecule functional rescue of PIEZO1 channel variants associated with generalised lymphatic dysplasia

**DOI:** 10.1101/2023.08.01.23292554

**Authors:** Melanie J Ludlow, Oleksandr V Povstyan, Deborah M Linley, Silvia Martin-Almedina, Charlotte Revill, Kevin Cuthbertson, Katie A Smith, Emily Fay, Elisavet Fotiou, Andrew Bush, Claire Hogg, Tobias Linden, Natalie B Tan, Susan M White, Esther Dempsey, Sahar Mansour, Gregory Parsonage, Antreas C Kalli, Richard Foster, Pia Ostergaard, David J Beech

**Affiliations:** Leeds Institute of Cardiovascular and Metabolic Medicine, School of Medicine, University of Leeds, Leeds, LS2 9JT, UK; Molecular and Clinical Sciences Institute, St George’s University of London, London, SW17 0RE, UK; School of Chemistry, University of Leeds, Leeds, LS2 9JT, UK; Paediatric Respiratory Medicine, NHLI, Imperial College London, London, UK; Department of Paediatric Respiratory Medicine, Royal Brompton Hospital, London SW3 6NP, and Paediatric Respiratory Medicine, Imperial College London, London, UK; Department of Neuropediatrics, University Children’s Hospital, Klinikum Oldenburg, Oldenburg, Germany; Victorian Clinical Genetics Services, Murdoch Children’s Research Institute, Parkville, Victoria, Australia; South West Thames Regional Centre for Genomics, St George’s NHS Foundation Trust, London, UK

**Keywords:** Calcium channel, Non-selective cation channel, Mechanical force, Pharmacology, Medicinal chemistry, Vascular biology, Endothelial cell, Lymphatics, Oedema, Non-immune fetal hydrops, Genetic disease.

## Abstract

Generalised Lymphatic Dysplasia (GLD) is characterised by widespread lymphoedema, with at least one of the following: fetal hydrops, intestinal or pulmonary lymphangiectasia, pleural effusions, pericardial effusions and ascites. Satisfactory medical therapies are lacking. A genetic association has been identified that prevents expression or surface trafficking of PIEZO1, a subunit of mechanically activated calcium-permeable channels. However, *PIEZO1* is a large and highly polymorphic gene and interpretation of variants identified in this gene can be challenging. *PIEZO1-*related GLD with non-immune fetal hydrops is autosomal recessive, however, heterozygous variants in *PIEZO1* (often gain-of-function) causing Dehydrated Hereditary Stomatocytosis (DHS) (a relative mild anaemia), may also present with perinatal non-immune hydrops (not caused by anaemia).

Here we sought to develop methods to confirm pathogenicity of missense variants of uncertain significance in *PIEZO1*, to gain deeper understanding and pharmacological solutions. Four novel GLD-associated missense variants in *PIEZO1* are identified that express and surface localise as full-length protein but with reduced or abolished mechanically activated channel function. Yoda1, a small-molecule agonist, functionally rescues the channels and their physiological regulation by mechanical force and hypo-osmolality. The GLD-associated variants mediate intracellular calcium release as well as calcium entry, suggesting two pools of channels and opportunity for increased rescue through access to the intracellular pool. New Yoda1 analogues are also identified that improve rescue.

The functional assays have assisted the interpretation of the variants of uncertain significance as the data suggest loss of PIEZO1 force sensing as a cause of the GLD observed in the patients. The potential to pharmacologically overcome the loss of force sensing was demonstrated and supports the concept of stimulation of PIEZO1 with an agonist to address wide-ranging problems of lymphatic insufficiency.

**GRAPHICAL ABSTRACT:** 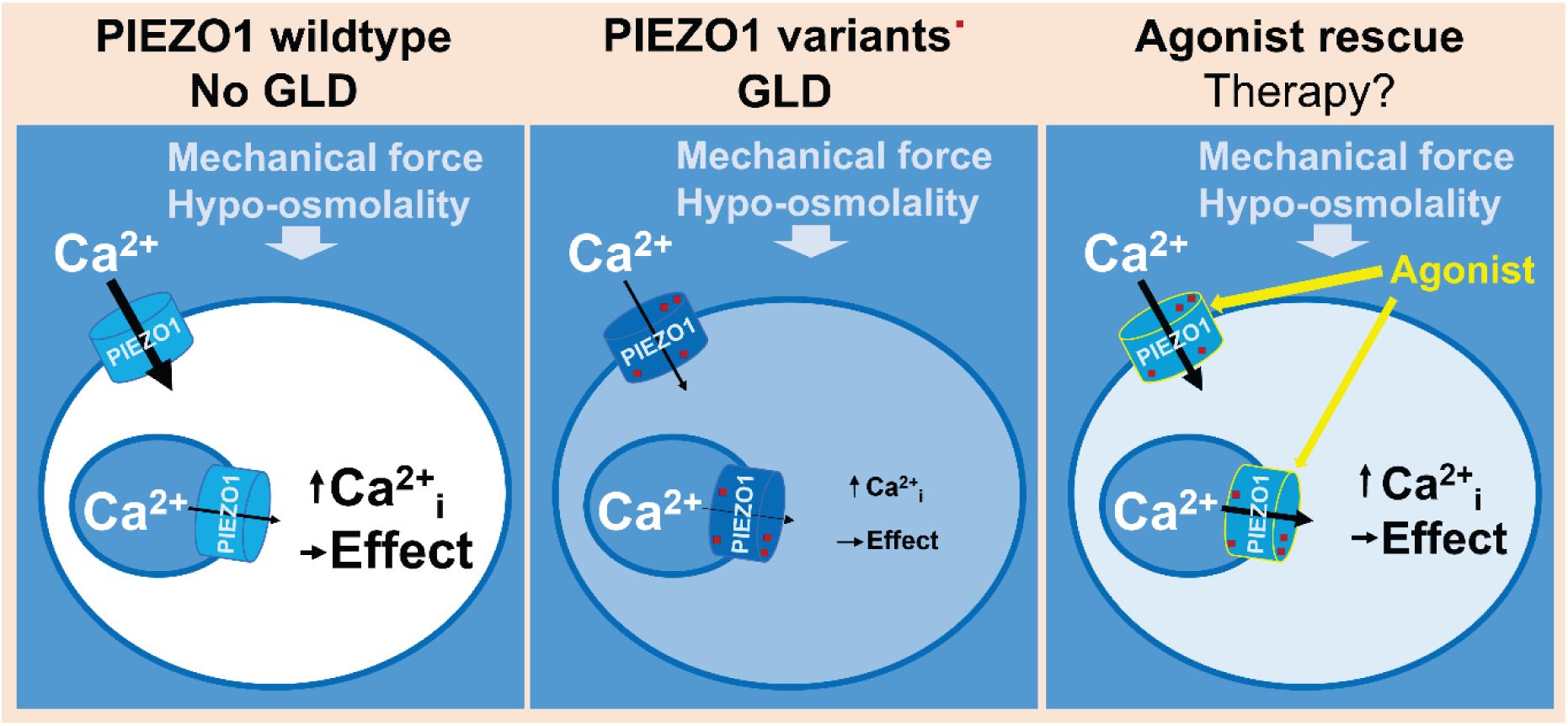

**HIGHLIGHTS:**

1. Previously unrecognised variants in *PIEZO1* that associate with GLD are identified and characterised and pathogenicity confirmed
2. The variants encode single amino acid changes that inhibit PIEZO1 channel activation by physiological mechanical forces
3. A small-molecule agonist rescues the channels and their physiological regulation
4. Variants are partly intracellular, suggesting an opportunity for improved rescue through the use of intracellular-acting agonists
5. New agonists are identified that improve rescue, suggesting routes to medical therapies for GLD and potentially other disorders of lymphatic insufficiency

## INTRODUCTION

The lymphatic system is required for fluid homeostasis, dietary fat absorption and immune function ^1–3^. It is a uni-directional system which begins as blind-ended lymphatic capillaries that converge into peristaltic collecting vessels, passing contents through lymph nodes to the thoracic duct and draining into the subclavian vein ^2, 4^. Lymphoedema results from imbalances between lymph formation and its absorption into lymphatic vessels, leading to excess interstitial fluid ^3^. Lymphoedema is a common condition that affects at least 120 million people globally and for which therapeutic options are limited ^4^. Most frequent is secondary lymphoedema (often caused by tissue damage e.g. infection, surgery, cancer) but primary disease (often caused by heritable defects) is a significant problem and one that has led to important insight into the molecular mechanisms of the human lymphatic system.

Generalised Lymphatic Dysplasia (GLD) is a form of primary lymphoedema estimated to affect about 1 in 6000 people ^5^. It presents with oedema of multiple segments (limbs, face, genitalia) and includes internal lymphatic problems such as pulmonary or intestinal lymphangiectasia, pleural effusions, chylothoraces, pericardial effusions, ascites and non-immune fetal hydrops (NIFH) in the second trimester ^5^. Patients with this condition frequently experience recurrent cellulitis particularly facial cellulitis which can be life-threatening, and there can be serious psychological consequences such as depression ^5, 6^. There are currently no cures or medical therapies; instead, symptoms are managed through diet and techniques that minimize fluid build-up and stimulate fluid flow, such as the wearing of compression garments, massage, skin care, and exercise ^5^.

Pathogenic genetic variants in at least 16 genes have been associated with GLD ^3^. One of the genes encodes PIEZO1 ^7, 8^, which is a large membrane protein of 38 membrane-spanning segments that assembles as a trimer to form a calcium ion (Ca^2+^)-permeable non-selective cationic channel ^9–12^. A striking feature of the channel is its exquisite sensitivity to activation by mechanical forces, leading to the hypothesis that its primary biological purpose is the detection of such forces and the coupling of them to cell function ^9, 13–16^. Although *PIEZO1* is widely expressed, murine gene disruption studies suggest particular importance in endothelial biology where it has a role in detecting shear stress, a force arising from the flow of blood or lymph abutting the endothelial surface ^17–24^. Essential roles for PIEZO1 in lymphatic development, maintenance and regeneration have been previously described ^21, 25, 26^. Studies on GLD patients suggest that the lymphatic endothelium is especially vulnerable to disruption of PIEZO1. GLD-associated *PIEZO1* variants have been identified and characterised that act by disrupting protein expression or localisation of PIEZO1 to the surface membrane ^7, 27^.

NIFH carries a high mortality rate. It is a non-specific, heterogeneous presentation, thus understanding the underlying cause can improve prognostication for the pregnancy. Fetal hydrops related to PIEZO1 may result in the baby’s demise but if the baby survives the neonatal period and the hydrops resolved, it can be associated with a reasonably good outlook. As *PIEZO1* is a large, highly polymorphic gene, variants of uncertain significance (VUS) are frequently identified. Interpretation of these variants can be challenging particularly in a fetus presenting with just hydrops. Developing a robust functional assay will assist in the interpretation of these VUS and improve patient management. Therefore, tools for interpretation of VUS are extremely important if identified during pregnancy.

Here we expand the depth of information about PIEZO1 in GLD. We identify novel, missense, variants in the *PIEZO1* gene. We characterise their effects, provide new mechanistic perspectives and suggest possible routes to a medical therapy.

## RESULTS

### Clinical cases with Generalised Lymphatic Dysplasia (GLD)

Since the report of 6 families with *PIEZO1* variants associated with NIFH and GLD ^7^, several additional probands with an overlapping phenotype have come to our attention. For this study, 3 probands and their families with missense variants were selected for further characterisation of the variants, initially to assist in evaluation of pathogenicity.

The clinical features of affected individuals are summarised in Table 1 and described in the SI. Pedigrees are presented in SI Figure S1. The proband (II.1) in family GLD07 had a typical presentation with antenatal NIFH, polyhydramnios and congenital oedema of the lower limbs, face and intermittent swelling of the scrotum with chylothoraces and some ascites (SI Figure S2).

**Table 1:**
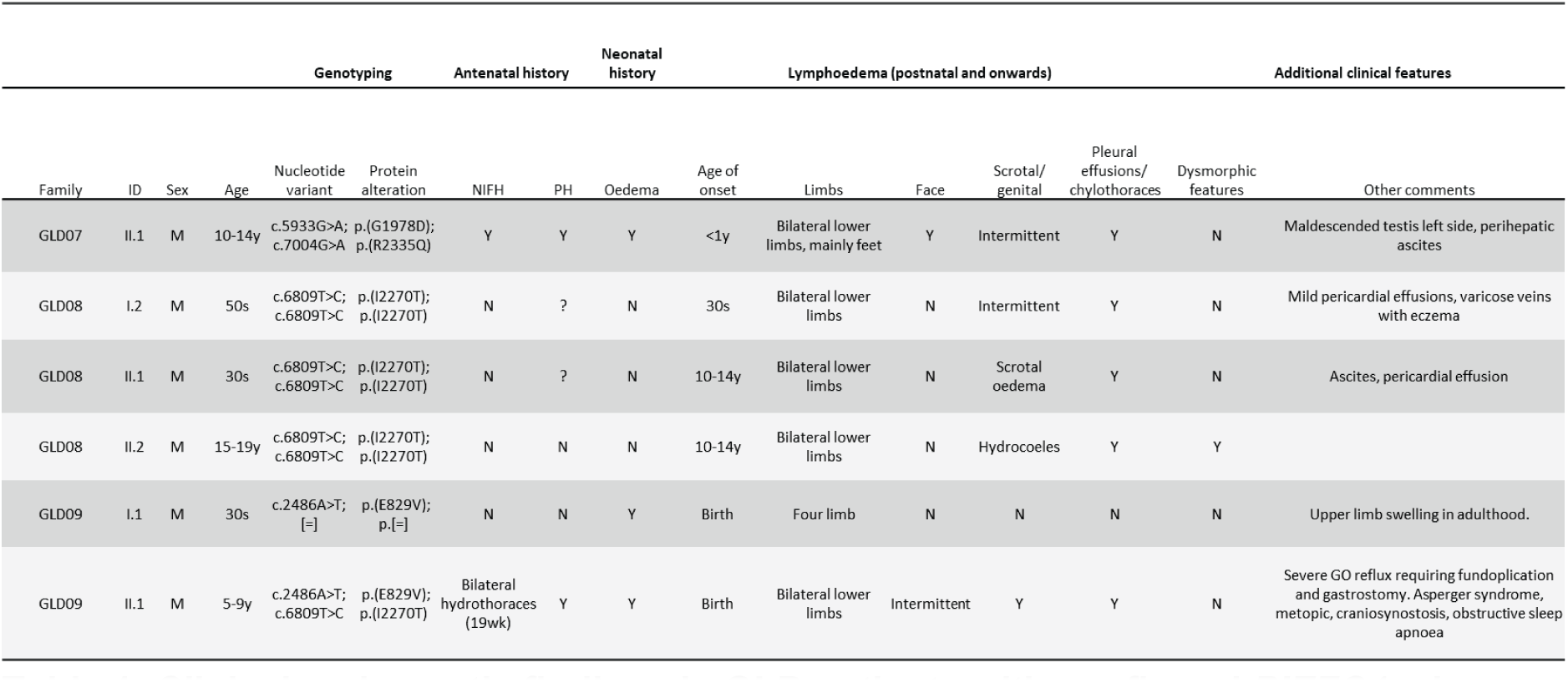
Clinical and genetic findings in GLD patients with confirmed *PIEZO1* missense variants. Family IDs are not known to anyone outside the research group and follow from the IDs used by us previously ^7^. Age at last clinical visit and age of onset are given as an age range. GO, gastroesophageal; M, male; N, no; NIFH, non-immune fetal hydrops; PH, polyhydramnios; wk, week; y, year; Y, yes; ?, not available.

A father (I.2) and two sons (II.1 and II.2) from family GLD08 presented with a pseudo-dominant mode of inheritance due to a high level of consanguinity. An affected aunt had died of chylothoraces. I.2 had bilateral chylothoraces and chylopericardium with onset in his 30s. He developed bilateral lower limb lymphoedema in his 40s. II.1 also had bilateral chylothoraces and chylopericardium in young adulthood (SI Figure S2). He developed bilateral lower limb lymphoedema with scrotal oedema some years later in his 30s. II.2 developed bilateral chylothoraces aged 10-14 years with mild lower limb lymphoedema and bilateral hydrocoeles.

The proband (II.1) from family GLD09 presented with bilateral hydrothoraces at 19 weeks gestation and polyhydramnios (SI Figure S2). He had congenital oedema of both lower limbs and the penile shaft and foreskin with intermittent scrotal oedema. His problems also included autistic spectrum disorder, metopic craniosynostosis and severe gastro-oesophageal reflux. His father (I.1) had congenital oedema of the lower limbs and developed swelling of both hands in his 20s but no systemic involvement.

### Variant identification, interpretation and classification

All coding exons of *PIEZO1* were screened in the 3 probands using Sanger sequencing, identifying 4 novel or rare missense variants in *PIEZO1* (SI Table S1). The variants encode the amino acid changes G1978D and R2335Q *in trans* in the proband of family GLD07, a homozygous I2270T variant in two generations of the second family GLD08 (pseudo-dominant inheritance), and E829V and I2270T *in trans* in the proband of the third family GLD09 (Table 1, SI Figure S1; single-letter amino acid codes are used).

Wild-type (WT) amino acids at these positions are conserved across diverse species and in the only related channel protein, PIEZO2, except for isoleucine’s replacement by leucine in the case of I2270 (SI Figure S3). The variants are predicted to be pathogenic by most *in silico* tools (SI Table S1). One variant, G1978D, is not observed in gnomAD, the other 3 variants are extremely rare (MAF = 0.0000197 – 0.00000654) and none is observed in homozygous carriers. However, classification of the variants using the ACMG guidelines ^28^ places all 4 in class 3, which indicates a VUS because there is not enough evidence for scoring it as pathogenic (SI Table S1). Therefore, we used laboratory methods to investigate the variants.

### Inhibitory effects of variants on channel function

WT human PIEZO1 (hPIEZO1) and variant forms of it were transiently overexpressed in a modified HEK 293 cell line. Intracellular Ca^2+^ was quantified by measuring fluorescence (F) from a ratiometric Ca^2+^ indicator fura-2 ^29^, since PIEZO1 channels are Ca^2+^ permeable and usually cause elevation in intracellular [Ca^2+^] when activated ^13^. Yoda1, a synthetic PIEZO1 agonist, was applied at a concentration of 1 μM to induce about 30% of its maximum effect in WT hPIEZO1 ^30^. WT channels respond with a peak ratio change (ΔF_ratio_) of about 0.2 compared with <0.05 in empty vector-transfected control cells (Figure 1a). hPIEZO1 mutants that mimic the GLD-associated variants (G1978D, R2335Q, E829V and I2270T) lead to signals above the background of control cells but 65- 90% less than those of WT (Figure 1a, b). Similar effects occur when the variants are recapitulated in mouse PIEZO1 (mPIEZO1) (G1994D, R2351Q, E824V and I2286T) (Figure 1b, c). Patch-clamp technique provided an orthogonal assay, enabling recording of electric current through the channels and determination of responses to membrane stretch (instead of Yoda1) induced by pressure applied to the patch pipette. The membrane patches contain multiple channels, so macroscopic (i.e., multiple unitary) currents are seen (Figure 1d). Increasing current is shown as downward deflection (i.e., negative values) based on convention for ionic current arising from inward flux of cations. WT channel currents were compared with currents through channels containing I2270T, which occurs in 2 of the families (SI Figure S1). WT channel shows the expected ^13^ rapid activation (i.e., initial fast rise in inward current) followed by inactivation (i.e., progressive loss of inward current despite sustained pressure) whereas I2270T shows no current (Figure 1d). Overall, the data suggest inhibitory effects of the GLD-associated variants; i.e., loss-of-function phenotypes.

**Figure 1:**
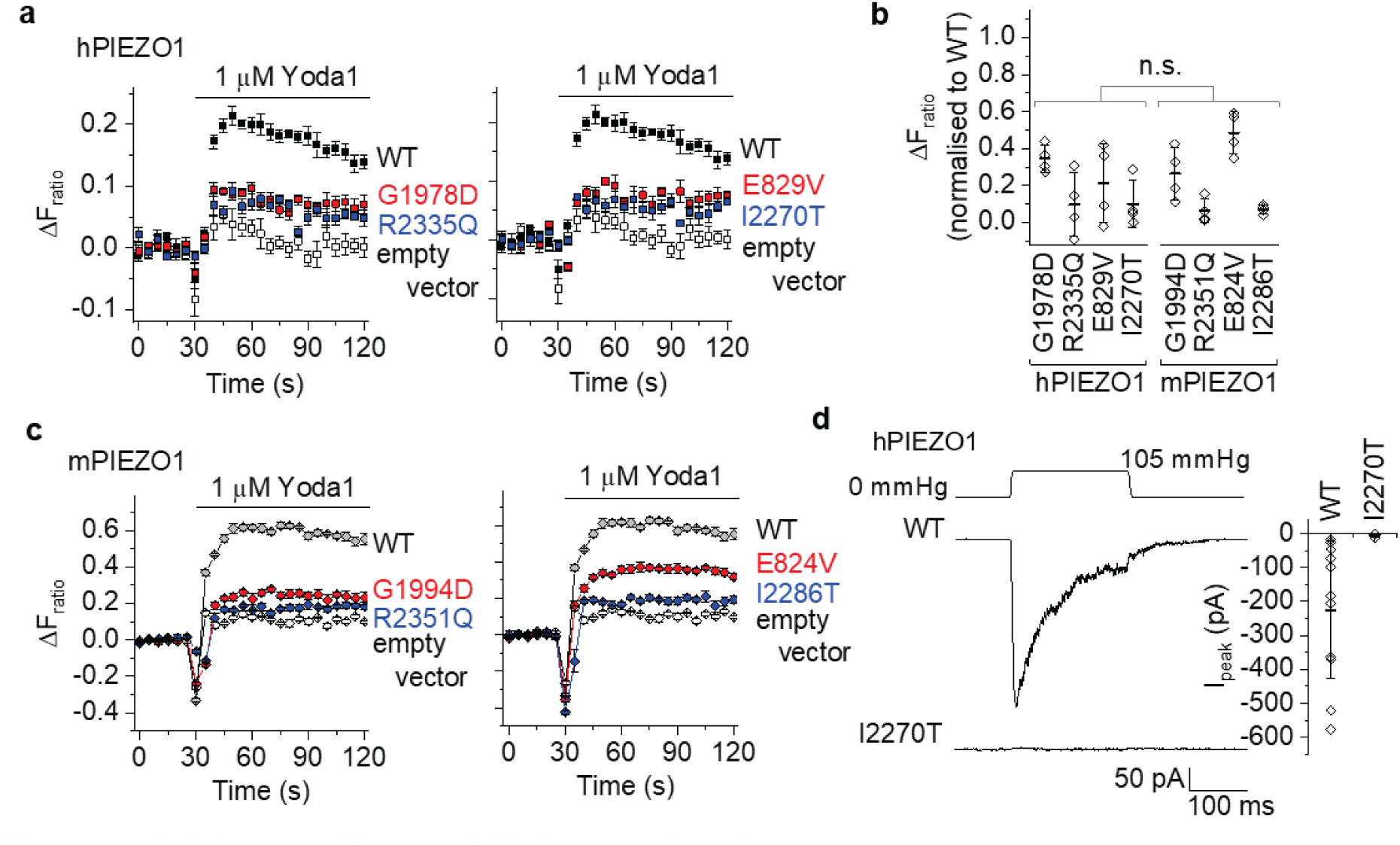
Inhibitory effects of GLD-associated variants. (a) Change (Δ) in intracellular Ca^2+^ concentration measured by fura-2 in T-REx-293 cells transiently transfected with hPIEZO1 wild-type (WT) and variant hPIEZO1 or empty vector control. Cells were stimulated with 1 μM Yoda1. Increase in Ca^2+^ is indicated by increase in the fura-2 fluorescence (F) ratio (ΔF_ratio_). Data are from single representative 96-well plate experiments (mean ± s.e.m., N = 4-5 wells each). (b) Summary data for experiments of the type shown in (**a**, **c**) for hPIEZO1 and mPIEZO1 measured between 30 and 60 seconds (s) after Yoda1 application (n = 4 independent experiments each). Data are mean ± s.d. normalised to the respective WT channel data. Data for analogous variants between the two species are compared by one-way Anova with Bonferroni post-hoc test (n.s., no significant difference). (c) As for (**a**) but mPIEZO1 and variant mPIEZO1 (data for N = 5 wells each). (d) Left: Example ionic currents in outside-out patch recordings from T-REx-293 cells expressing WT or I2270T hPIEZO1 exposed to the pressure pulse shown schematically at the top. Right: Summary data for the types of experiment shown on the left. Amplitude of the peak current is represented as mean ± s.d. and each independent data point is superimposed (WT n = 13, I2270T n = 4).

### Suppressed but not abolished expression

Potential explanations for loss-of-function are failed expression or surface localisation ^7, 27^. We investigated changes in protein expression and localization for the 4 novel variants by expressing hemagglutinin (HA)-tagged WT and variant hPIEZO1 or mPIEZO1 in the HEK 293 cell line. We performed biochemical analysis including protein surface biotinylation, using anti-HA antibody to specifically recognise exogenously-expressed channels. Importantly, the variants express and localise to surface membrane (Figure 2a-c). The variants are detected in lower amounts than WT but for 3 of them the reductions are relatively small (Figure 2a-c) compared with the large effects on their function (Figure 1a-d). The E829V and E824V variants (hPIEZO1 and mPIEZO1 respectively) are an exception, showing strongly reducing protein expression relative to WT and reducing expression of WT when it is coexpressed, suggesting dominant negative effect of the E829V variant (Figure 2a-d). Overexpression of WT did not rescue this variant (Figure 2d). The data suggest that all variants express as intact proteins at the surface membrane. There are reductions in protein amounts relative to WT but not sufficient to explain the strong loss-of-function seen with variants such as I2270T (e.g., Figure 2a compared with Figure 1d).

**Figure 2:**
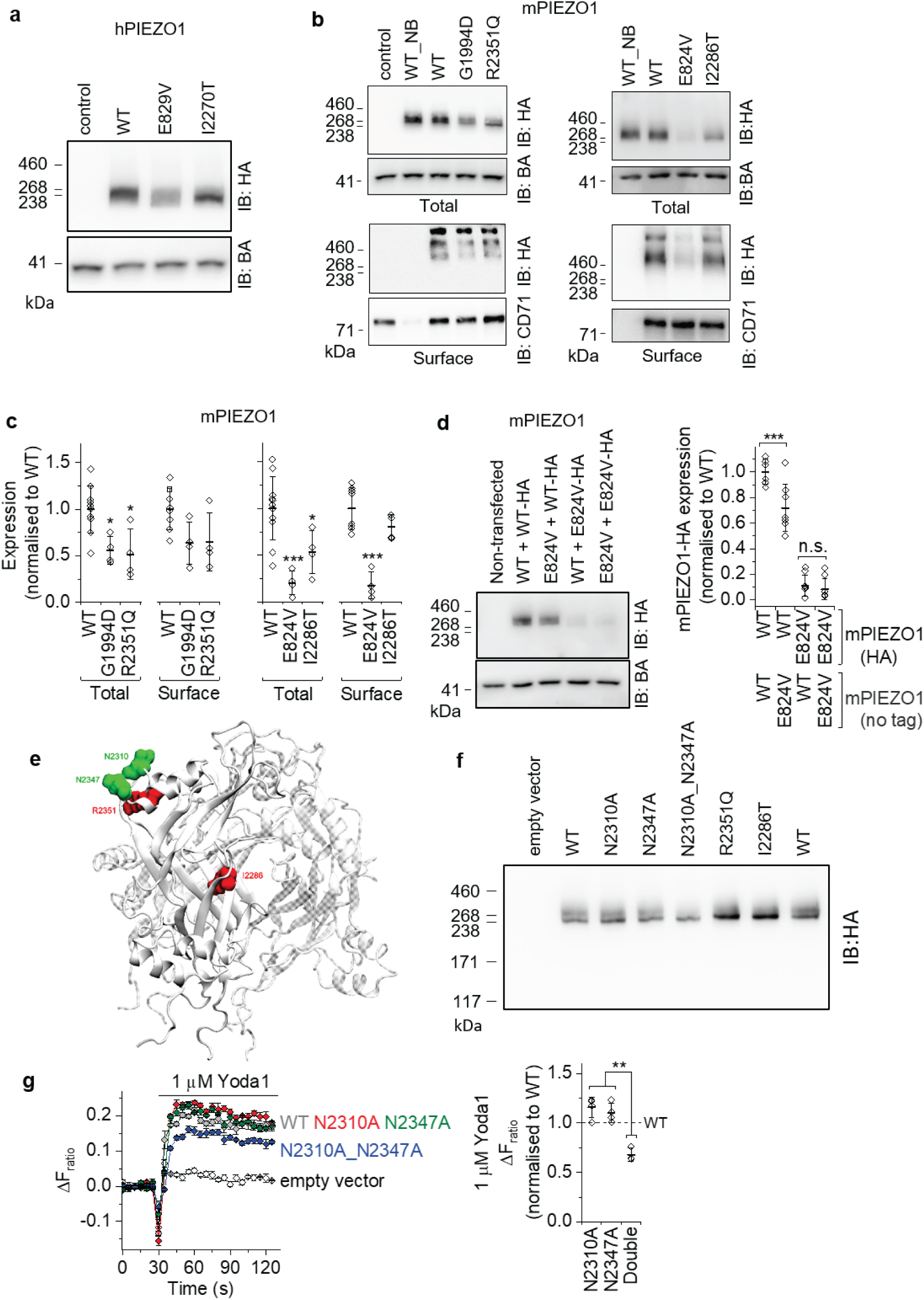
Suppressed protein expression and glycosylation. (a) Representative western blot for empty vector control and total wild-type (WT) hPIEZO1 and HA-tagged variant (E829V and I2270T) hPIEZO1_HA constructs transiently expressed in T-REx-293 cells and immuno-blotted (IB) with anti-HA antibody (upper panel) and anti-beta-actin (BA) antibody (lower panel, loading control). Mean ± s.d. 0.59 ± 0.17 E829V relative to 1.0 WT (n = 3). (**b**, **c**) Representative western blots (**b**) and summarised mean ± s.d. data from such blots (**c**) (n = 4-11) for total and surface (extracted following cell surface protein biotinylation) WT and variant mPIEZO1_HA constructs transiently expressed in T-REx-293 cells (control, empty vector; NB, non-biotinylated). Beta-actin (BA) and CD71 were the protein loading and surface protein controls respectively. (d) Representative western blot (left) and summarised density quantification data (right) (n = 6) following co-transfection of the E824V variant with WT mPIEZO1. Each pairing includes one HA-tagged and one untagged construct, allowing the impact of the untagged on the tagged protein to be assessed. Data are shown as mean ± s.d. normalised to WT channel with statistical significance determined by one-way Anova and Bonferroni post-hoc test (\*\*\**P*<0.001, n.s. not significantly different). (e) Molecular model of mPIEZO1 C-terminal extracellular domain (CED) highlighting the locations of predicted N-linked glycosylation sites in green and CED-localized GLD-associated variants in red. The CED is shown in cartoon representation. The channel comprises 3 PIEZO1 proteins, which are shown for the CED as 3 chains (A, B and C). Chains B and C are transparent. The GLD-associated variants are shown on A only. Residues are shown in surface representation. (f) Representative western blot for total WT and variant mPIEZO1_HA constructs (as indicated in the figure panel) transiently expressed in T-REx-293 cells. Immuno-blotted (IB) with anti- HA antibody. Note the twin band in the WT lane but not the double mutant lane (N2310A_N2347A). (g) Left: Change (Δ) in intracellular Ca^2+^ concentration in HEK 293 cells transiently transfected with mPIEZO1 WT and mutant (N2310A, N2347A or both of these mutations N2310A_N2347A or ‘Double’) constructs or empty vector control. Cells were stimulated with 1 μM Yoda1. Data are from a single representative 96-well plate experiment (mean ± s.e.m., N = 5 wells each). Right: Summary for experiments of the type shown on the left, measured between 30 and 60 s after Yoda1 application (n = 3-4). Data are mean ± s.d. normalised to WT channel (dotted line), with statistical significance determined by one-way Anova and Bonferroni post-hoc test (\*\**P*<0.01).

### Diverse structural loci

To generate hypotheses for how channel function is lost and might be regained, we generated molecular models of hPIEZO1 and mPIEZO1 channels (SI Figure S4) based on experimentally-derived high resolution structural data for mPIEZO1 ^11, 31–33^. I2270T/I2286T (hPIEZO1/mPIEZO1) and R2335Q/R2351Q are at opposite corners of the C-terminal extracellular domain (CED), G1978D/G1994D is in the central membrane-spanning region (but outside the pore-lining helices and inactivation gate) and E829V/E824V is distal from the central core, far out in the blade structures that determine mechanical sensitivity and are linked to the ion pore region via the beam structure (SI Figure S4). The putative interaction site for Yoda1 ^34^ does not overlap with the affected amino acids (SI Figure S4). Comparisons of hPIEZO1 and mPIEZO1 suggest strong conservation of structure, including at the loci of the 4 variant residues (SI Figure S4), which is consistent with the similar functional results for hPIEZO1 and mPIEZO1 (Figure 1b). The data suggest diverse structural locations of the affected amino acids, previously unrecognised functional domains of PIEZO1 and cross-species conservation of the affected sites.

### Suppressed glycosylation

To investigate variant channels in detail, we focused on mPIEZO1 because it is the basis of PIEZO1 structural data and generates larger and more robust functional signals than hPIEZO1, facilitating quantitative analysis (e.g., Figure 1c compared with Figure 1a). As shown in SI Figure S4, I2286T and R2351Q are in the CED. This is the site of functionally important N-linked glycosylation ^35^. Both residues are close to the glycosylated asparagine (N) residues (Figure 2e, SI Figure S5). Consistent with mPIEZO1 being glycosylated at these residues, it migrates at two molecular masses (Figure 2f). Mutation of the asparagine residues to alanine (N2310A and N2347A) abolishes the upper mass band (Figure 2f). I2286T and R2351Q reduce the density of the upper relative to the lower mass (Figure 2f), suggesting impaired glycosylation. Similarly, I2270T reduces the intensity of the upper mass in hPIEZO1 (Figure 2a). N2310A_N2347A double mutation reduces Yoda1- evoked Ca^2+^ signals by about 30%, consistent with there being importance of glycosylation in channel function (Figure 2g). The data suggest loss of glycosylation as a contributor to the loss-of-function seen in I2270T and R2335Q variants, but that it is not a large enough functional effect to explain their major impact.

### Suppressed or abolished mechanical activation

The I2270T variant shows loss of mechanical sensitivity (Figure 1d) and the location of 2 of the variant residues is the blade structure (SI Figure S4), which is a region that determines mechanical sensitivity of the channels ^16^. We therefore hypothesised that some variants affect the mechanical sensitivity of the channels. We sought to test this idea experimentally. Low endogenous expression of hPIEZO1 sometimes occurs in HEK 293 cell lines ^36^, but no pressure-evoked currents are detected in cells transfected with vector alone (Figure 3a, SI Figure S6). By contrast, in cells transfected with WT mPIEZO1 there are inward currents that increase and gradually saturate as the maximum pressure step is approached (Figure 3b-d, SI Figure S6). WT currents exhibit three kinetic types: fast activation and inactivation; fast activation and non-inactivation; and slow activation and non-inactivation (Figure 3b, c). Native PIEZO1 channels similarly show variable inactivation ^33, 37, 38^, possibly due to variable lipid regulation ^33^. Pressure required for 50% activation (P_50_) of peak WT current is 47.8 mmHg (Figure 3c, d). This is similar for currents of the three types or current measured at the end of the 200-ms pressure pulse (SI Figure S6). Like I2270T in hPIEZO1 (Figure 1d), I2286T in mPIEZO1 prevents mechanical activation of current (Figure 3e, f). R2351Q similarly prevents mechanical activation (Figure 3e, f). G1994D and E824V do not prevent current but shift the pressure curve to the right and reduce the activation rate and inactivation (Figure 3e, g, h). G1994D currents do not reach a maximum (Figure 3h). E824V channels activate at a similar or lower threshold compared with WT except there is a biphasic pressure-response relationship, with the second phase requiring high pressures (Fig 3h). P_50_ for the first phase is 38.9 mmHg but the second P_50_ could not be determined because saturating responses did not occur (Figure 3h). Pressure pulses larger than 105 mmHg often damaged patches, making them leaky to non-specific current, so no data are shown past this pressure. The data suggest that the variants inhibit the ability of the channels to respond to mechanical force.

**Figure 3:**
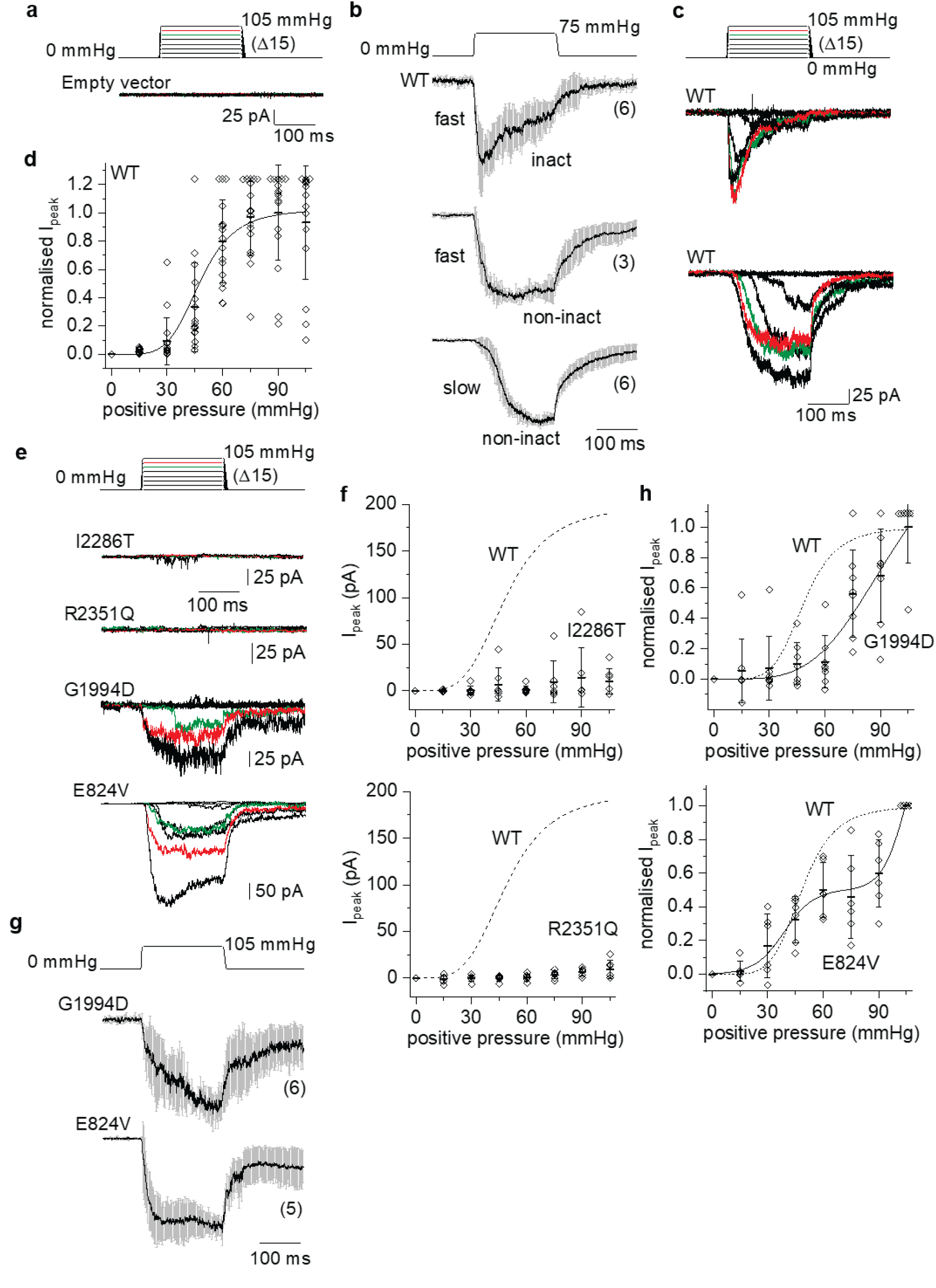
Suppressed or abolished mechanical activation. Data for outside-out patch recordings from T-REx-293 cells transfected with empty vector or mPIEZO1 constructs. The voltage across the patches during recordings was -80 mV. (a) Upper: Schematic of the pressure pulse protocol in which a 200-ms pressure pulse was applied to 15 mmHg and then, every 12 s, incremented (Δ) in steps of 15 mmHg up to a maximum of 105 mmHg. Lower: Example ionic currents from a patch excised from an empty vector-transfected cell. No currents were evoked by the pressure pulses (current traces superimpose). (b) As for (**a**) but for a single pressure step and wild-type (WT) mPIEZO1-transfected cells. Data shown mean ± s.e.m. for the 3 categories of ionic current kinetics observed, as described in the text (n = 6, 3 and 6, as indicated in parentheses). (c) As in (**a**) but examples of the 2 most common current kinetics for WT channels (fast and inactivating, and slow and non-inactivating). Currents for 75 and 90 mmHg are in green and red respectively. (d) Quantification of peak current amplitude for all WT experiments designed as in (**c**), plotted across the pressure range, normalised to the current at 90 mmHg in each experiment and shown as mean ± s.d. with individual data points for each experiment superimposed (n = 15- 17). The smooth curve is a fitted Boltzmann function with mid-point (P_50_) of 47.8 mmHg. (e) As in (**c**) but for the variant channels indicated and showing only one example per variant. (f) As in (**d**) but data for the I2286T (n = 7) and R2351Q (n = 6) variant channels. The Boltzmann function is not fitted to these data because there is little or no channel activity. The WT Boltzmann curve from (**d**) is superimposed for comparison as a dashed line. (g) As for (**b**) but G1994D and E824V channels and with a pressure pulse to 105 mmHg. A 6^th^ recording from E824V channels (not displayed) yielded a fast activating and inactivating current. (h) As in (**d**) but data for the G1994D (n = 7-8) and E824V (n = 6) channels. A single Boltzmann function is fitted to the G1994D data but no P_50_ is indicated because saturation is not achieved. A double Boltzmann function is fitted to the E824V data. The P_50_ value for the first phase is 38.9 mmHg. No P_50_ is indicated for the second phase because saturation is not achieved. The WT Boltzmann curve from (**d**) is superimposed for comparison as a dashed line.

### Pharmacological rescue

Although R2351Q and I2286T channels do not activate in response to mechanical force (Figure 3e, f), they show small responses to 1 μM Yoda1 (Figure 1b, c). Moreover, the structural models place all variant residues away from the putative Yoda1 interaction site ^34, 39^ (SI Figure S4), suggesting that Yoda1 activity might be retained. We investigated this further by constructing Yoda1 concentration response curves in Ca^2+^ assays for empty vector- and mPIEZO1-transfected HEK 293 cells (Figure 4a-c). Empty vector-transfected cells show small or no response to Yoda1 (Figure 4a) whereas overexpressed WT channels show robust responses; a sigmoidal concentration-response curve with an approximate threshold for activation at about 0.03 μM, saturating maximum response between 1 and 10 μM and concentration for 50% activation (EC_50_) at 0.24 μM (Figure 4b, c). All of the variant channels show robust Ca^2+^ responses (Figure 4c-g). Concentrations of Yoda1 above 1 μM greatly amplify their responses but thresholds for activation are to the right of WT (above 0.1 μM) and saturating curves and EC_50_s are not obtained (Figure 4c). Concentrations of Yoda1 above 10 μM are not reliably investigated because Yoda1 has poor aqueous solubility at such concentrations ^40^. Similar results occur in independent stable cell lines for WT and variant channels (SI Figure S7). Coexpressed G1994D and R2351Q channels aimed at mimicking the compound heterozygous state of patient GLD07 II.1 (SI Figure S1) are also activated by Yoda1 (Figure 4h, i). The data suggest potential for pharmacological rescue of variant channels.

**Figure 4:**
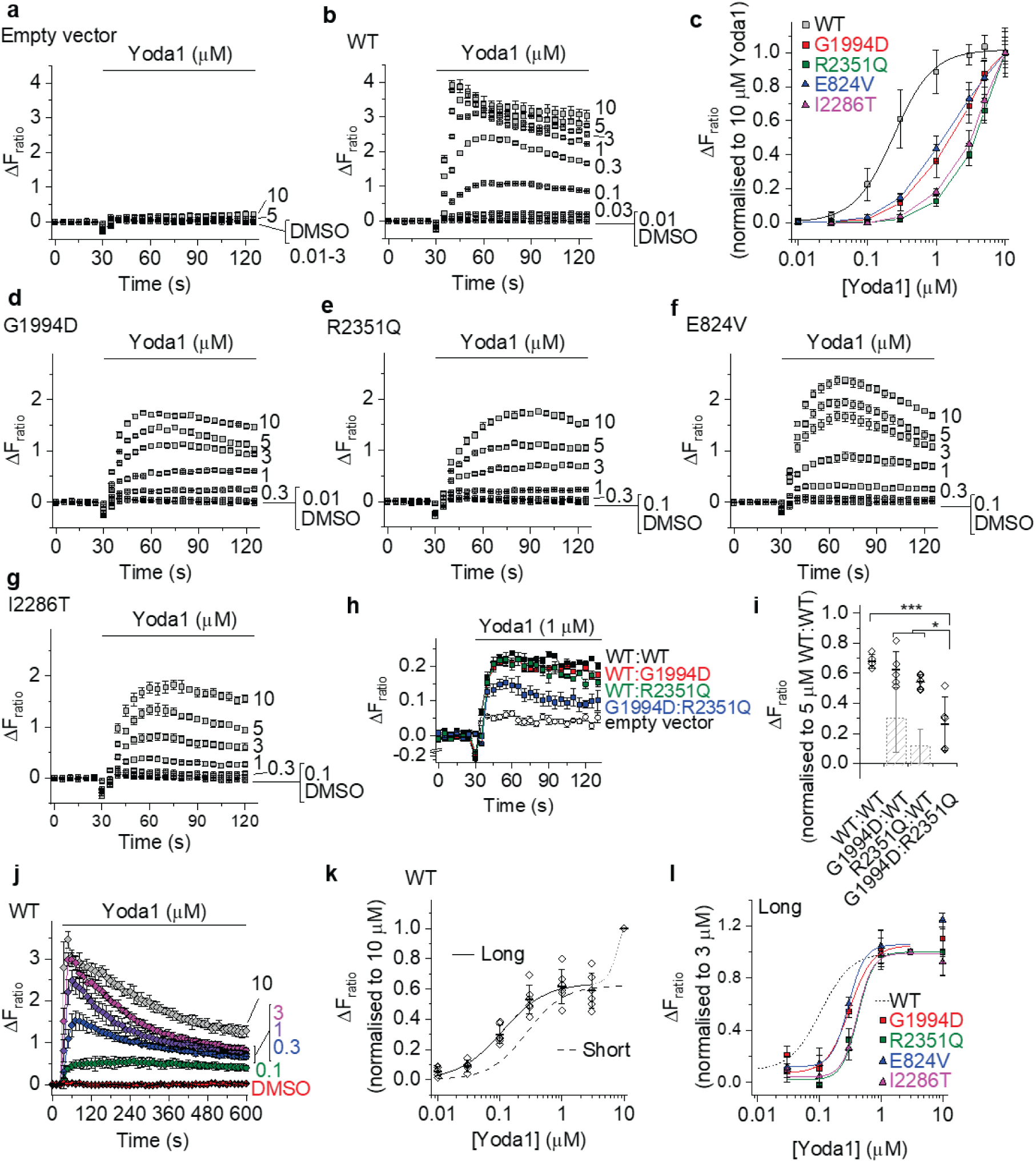
Rescue of Ca^2+^ signalling by Yoda1. Ca^2+^ measurement data from T-REx-293 cells transfected with empty vector or wild-type (WT) or variant mPIEZO1 constructs. Data are displayed as the change (Δ) in intracellular Ca^2+^ as indicated by change in the fura-2 fluorescence (F) ratio (ΔF_ratio_), with increase in ratio indicating increase in Ca^2+^ concentration in the cytosol. (**a**, **b**, **d**-**h**, **j**) Data are each from a single representative 96-well plate experiment exposed to the indicated Yoda1 concentrations in μM or vehicle (DMSO) for cells transfected with: (**a**) empty vector; (**b**) WT mPIEZO1; (**d**) G1994D mPIEZO1; (**e**) R2351Q mPIEZO1; (**f**) E824V mPIEZO1; (**g**) I2286T mPIEZO1; (**h**) Cells transfected with the combinations of constructs or empty vector as indicated in the figure panel; (**j**) As for (**b**) but with longer exposure to Yoda1 at the indicated concentrations in μM. Mean ± s.e.m. and N = 4-5 wells each. (**c**) Summary concentration-response data for experiments of the type shown in (**b**, **d**, **e**, **f**, **g**) (n = 3-6) with a Hill equation fitted to the WT data (EC_50_ 0.24 μM) and data points for other constructs joined by straight lines. (**i**) Summary mean ± s.d. data normalised for experiments of the type shown in (**h**) with individual data points superimposed (n = 5). Data were generated using 1 μM Yoda1 (as in **h**) and normalised to responses of WT:WT to 5 μM Yoda1. \**P*<0.05 \*\*\**P*<0.001. The 2 hatched bars are for G1994D and R2351Q expressed with empty vector only (n = 4). (k) As for (**c**) but WT only and long exposure to Yoda1 (20-50 min). A double Hill equation is fitted, generating an EC_50_ for the first phase of 0.11 μM. The dashed curve for short exposure data is the Hill fit for the WT data in (**c**). (n = 5-6). (l) Yoda1 long exposure data as for (**k**) but for the 4 variant channels as indicated and with fitted Hill equations giving EC_50_s of 0.31 μM (G1994D), 0.42 μM (R2351Q), 0.29 μM (E824V) and 0.41 μM (I2286T). The first phase of the WT fitted curve from (**k**) is superimposed for side- by-side comparison. (n = 3-5).

### Improved rescue with long agonist exposure

Responses of variant channels are slow compared with WT channels (Figure 4d-g compared with Figure 4b) and so we recorded Ca^2+^ for longer, should short recordings underestimate variant channel responses. WT channel responses to high concentrations of Yoda1 peak rapidly and decay but there is still Ca^2+^ elevation at 10 min (Figure 4j). Responses to low concentrations of Yoda1 are mostly sustained (Figure 4j). We extended exposure to at least 20 min and constructed concentration-responses curves. The Yoda1 EC_50_ for WT channels is now 2.2-fold improved at 0.11 μM (Figure 4k compared with Figure 4c). A second phase of effect occurs at 10 μM, which is not seen in the short recordings (Figure 4k). Long exposure enables full concentration-responses curves for all variant channels, revealing EC_50_s closer to those of WT channels at 0.31 μM (G1994D), 0.42 μM (R2351Q), 0.29 μM (E824V) and 0.41 μM (I2286T) (Figure 4l). The data suggest that long exposure enhances Yoda1 potency and rescue.

### Involvement of intracellular PIEZO1 channels

Long exposure to Yoda1 could activate downstream mechanisms that amplify the Ca^2+^ signal, such as ATP release activating P2Y2 receptors that lead to Ca^2+^ release via inositol 1,4,5-triphosphate signalling ^23^ and arachidonic acid metabolism activating Ca^2+^ permeable TRPV4 channels ^41^. However, inhibiting ATP and TRPV4 responses has no effect on Yoda1 responses (SI Figure S8). Alternatively, there could be PIEZO1 in intracellular membranes mediating Ca^2+^ release ^42^ in addition to PIEZO1 in surface membrane mediating Ca^2+^ entry, each with different Yoda1 sensitivity due to different accessibility. Lowering of the extracellular Ca^2+^ concentration only partly inhibits the Yoda1 response (Figure 5a), consistent with Ca^2+^ release occurring as well as Ca^2+^ entry. In support of a Ca^2+^ release contribution, predepletion of Ca^2+^ stores by thapsigargin ^43^ also partly inhibits the Yoda1 response (Figure 5b). As expected because of limited Ca^2+^ storage capacity, extracellular Ca^2+^ becomes critical when Yoda1 exposure is long (Figure 5c). Yoda1 responses of G1994D channels are relatively little affected by thapsigargin but R2351Q, E824V and I2286T channel responses are strongly inhibited (Figure 5d-f). The data suggest a contribution of intracellular Ca^2+^ release mediated by PIEZO1 channels in the functional rescue by Yoda1.

**Figure 5:**
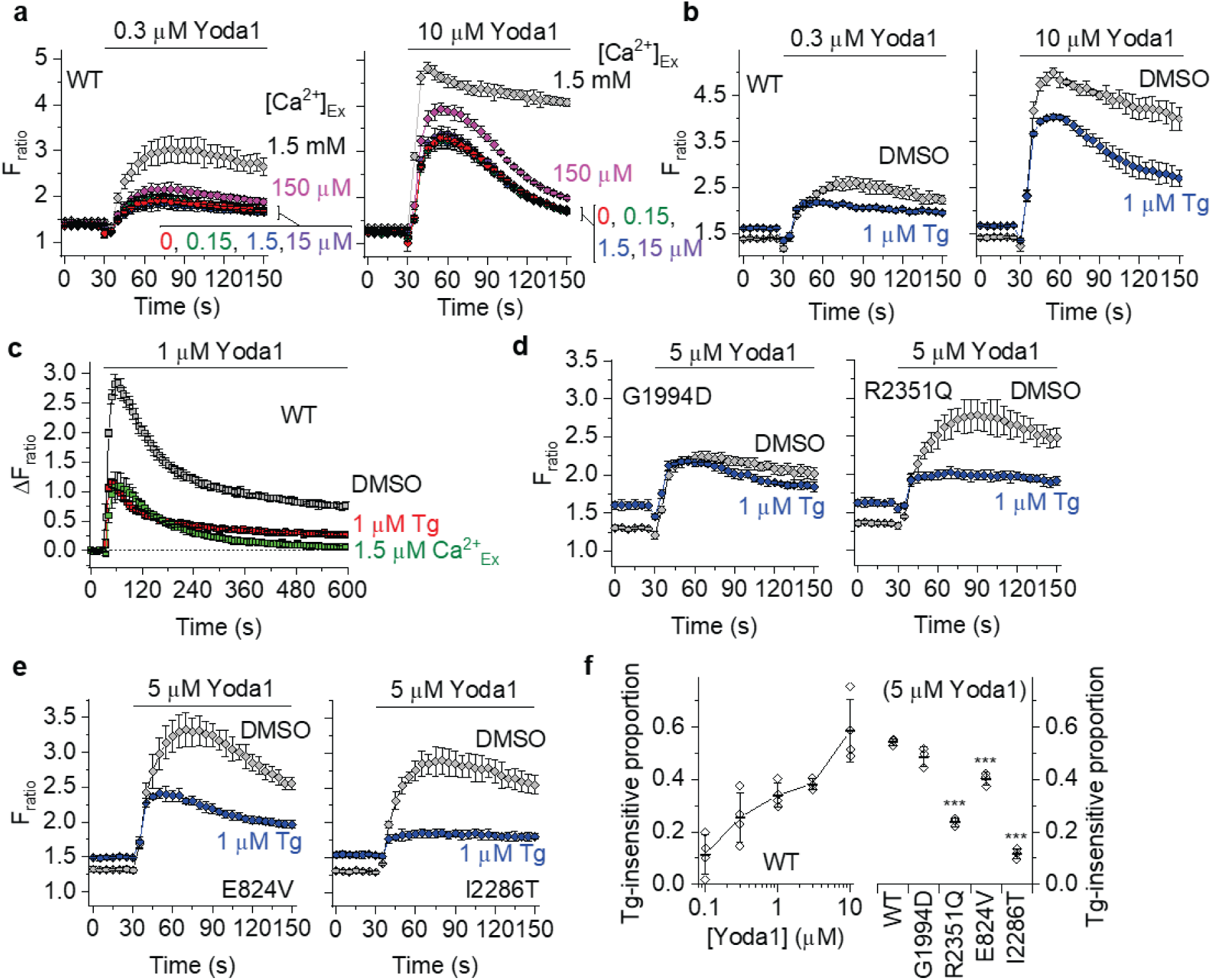
Contribution of intracellular channels to the Yoda1 response. Data for Ca^2+^ measurements from T-REx-293 cells transfected with wild-type (WT) or variant mPIEZO1 constructs. (**a**-**e**) Data are displayed as the intracellular Ca^2+^ concentration indicated by absolute or change (Δ) in fura-2 fluorescence (F) ratio (F_ratio_ or ΔF_ratio_), with increase in ratio indicating increase in Ca^2+^ concentration in the cytosol. Data are each from a single representative 96- well plate experiment with the indicated Yoda1 concentrations in μM or vehicle (DMSO) control (mean ± s.e.m., N = 4-5 wells each). (**a**-**c**) Data for WT channel. (**d**, **e**) Data for the indicated variant channels. (a) Responses to 0.3 μM (left) and 10 μM (right) Yoda1 in various concentrations of extracellular Ca^2+^ ([Ca^2+^]_Ex_) as indicated in the figure panels. (b) Responses to 0.3 μM (left) and 10 μM (right) Yoda1 in 1.5 mM [Ca^2+^]_Ex_ in the presence of 1 μM thapsigargin (Tg) or its vehicle control (DMSO). (c) Long-duration exposures to 1 μM Yoda1 in 1.5 mM [Ca^2+^]_Ex_ in the presence of 1 μM thapsigargin (Tg) or its vehicle control (DMSO) or in 1.5 μM [Ca^2+^]_Ex_ in the absence of Tg. (**d**, **e**) Responses to 5 μM Yoda1 in 1.5 mM [Ca^2+^]_Ex_ in the presence of 1 μM Tg or its vehicle control (DMSO) for the 4 variant channels as indicated in the figure panels. (**f**) Summary data for Tg experiments of the type exemplified in (**b**, **d**, **e**) (n = 3-4 each) showing the amplitudes of the Tg-insensitive responses as fractions of the total responses. Data are mean ± s.d. with individual data points superimposed. Statistical significance was determined by one-way Anova with Bonferroni post-hoc test. \*\*\**P*<0.001 compared with WT.

### Rescued mechanical force sensitivity

Yoda1 synergises with mechanical force in PIEZO1 activation ^44^ and this could be important *in vivo* because of the presence of endogenous forces such as lymphatic pressure. Pressure pulses were therefore again used to increase membrane tension and activate the WT and variant channels. Pressure-activated WT currents in outside-out patches are enhanced by Yoda1 (Figure 6a). G1994D channels were investigated next because they express robustly at the surface membrane (Figure 2b, c) and reduce without abolishing pressure responses (Figure 3h). Similar to WT, G1994D currents are enhanced by Yoda1 (Figure 6b). Yoda1 protected these channels from spontaneous run- down (Figure 6b). Basal activation by Yoda1 is minimal in the absence of the pressure pulse, consistent with Yoda1 synergising with force (Figure 6a, b). In the absence of Yoda1, G1994D channel currents do not reach a maximum even at 105 mmHg (Figure 3h) but Yoda1 increases the pressure sensitivity so that a full curve can be constructed, revealing a P_50_ of 64.6 mmHg (Figure 6c), which is an improvement, although still to the right of WT in the absence of Yoda1. R2351Q and I2386T variant channels are pressure insensitive without Yoda1 (Figure 3f) but respond to pressure in Yoda1’s presence (Figure 6d), showing greater fold-change response to Yoda1 than WT channels (SI Figure S9). E824V channels were not investigated in this assay because their pressure sensitivity is only modestly different from that of WT channels (Figure 3h). The data suggest that pharmacologically rescued variant channels become sensitive to mechanical force, thus with the potential to respond to physiological stimuli.

**Figure 6:**
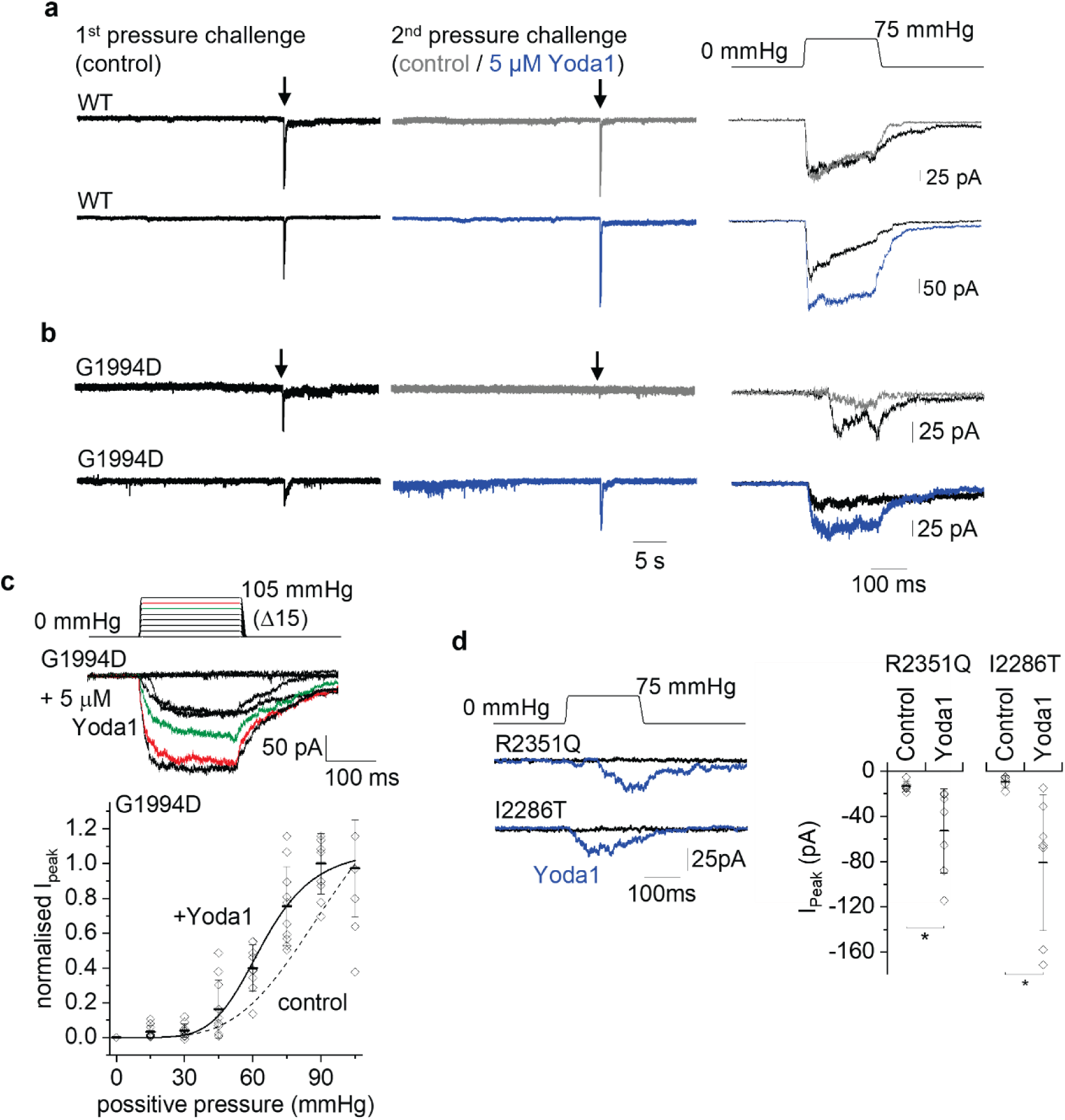
Synergy with mechanical activation. Data for outside-out patch recordings from T-REx-293 cells transfected with the indicated wild- type (WT) or variant mPIEZO1 constructs. The voltage across the patches during recordings was -80 mV. (a) On the left are sections of continuous current recording for WT channels, during which single pressure steps to 75 mmHg were applied as indicated by arrows. On the right are the currents during the pressure steps on expanded time-base and using the same colour code as on the left. (b) As for (**a**) but G1994D variant channel. (c) Upper: Schematic of the pressure pulse protocol in which a 200-ms pressure pulse was applied to 15 mmHg and then, every 12 s, incremented (Δ) in steps of 15 mmHg up to a maximum of 105 mmHg. Middle: Example ionic currents from a patch excised from a cell expressing G1994D variant mPIEZO1 in the presence of 5 μM Yoda1. Currents for 75 and 90 mmHg are in green and red respectively. Lower: Quantification of peak current amplitude for all G1994D experiments of this type, plotted across the pressure range, normalised to the current at 90 mmHg in each experiment and shown as mean ± s.d. with individual data points for each experiment superimposed (n = 9-10). The smooth curve is a fitted Boltzmann function with mid-point (P_50_) of 64.6 mmHg. The dotted curve is the Boltzmann fit to data for this variant without Yoda1 (-Yoda1) from Figure 3h. (d) Left: Similar to the data in (**b**) but for R2351Q and I2286T variants. Right: For experiments of the type shown on the left, maximum amplitude of current evoked by pressure steps represented as mean ± s.d. with each independent data point superimposed (n = 5-7). \**P*<0.05.

### Synergy with hypo-osmolality

Hypo-osmolality may occur in oedema, affecting lymphatic function and enhancing PIEZO1 activity ^45, 46^. In empty-vector transfected HEK 293 cells, there are small Ca^2+^ elevations in response to hypo-osmolality and these signals are enhanced when WT channels are transfected in (Figure 7a), consistent with the suggestion that PIEZO1 activity is stimulated by hypo-osmolality ^46^. In the presence of Yoda1, hypo-osmolality responses are larger, despite the elevated baseline signal caused by Yoda1 (Figure 7b). All 4 variant channels show hypo-osmolality responses in Yoda1’s presence (Figure 7c) but not in its absence (SI Figure S10). The Yoda1 concentration-response curve for WT channels does not reach a maximum in the hypo-osmolality condition and the curves for variant channels are now similar to those for WT channels except for the modest right-shifts of R2351Q and I2286T channels (Figure 7d, SI Figure S10). The data suggest that pharmacologically rescued variant channels are sensitive to hypo-osmolality as well as mechanical force, which could enable their preferential activation in oedema.

**Figure 7:**
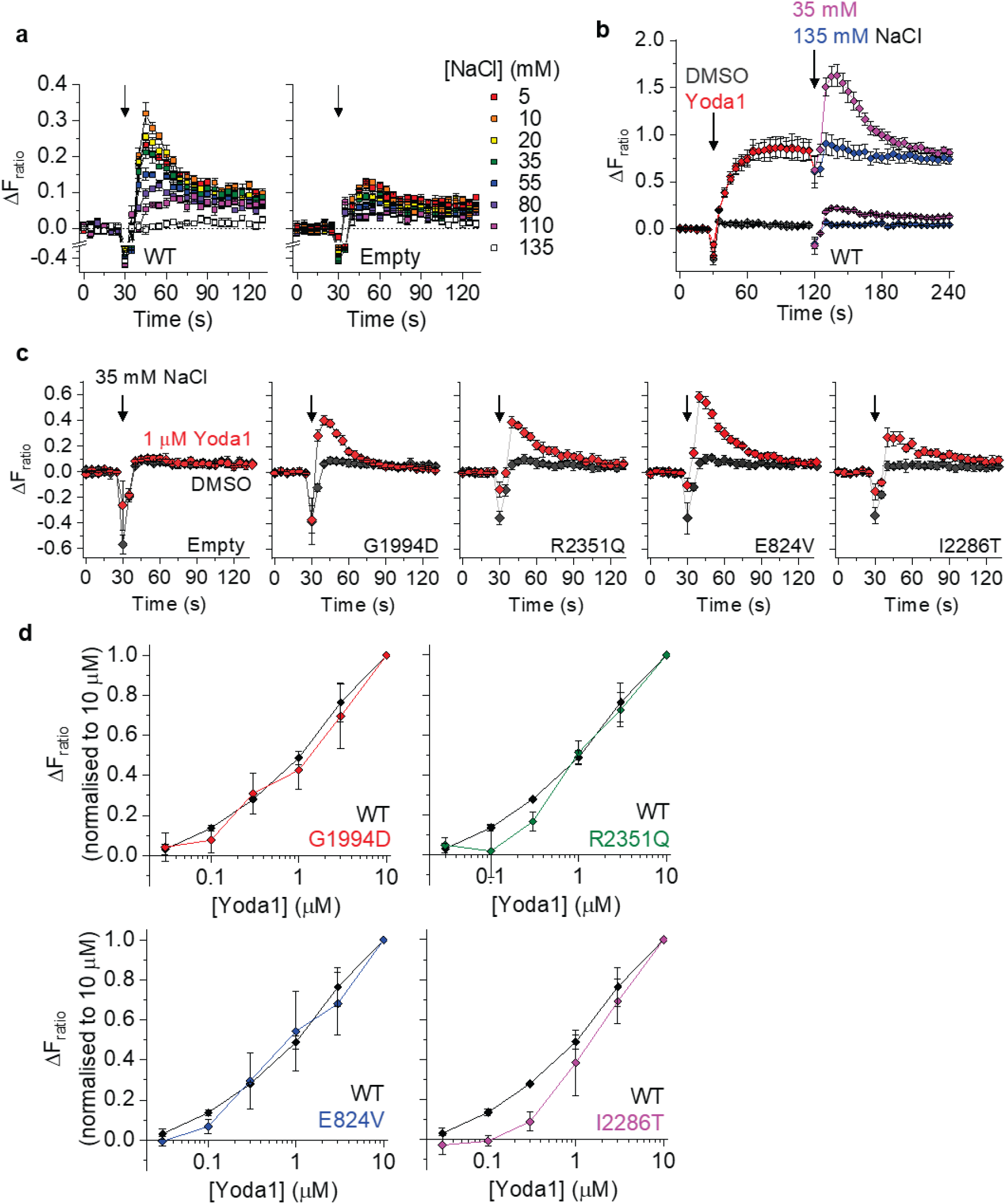
Synergy with the effect of hypo-osmolality. Ca^2+^ measurement data for T-REx-293 cells transfected with empty vector (Empty) or wild- type (WT) or variant mPIEZO1 constructs. Data are displayed as the change (Δ) in intracellular Ca^2+^ as indicated by change in the fura-2 fluorescence (F) ratio (ΔF_ratio_), with increase in ratio indicating increase in Ca^2+^ concentration in the cytosol. (**a**-**c**) Data are from single representative 96-well plate experiments exposed to the conditions indicated (mean ± s.e.m., N = 4-5 wells each). (a) Extracellular buffers were injected containing decreasing concentrations of NaCl starting at the arrow in the graphs and following the colour code on the right. The injected volume was equal to that of the 135 mM NaCl buffer initially bathing the cells. (b) For WT channels, responses to 0.1 μM Yoda1 or vehicle control (DMSO) and subsequent challenge with normotonic (135 mM NaCl) or hypotonic (35 mM NaCl) solution (in equal volume) according to the colour code indicated. (c) Responses to hypotonic challenge (35 mM NaCl) at the arrow, in the continuous presence of 1 μM Yoda1 (red) or vehicle control (DMSO, black) for the empty vector-transfected or variant-transfected cells. (d) Mean ± s.d. responses to hypo-osmolality (35 mM NaCl in equal volume to 135 mM NaCl buffer) in increasing concentrations of Yoda1 for WT and variant channels as indicated, normalised to the hypo-osmolality response in the presence of 10 μM Yoda1 (n = 3-6). Example individual data sets are shown in SI Figure S10 (“Further data for the effect of hypo- osmolality”).

### Thiazole modification of Yoda1 improves rescue

Seven novel Yoda1 analogues (SI Figure S11) and Dooku1 (KC41) ^30^ were tested for their ability to enhance the hypo-osmolality and sustained Ca^2+^ responses beyond those of Yoda1 (SI Figure S12). The thiazole analogue, CHR-1871-032, shows similar or sometimes larger sustained activation and increased hypo-osmolality responses (SI Figure S12). Short-duration exposure to CHR-1871-032 generates an EC_50_ of 0.14 μM at WT channels (Figure 8a, b), which is better than that of Yoda1 (Figure 4c). Construction of complete concentration-response curves is enabled for 3 of the variant channels, revealing EC_50_s of 0.71, 0.76 and 1.0 μM for I2286T, E824V and G1994D channels respectively (Figure 8a, b). The I2286T channel is now the most potently activated variant channel. R2351Q channel activates but the curve does not saturate, preventing EC_50_ determination (Figure 8a, b). Side-by-side comparison of the effects of Yoda1 and CHR-1871- 032 further supports the suggestion that CHR-1871-032 improves rescue of variant channels (Figure 8c). Five of the other analogues were tested for effectiveness at variant channels (SI Figure S12). None shows better rescue than Yoda1 but analogue KC183 has similar effect to Yoda1 at the G1994D variant channel (SI Figure S12). The KC183 response is slow (SI Figure S12), a feature that may be useful for generating prolonged responses *in vivo*. The data suggest opportunities for improved rescue of variant channels through Yoda1 modification.

**Figure 8:**
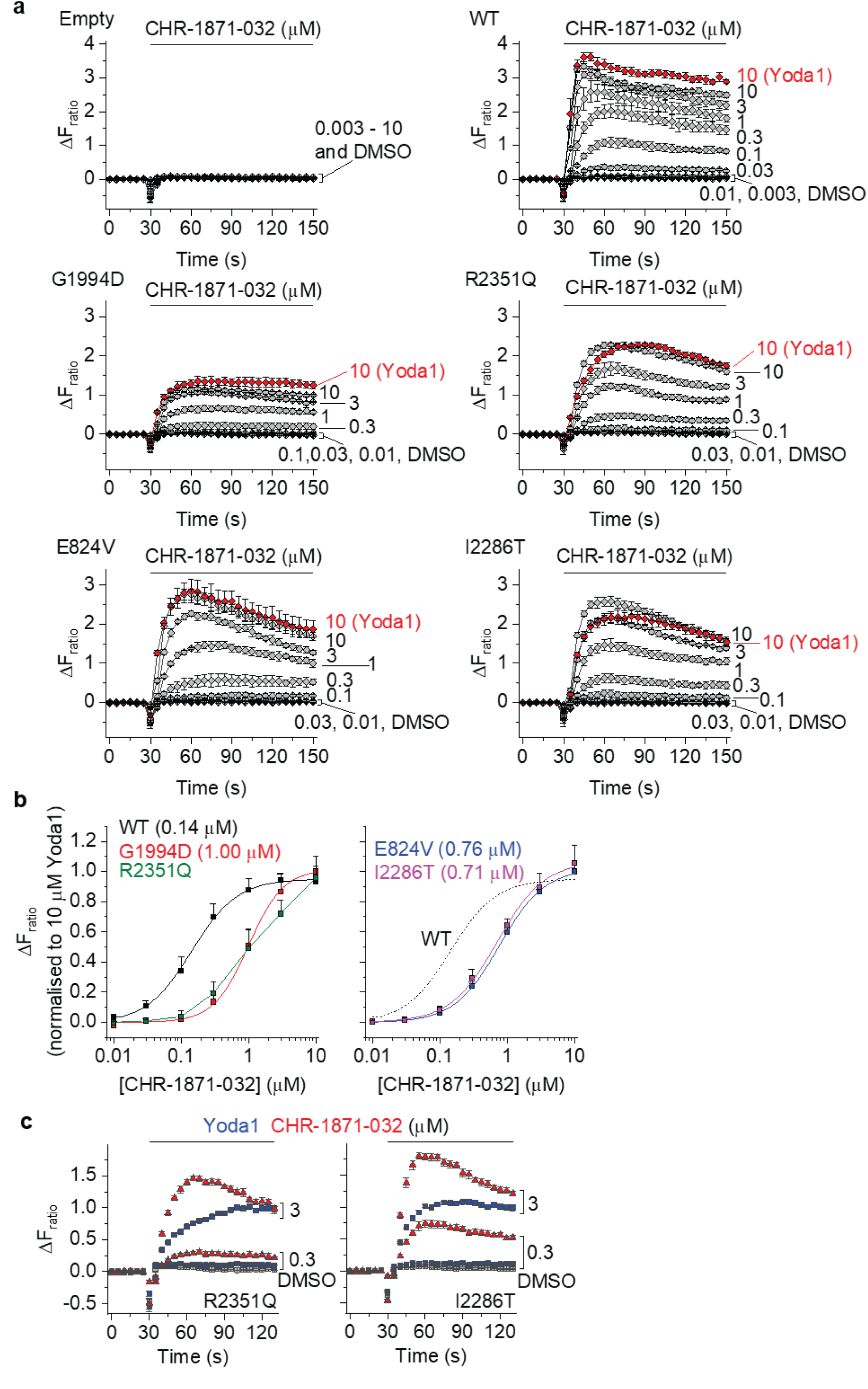
Enhanced rescue by a thiazole derivative of Yoda1 (CHR-1871-032) Ca^2+^ measurement data for T-REx-293 cells transfected with empty vector (Empty) or wild- type (WT) or variant mPIEZO1s. Data are displayed as the change (Δ) in intracellular Ca^2+^ as indicated by change in the fura-2 fluorescence (F) ratio (ΔF_ratio_), with increase in ratio indicating increase in Ca^2+^ concentration in the cytosol. (**a**, **c**) Data from single representative 96-well plate experiments exposed to the indicated CHR-1871-032 or Yoda1 concentrations in μM or vehicle (DMSO) control (mean ± s.e.m., N = 4-5 wells each). (b) Summary concentration-response data for experiments of the type shown in (**a**) with Hill equations fitted to the data except for R2351Q data, which are joined by straight lines (n = 3 each). The fitted Hill equations yielded EC_50_ values of 0.14 μM (WT), 1.00 μM (G1994D), 0.76 μM (E824V) and 0.71 μM (I2286T). (c) Example side-by-side comparison of the effects of CHR-1871-032 and Yoda1 tested against R2351Q and I2286T variant channels (mean ± s.e.m., N = 4-5 wells each).

### Similar channel properties in lymph node endothelial cells

To investigate if endogenous channels have characteristics comparable to those of overexpressed channels, an endothelial cell line derived from lymph nodes (SVEC4-10 cells) was studied. These cells show transient and sustained Ca^2+^ elevations in response to Yoda1, the concentration-dependence of which shifts to the left with prolonged exposure (Figure 9a, b). There is partial dependence on extracellular Ca^2+^ and partial inhibition by thapsigargin (Figure 9c, d). Yoda1 responses are not inhibited by ATP or TRPV4 inhibitors (SI Figure S13) but enhanced by hypo-osmolality (Figure 9e). KC183 causes a slower and more sustained response than Yoda1 (Figure 9f, g). The data suggest that characteristics seen for channels overexpressed in the HEK 293 cell line exist also for endogenous channels of lymph node endothelial cells.

**Figure 9:**
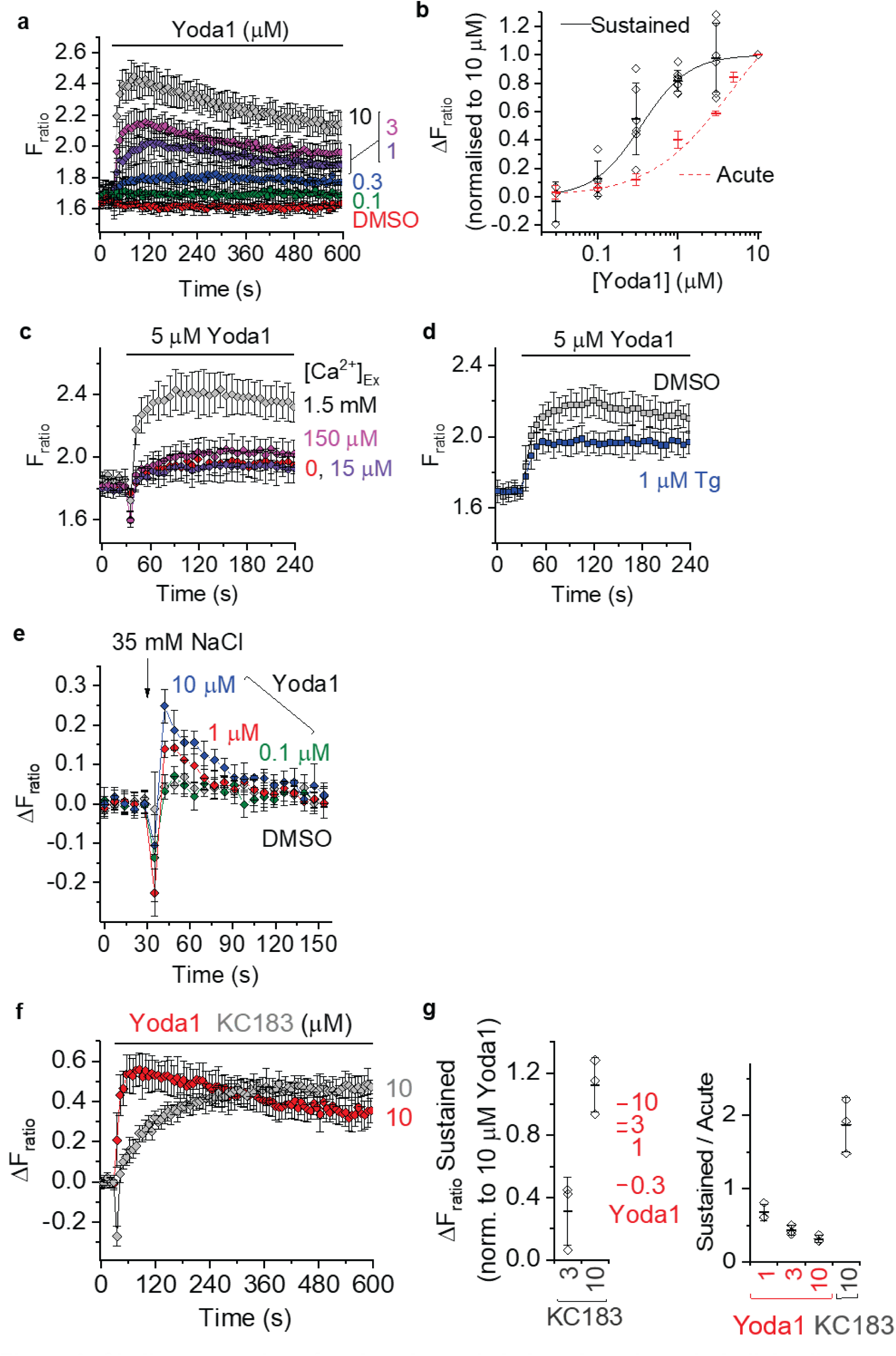
Similar properties of native channels in lymph node endothelial cells. Ca^2+^ measurement data for an endothelial cell line derived from lymph nodes (SVEC4-10 cells). Data are displayed as the intracellular Ca^2+^ concentration indicated by absolute or change (Δ) in fura-2 fluorescence (F) ratio (F_ratio_ or ΔF_ratio_), with increase in ratio indicating increase in Ca^2+^ concentration in the cytosol. (a) Data from a single representative 96-well plate experiment showing effects of increasing concentrations of Yoda1 or DMSO over the prolonged period up to 10 min (mean ± s.e.m., N = 4-5 wells each). (b) Summarised mean ± s.d. Yoda1 concentration-response data and individual data points superimposed comparing the responses to acute (30-60 s) and sustained (≥ 20 min) exposure to Yoda1 (n = 3-8). The Hill equation fitted to the sustained exposure data yielded an EC_50_ of 0.35 μM. (**c**-**f**) Data from single representative 96-well plate experiments, showing effects of (mean ± s.e.m., N = 4-5 wells each): (**c**) 5 μM Yoda1 in the presence of different concentrations of extracellular Ca^2+^ ([Ca^2+^]_Ex_) as indicated in the figure panel; (**d**) 5 μM Yoda1 in the presence of 1 μM thapsigargin (Tg) or its vehicle control (DMSO); (**e**) hypo-osmolality (35 mM NaCl) in the presence of different concentrations of Yoda1 or its vehicle control (DMSO) as indicated in the figure panel; or (**f**) 10 μM Yoda1 or KC183. (**g**) Left: Sustained responses after ≥ 20 min exposure to 3 or 10 μM KC183 normalised to the 10 μM Yoda1 response. Mean Yoda1 response values are indicated in red for comparison. Right: Sustained (≥ 20 min) response as a ratio of acute (30-60 s) response to Yoda1 and KC183 at the indicated concentrations in μM. Data are presented as mean ± s.d. with individual data points superimposed (n = 4-7).

## DISCUSSION

Autosomal recessive pathogenic variants in *PIEZO1* have been described in patients with NIFH and/or generalised lymphatic dysplasia (GLD). Interpretation of the variants in this large, highly polymorphic gene has been consistently difficult particularly if there is a missense variant. Accurate genetic diagnosis is important for prognostication and recurrence risks. With many of the variants in this gene (and other genes), a tool to assess pathogenicity is critical for interpretation. Furthermore, little was previously understood about the underlying mechanism resulting in lymphatic failure in this condition.

This study reveals four previously unreported missense variations in the *PIEZO1* gene that associate with GLD. Full-length PIEZO1 expression is still possible with these variants, but the single amino acid changes lead to less or abolished PIEZO1 channel activation by mechanical force as the possible disease mechanism.

Similar clinical presentations in the two generations of the GLD08 family is suggestive of autosomal dominant inheritance. However, sequencing identified that both generations are biallelic for I2270T, which is shown here to cause PIEZO1 loss-of-function. This confirms a pseudo-dominant mode of inheritance which is due to a high level of consanguinity, although the genotype of the parents of GLD08 I.2 could not be confirmed.

In the GLD09 family, the proband presents with a typical GLD phenotype and a biallelic genotype (compound heterozygous for E829V and I2270T). The I2270T variant is maternally inherited and the mother (GLD09 I.2) is unaffected. However, the father (GLD09 I.1) who is monoallelic for the E829V variant has a history of congenital onset primary lymphoedema. If the lymphoedema symptoms in the father are related to *PIEZO1* defects, the paternally inherited variant E829V must exert a different disease mechanism, which would most likely be dominant negative. Results of our co-expression studies of E829V with WT PIEZO1 support this idea (Figure 2d). This type of gain-of-function (i.e., dominant negative effect) is antimorphic and thus different to the hypermorphic gain-of-function observed in individuals with autosomal dominant *PIEZO1*-associated DHS ^47^.

Co-inheritance of a loss-of-function variant with a dominant negative gain-of-function variant *in trans* in the *ITGB3* gene has been reported as a cause of Glanzmann thrombasthenia ^48^, where family members carrying the dominant negative variant also present with symptoms, although milder than proband. While the symptoms appear milder in the GLD09 father, we do not have enough evidence to make firm conclusions, but this study suggests that primary lymphoedema can be caused by monoallelic *PIEZO1* variants, while a more severe GLD phenotype is caused by biallelic inheritance of variants, which can be either two loss-of-function variants or a compound of co-inherited loss-of-function and dominant negative gain-of-function variants. Our study will enable clinicians in reclassifying the *PIEZO1* VUS. It has also opened our eyes to the complexity of *PIEZO1* variant-associated disease. The possibility of antimorphic acting *PIEZO1* missense variants leading to autosomal dominant primary lymphoedema without DHS symptoms is going to make the decision-making more difficult in the clinical genetic setting when there are also hypermorphic acting *PIEZO1* missense variants that can lead to DHS, which recently was also reported to associate with primary lymphoedema ^49^.

We show that PIEZO1 channel activity can be rescued by Yoda1 and Yoda1-related small-molecule agonists, and that these rescued channels show mechanical and hypo-osmolality sensitivities, suggesting their potential for physiological regulation once stimulated by an agonist. Some of these faulty proteins are partly intracellular Ca^2+^ release channels, suggesting that increased membrane permeability of an agonist can enhance rescue by its ability to also access these channels. Improved rescue by new Yoda1 analogues is shown, suggesting foundations for new pharmacology aimed at overcoming problems of GLD and potentially addressing lymphatic insufficiency in other types of lymphoedema because of the widespread importance of PIEZO1 generally in lymphatic biology.

Yoda1 accelerates lymphatic valve formation in newborn mice ^26^ and suppresses postsurgical lymphoedema in mice ^25^, encouraging the idea that PIEZO1 agonism could be a therapeutic strategy for problems of lymphatic function beyond rare cases of GLD. Effectiveness of PIEZO1 agonists is likely to depend on factors such as the availability of PIEZO1 channels and lymphatic structures for modulation. Assays that determine or predict these factors will inform the tailoring of therapies to patients who are most likely to benefit. We anticipate that therapies of this type are likely to be safe to administer at an appropriate dose because *PIEZO1* gain-of-function variants are common in some human populations and without major adverse effect ^50–52^. Safety of administering agonists will, however, depend on the specific chemistry and any off-target (i.e., non-PIEZO1) effects. Understanding of PIEZO1 agonist structure-activity relationships is emerging but the field is still in its infancy. As shown elsewhere ^30, 40, 53, 54^ and here, better agonism and improved physicochemical properties of agonists are possible based on the Yoda1 template.

Understanding of the molecular mechanisms of PIEZO1 channels is also in its infancy ^16^. The variants we identified are in regions of the channel that are particularly poorly understood. The CED-located variants have the most damaging effects on mechanical activation, yet mechanical sensitivity is thought to arise in the blades. The CED is attached to and sits above the ion pore but also projects feet-like structures to the blades ^11^. We speculate that the CED variants somehow stiffen the CED and its associated blade regions, thereby right-shifting mechanical sensitivity out of the physiological range. Yoda1 left-shifts mechanical-response curves ^44^, so Yoda1 and Yoda1-related agonists may act by bringing the channels back into range for physiological regulation. The variants in the blades may similarly cause stiffening of the channels. However, we need new structural data to test such hypotheses and specific structural information for GLD variants and their modulation. If atomic resolution can be achieved in relevant parts of the channel, it may inform the design of new modulators.

Various mechanical and Yoda1 sensitivities are reported for PIEZO1 channels, with the vast majority of data being for mouse PIEZO1 ^13, 14, 30, 33, 40, 44^. In part, such variability may arise due to the recording technique and experimental conditions, which may affect, for example, the number of PIEZO1 channels per unit area of membrane and the membrane properties, which are likely to affect mechanical sensitivity because of PIEZO1’s lipid modulation ^33, 55–57^. The patch-clamp technique has several configurations ^58^ and presents multiple opportunities for force application ^16^, all of which may affect the estimates of mechanical sensitivity and in turn the sensitivity to Yoda1 ^44^. Comparisons in a single study showed that pressure sensitivity and inactivation in outside-out patch recordings are much less than those recorded in other patch-clamp configurations ^14^. We used the outside-out patch for our experiments and similarly observed absence of inactivation and apparently low pressure sensitivity. We chose this configuration because the channels are studied in relative isolation, in the absence of the main body of the cell and with reduced cytoskeletal interference. In this way, a potentially simpler perspective on the mechanical sensitivity should be achieved. The absolute values obtained may, however, be different from those in more physiological conditions. Our hypo-osmolality studies provide independent perspective on this situation because they were performed on intact cells without the invasive impact of the patch pipette. The mechanism of action of hypo-osmolality on the PIEZO1 channels is unclear, but cell-swelling may be caused by hypo-osmolality as water moves into the cells, increasing membrane tension. Even in the relatively non-invasive situation of the calcium assays, variabilities in the membrane properties and mechanical condition of the cells may impact the absolute values obtained. We observed Yoda1 sensitivity to vary even between similar Ca^2+^ assays and using the same cell line; this is evident when the exact Yoda1 EC_50_ values are compared across our studies here and published elsewhere ^30, 40^. Therefore, we emphasize the importance of internal controls within the same study, as we did here.

In summary, we have shown clear pathogenicity in these four missense variants initially classed as VUS, demonstrating the clinical utility of our methods. We suggest these variants express and surface localise as full-length protein but with reduced mechanically activated channel function which is the primary cause of the GLD and NIFH observed in the families under study. We also show for the first time a dominant negative disease-causing variant. Thus, caution must be exercised in interpretation of variants in this gene when disease presentation in a family does not conform to the expected inheritance pattern. We also show that small-molecule agonists can functionally rescue these dysfunctional channels, revealing channels that are physiologically regulated. Intracellular Ca^2+^ release by some of the variants suggests value in membrane-permeable agonists that access this internal pool of channels as well as those at the surface membrane. It is possible to improve on the Yoda1 agonist through chemical modification. Therefore, the cases presented here are examples of GLD in which the underlying molecular defect could be overcome by small-molecule modulators, and thus potentially therapeutic drugs based on such modulators. However, GLD is a rare disease and not all cases are caused by such *PIEZO1* variants, but the principles of functional PIEZO1 rescue presented here could potentially have a broader relevance as a way to overcome suboptimal functionality in the lymphatic system not just in *PIEZO1* variant-associated GLD, but other types of lymphoedema.

## METHODS

### Patient ascertainment

Three individuals with a GLD phenotype not previously reported and available family members were included in the study. Genomic DNA was analysed for sequence variants in all exons of *PIEZO1* by Sanger sequencing as described previously ^7^. Samples of available family members were subsequently analysed by Sanger sequencing for the variants identified in their respective proband. Findings were subsequently confirmed in a molecular diagnostics laboratory.

### *PIEZO1* variants *in silico* analysis

All the relative genomic and protein positions of PIEZO1 reported here correspond to the transcript PIEZO1-001 (RefSeq: NM_001142864, Ensembl: ENST00000301015.9) and Q92508 Uniprot protein accession ID, respectively. The reported genomic coordinates refer to the GRCh38/hg38 human genome reference. Changes in the gene structure and/or amino acid sequence, due to the reported variants, were retrieved by the Refgene database of UCSC ^59^. The Allele Frequencies (AF) of the reported variants were checked in gnomAD databases ^60^ and their pathogenicity was predicted by the Combined Annotation Dependent Depletion (CADD) tool ^61^, Mutation Taster ^62^, SIFT ^63^ and PolyPhen2^64^.

### PIEZO1 constructs

Human PIEZO1_AcGFP ^38^ was used as a template to clone the human PIEZO1 (hPIEZO1) sequence with a C-terminal HA-epitope. Overlapping hPIEZO1 (forward primer 5’ GGTCTACCTGCTCTTCCTGCTG 3’ and reverse primer incorporating a ‘ASA’ linker and the HA sequence a 5’ CCCATACGATGTTCCAGATTACGCTTAGGCGACTCTAGATCATAATCAGCCATACC 3’) and vector (forward primer 5’ CCCATACGATGTTCCAGATTACGCTTAGGCGACTCTAGATCATAATCAGCCATACC 3’ and reverse primer 5’ CAGCAGGAAGAGCAGGTAGACC 3’) PCR products were assembled using Gibson Assembly. Missense variants were introduced by site-directed mutagenesis (PCR primer sequences are provided in SI Table S2). pcDNA3_mouse PIEZO1_IRES_GFP, a gift from A Patapoutian, was used as a template to clone the mouse PIEZO1 (mPIEZO1) coding sequence with a C-terminal HA-epitope, into pcDNA™4/TO. Overlapping mPIEZO1 (forward primer 5’ GTAACAACTCCGCCCCATTG 3’ and reverse primers 5’ CTAAGCGTAATCTGGAACATCGTATGGGTACTCCCTCTCACGT 3’) and pcDNA™4/TO (forward primer 5’ CATACGATGTTCCAGATTACGCTTAGCCGCTGATCAGCCTCG 3’ and reverse primer 5’ CAATGGGGCGGAGTTGTTAC 3’) PCR products (PrimeSTAR HS DNA Polymerase, TaKaRa) were assembled using Gibson Assembly (NEB). Missense variants were introduced into mPIEZO1 with and without HA-epitope by site-directed mutagenesis (PCR primer sequences are in SI Table S2).

### Cell culture

HEK 293 cell line was transiently transfected at 90% confluence with 500 ng DNA using Lipofectamine 2000 (Invitrogen) in OptiMEM (Gibco) according to the manufacturer’s instructions. For co-expression experiments, cells were transfected with only 75 ng of each construct for submaximal expression of each subunit. Medium was replaced after 5 hours and cells were used for experimentation 48 hours after transfection. T-REx-293 cell line was transfected with pcDNA4/TO-mPIEZO1 constructs using Lipofectamine 2000 (Invitrogen) and treated with 200 μg.mL^-1^ zeocin (InvivoGen) to select for stably transfected cells. At least two strongly expressing clonal cell lines were established and tested for each variant. All cell lines were maintained in Dulbecco’s Modified Eagle’s medium (Invitrogen) supplemented with 10% foetal calf serum (Sigma-Aldrich), penicillin (50 units.mL^-1^) and streptomycin (0.5 mg.mL^-1^) (Sigma-Aldrich) and grown at 37 °C in a 5% CO_2_ incubator.

### Western blotting and cell surface biotinylation

For western blotting, cells were harvested in lysis buffer (10 mM Tris, pH 7.4, 150 mM NaCl, 0.5 mM EDTA, 0.5% Nonidet P40 substitute) containing protease inhibitor cocktail (Sigma). Equal protein amounts were loaded on 7% polyacrylamide gels and resolved by electrophoresis. Samples were transferred to PVDF membranes and labelled overnight with anti-HA (0.01 μg.mL^-1^, Roche clone 3F10), anti-β- actin (200 ng.mL^-1^, Santa Cruz) or anti-CD71 (1:5000, Cell Signaling Technology clone D7S5Z). Horseradish peroxidase donkey anti-mouse/rabbit/rat secondary antibodies (1:10000, Jackson ImmunoResearch) and SuperSignal Femto detection reagents (Pierce) were used for visualisation. For cell surface biotinylation, cells were grown in 6-well plates to confluency, washed twice with ice-cold PBS containing 1 mM CaCl_2_ and 1 mM MgCl_2_ and biotinylated on ice for 30 minutes using 0.22 mg.mL^-1^ EZ-Link Sulfo-NHS-Biotin (Pierce) in PBS/1 mM CaCl_2_/1 mM MgCl_2_. Subsequently, cells were washed three times with ice-cold PBS/1 mM CaCl_2_/1 mM MgCl_2_ containing 50 mM glycine to quench excess biotin, and finally once with ice-cold PBS/1 mM CaCl_2_/1 mM MgCl_2_. Cells were harvested in lysis buffer (10 mM Tris, pH 7.4, 150 mM NaCl, 0.5 mM EDTA, 0.5 % Nonidet P40 substitute) containing protease inhibitor cocktail (Sigma). Biotinylated proteins were concentrated from 300 μg total protein using streptavidin-agarose (Pierce) overnight at 4 °C. Agarose beads were washed three times with ice-cold lysis buffer and bound proteins eluted using sample buffer (4x SB: 250 mM Tris pH 6.8, 8% SDS, 40% glycerol, 8% βmercaptoethanol) and heating at 70°C (higher temperatures resulted in PIEZO1 protein aggregation).

### Intracellular Ca^2+^ measurement

For intracellular Ca^2+^ assays, cells were plated at 80-90% confluence in 96-well plates 24 hours prior to recordings (6-8 x 10^4^ cells per well). To measure intracellular Ca^2+^, cells were incubated for 1 hour at 37 °C in standard bath solution (SBS) of composition 135 mM NaCl, 5 mM KCl, 1.2 mM MgCl_2_, 1.5 mM CaCl_2_, 8 mM glucose and 10 mM HEPES (pH titrated to 7.4 using NaOH) containing 2 μM fura-2 acetoxymethyl ester (fura- 2-AM, Molecular Probes) with 0.01% weight/volume pluronic acid. Cells were washed with SBS and incubated at room temperature for 20 minutes prior to recordings. Equal volumes of 2x concentrated compounds were injected to test for acute channel activation. Pre-incubation with compounds occurred during the 20 minutes prior to recordings. To expose cell membranes to increased hypo-osmolality, an equal volume of hypotonic SBS (containing only 5-110 mM NaCl) was injected onto the cells. Measurements were made on a fluorescence plate reader (Flexstation III, Molecular Devices) at room temperature (21 ± 2 °C). Fura-2 was excited at 340 nm and 380 nm and emitted light collected at 510 nm, with measurements shown as the change in fluorescence (F) ratio (ΔF_340/380_).

### Electrophysiology and mechanical stimulation

Ionic currents were recorded through outside-out patches from cells using standard patch-clamp techniques in voltage-clamp mode. Patch pipettes were fire-polished and had resistance of 4–7 MΩ when filled with pipette solution. Ionic solution of composition (mM) NaCl 140, HEPES 10 and EGTA 5 (titrated to pH 7.4 using NaOH) was used in both the pipette and bath. Recordings were at a constant holding potential of −80 mV. 200-ms pressure steps were applied to the patch pipette with an interval of 12 s using High Speed Pressure Clamp HSPC-1 System (ALA Scientific Instruments, USA). All recordings were made with an Axopatch-200B amplifier (Axon Instruments, Inc., USA) equipped with Digidata 1550B and pClamp 10.6 software (Molecular Devices, USA) at room temperature (21 ± 2 °C). Currents were filtered at 2-5 kHz and digitally sampled at 5-20 kHz.

### Molecular modelling

Structural data for mPIEZO1 were obtained at Protein Data Bank (PDB:6B3R). Missing loop regions were modelled using MODELER (v9.19). Large intracellular loops (located at residues 718-781, 1366-1492, 1579-1654 and 1808-1951) remained unstructured following modelling and were removed from the final model. A structural model of hPIEZO1 was generated based on the mPIEZO1 model.

### Chemistry

KC41 (Dooku1) was prepared as described ^30^. All purchased chemicals and solvents were used without further purification unless otherwise stated. All compounds were at least 95% pure by 1H NMR. 1H Nuclear Magnetic Resonance spectra were recorded at 500 MHz using a Bruker DRX 500 instrument or at 400 MHz using a Bruker DPX 400. 1H spectra are referenced based on the residual proton in the solvent (e.g., the CHCl_3_, 0.01 % in 99.99 % CDCl_3_). Coupling constants (J) are reported to the nearest 0.1 Hz. 13C NMR spectra were recorded at 125 MHz on 500 MHz spectrometers or at 100 MHz on 400 MHz spectrometers. LC-MS was performed on a Bruker Daltronics running a gradient of increasing acetonitrile (5 to 95 %) in H_2_O both containing 0.1 % formic acid at 1 ml.min^-1^, on a short path C18 reverse phase column, detecting compounds with both a diode array detector and a Bruker mass spectrum analyser. HRMS was performed on a Bruker Daltonics micrOTOF using positive electrospray ionisation (ES+). Automated column chromatograph was carried out using a Biotage Isolera Four, using either Sfär Silica D or KP-Silica cartridges. HPLC was performed on an Agilent 1290 Infinity Series equipped with a UV detector and Hyperprep C18 reverse phase column. Key to NMR abbreviations: s (singlet), br s (broad singlet) d (doublet), dd (doublet of doublets), ddd (doublet of doublets of doublets), t (triplet), dt (doublet of triples), q (quartet), m (multiplet), ap. t (apparent triplet). Chemistry details for each novel analogue are provided in the SI Chemistry section.

### Statistical data analysis

Averaged data are shown as mean ± standard error of the mean (s.e.m.) or standard deviation (s.d.), as specified in the figure legends. Data were produced in pairs and analysed statistically with Student’s t-test or in groups and analysed by one-way ANOVA using Origin^R^2018 software (OriginLab). Statistically significant difference is indicated by \**P*<0.05, \*\**P*<0.01 or \*\*\**P*<0.001. n.s., indicates not significantly different. “n” is used to denote the number of independent data points. Representative traces contain “N” technical replicates from within a single experiment. For patch-clamp studies, data were analysed using pClamp 10.6, MicroCal Origin 2018 (OriginLab Corporation, USA) and GraphPad Prism (GraphPad Software, Inc., USA) software packages.

## DATA AND MATERIALS AVAILABILITY

The data that support the findings of this study are available from the corresponding author upon reasonable request. Some data may not be made available because of privacy or ethical restrictions. All original data are available in an Excel file. Unique laboratory materials created in the project are available on request (D.J.B. for biological materials and R.F. for chemicals).

## FUNDING STATEMENT

The work was supported by research grants from Wellcome (grant number 110044/Z/15/Z) and British Heart Foundation (BHF) (RG/17/11/33042, SP/13/5/30288) and a joint MRC/BHF program grant (MR/P011543/1 and RG/17/7/33217) and Newlife Foundation for Disabled Children (12-13/01). Studentships from University of Leeds (for K.C.) and the BHF (FS/4yPhD/F/20/34130 for K.A.S. and FS/18/78/33932 for E.D.). For the purpose of Open Access, the authors have applied a CC BY public copyright license to any Author Accepted Manuscript version arising from this submission.

## AUTHOR CONTRIBUTIONS

M.J.L. performed recombinant DNA experiments and mutagenesis, generated cell lines, designed and performed calcium measurement assays, made figures, orchestrated the figure designs, data analysis and data transparency, made intellectual contribution and wrote major parts of the first draft of the manuscript. O.V.P. designed and performed patch-clamp experiments, made figures and performed data analysis and made intellectual contribution.

D.M.L. designed and performed patch-clamp experiments, made figures and performed data analysis and made intellectual contribution. S.M.-A. supervised the genetic studies of patients, identified the genetic variants in patients and produced information for figures and tables associated with the patient genetics and clinical features, writing (review and editing), project administration. C.R. designed, synthesized and analysed chemicals. K.C. designed, synthesized and analysed chemicals. K.A.S. performed the computer modelling and made the associated figure panels. E.Fa. performed data curation and writing in the original draft. E.Fo. performed investigation (Sanger sequencing), formal analysis. A.B. performed investigation (seen patient in clinic) and provided resources (patients). C.H. performed investigation (seen patient in clinic) and provided resources (patients). T.L. performed investigation (seen patient in clinic) and provided resources (patients). N.T. performed investigation (seen patient in clinic) and provided resources (patients). S.W. performed investigation (seen patient in clinic), and provided resources (patients). E.D. performed formal analysis (variant interpretation) and writing (review and editing). S.M. performed investigation (seen most patients in clinic), and provided resources (patients), supervision of clinical staff and writing (review and editing). G.P. provided technical assistance. A.C.K. supervised the computer modelling. R.F. supervised the chemistry and helped to generate funds. P.O. supervised the genetic studies of patients, identified the genetic variants in patients, generated associated funding and produced information for figures and tables associated with the patient genetics and clinical features, writing (review and editing), conceptualization, resources, supervision, funding acquisition, coordination with clinical authors. D.J.B. conceptualised the study, made intellectual contribution, supervised and orchestrated the laboratory project and team, generated funding, interpreted data and wrote parts of the manuscript.

## CONFLICT OF INTEREST DISCLOSURE

D.J.B. and R.F. are partners of CalTIC GmbH, a pharmaceutical startup company with a mission to develop ion channel modulators as new classes of medicines. No other conflicts of interests are disclosed.

## ETHICS APPROVAL STATEMENT AND PERMISSION TO PUBLISH

Ethical approval for this study was obtained from the South West London Research Ethics Committee (REC Ref: 05/Q0803/257) and written informed consent was obtained from all participants. Permission to publish was also obtained.

## AUTHOR DETAILS

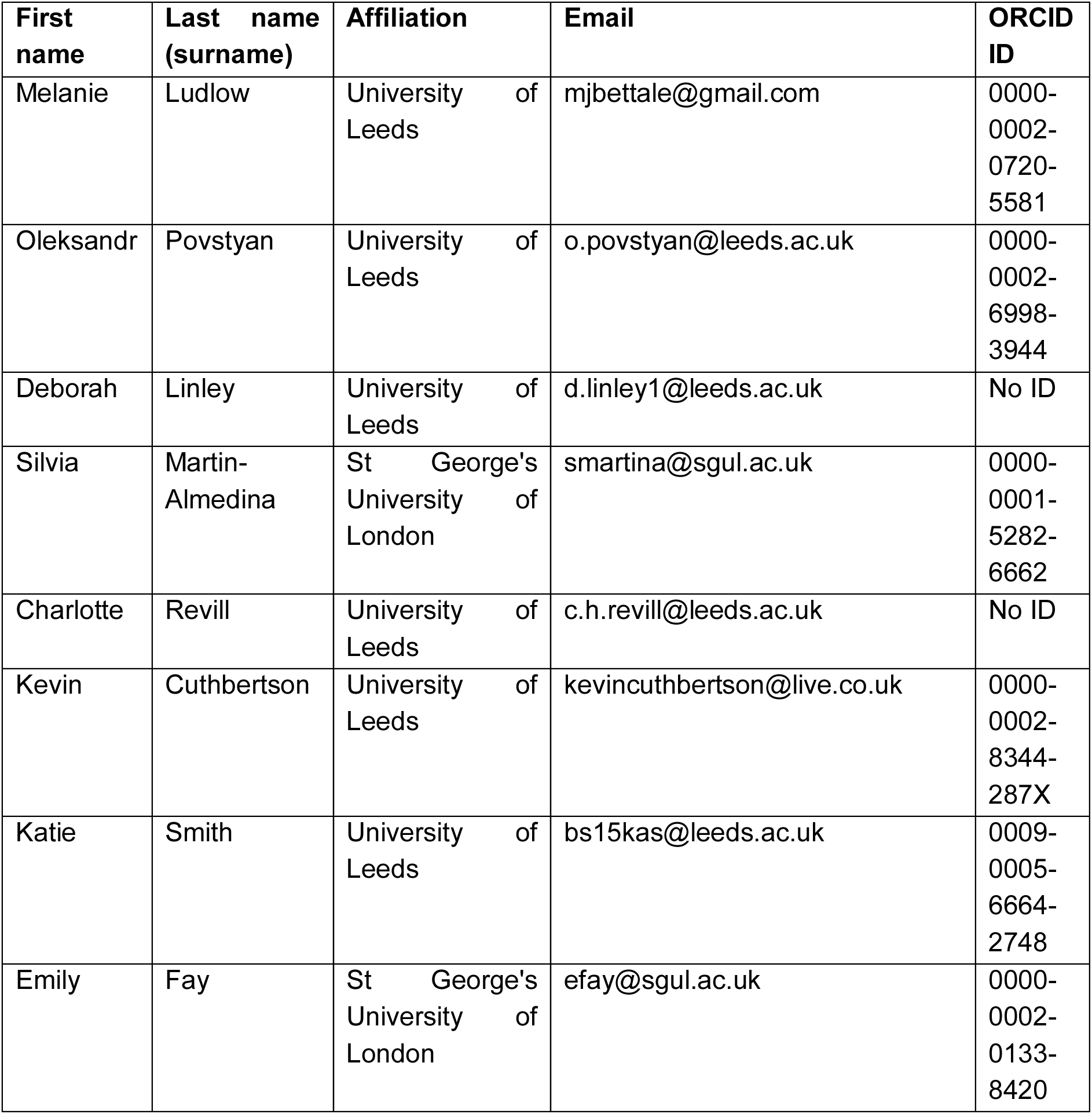
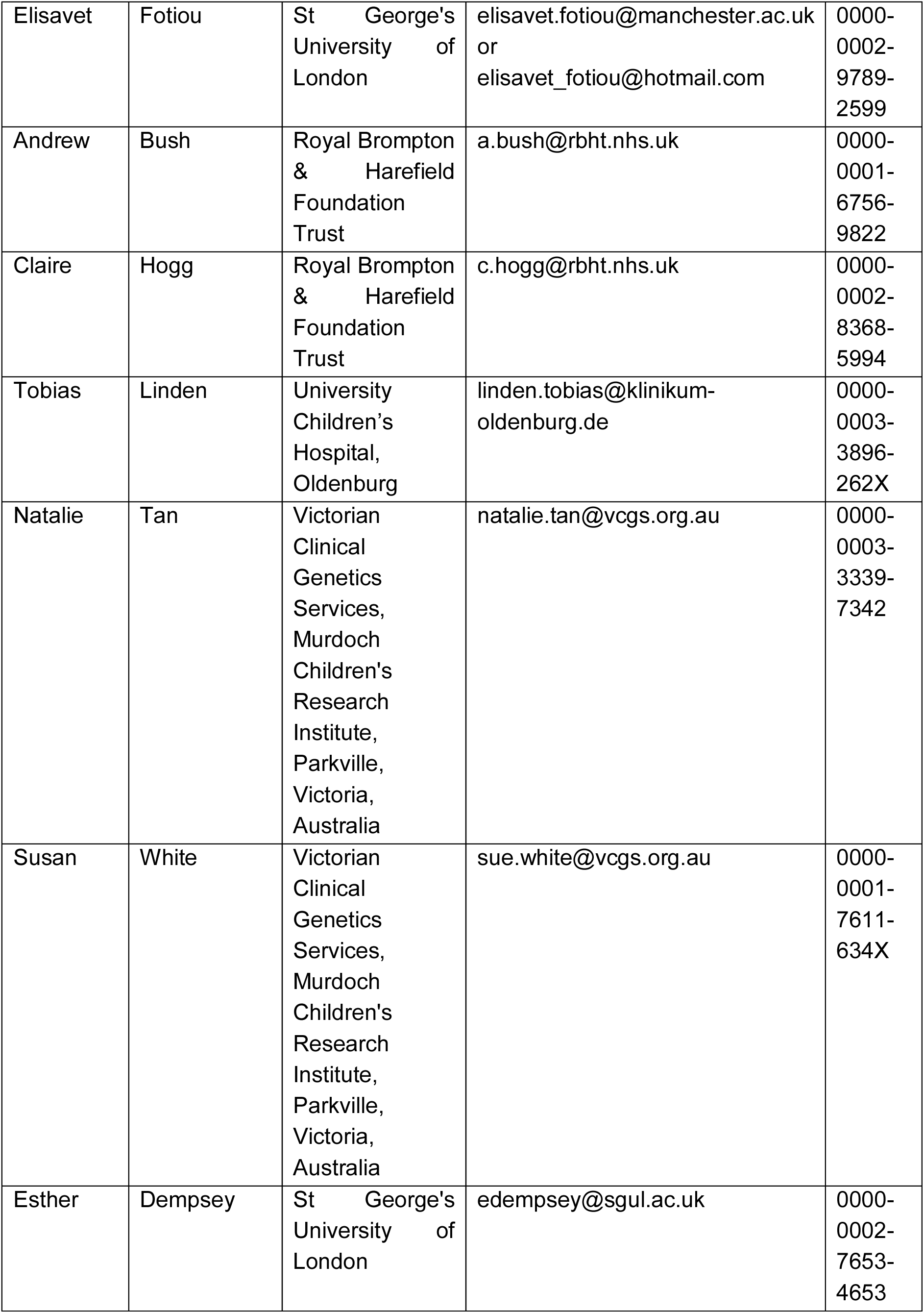
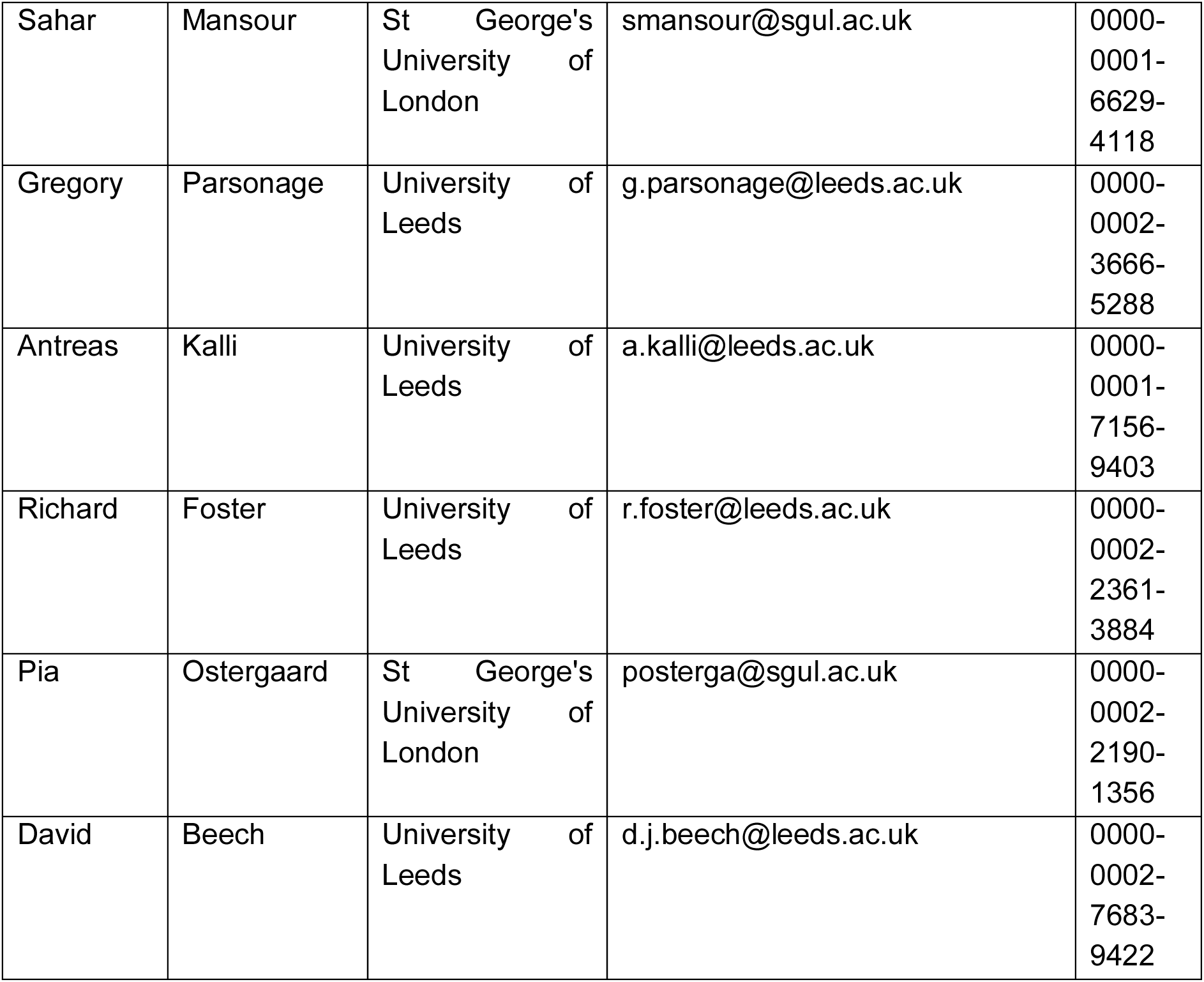

## SUPPLEMENTAL INFORMATION (SI)

**Table S1:** Annotation of the four *PIEZO1* variants investigated in this study. Genomic coordinates, nucleotide and protein changes, predicted pathogenicity and population allele frequencies are summarized. All variants are not reported in gnomAD databases or their allele frequency is infinitesimal and they are not found in a homozygous state, supporting the argument that they are extremely rare in the general population. They are all reported to be pathogenic or possibly damaging by most of the prediction tools used (CADD, PolyPhen2, MutationTaster, SIFT). AF, allele frequency; BP4, Multiple lines of computational evidence suggest no impact on gene or gene product; CED, C-terminal extracellular domain; GLD, generalised lymphatic dysplasia; TM, transmembrane; VUS, variant of uncertain significance; VUS+, strong variant of uncertain significance; PM2, absent from controls; PP1, cosegregation with disease; PP3, multiple lines of computational evidence support a deleterious effect; PP4, patient’s phenotype or family history is highly specific for a disease.

| ID | Genomic coordinates (GRCh38) | Nucleotide change | Predicted protein change | Domain affected | CADD Phred score | Polyphen2 HVAR rankscore | MutationTaster prediction | SIFT prediction | gnomAD v2.1 AF | gnomAD v2.1 Homozygous carriers | gnomAD v3 AF | gnomAD v3 Homozygous carriers | ACMG evidence code | ACMG score |
| --- | --- | --- | --- | --- | --- | --- | --- | --- | --- | --- | --- | --- | --- | --- |
| GLD09 | chr16:88733589-88733589 | c.2486A>T | E829V | TM | 29.4 | 0.732 | Pathogenic | Pathogenic | - | - | 6.98E-06 | 0 | PM2, PP3, PP4 | VUS |
| GLD07 | chr16:88720401-88720401 | c.5933G>A | G1978D | TM | 32 | 0.818 | Pathogenic | Pathogenic | - | - | - | - | PM2, PP3, PP4 | VUS |
| GLD08; GLD09 | chr16:88716676-88716676 | c.6809T>C | I2270T | CED | 24.6 | 0.693 | Pathogenic | Pathogenic | 6.54E-06 | 0 | - | - | PM2, (PP1), PP3, PP4 | VUS+ |
| GLD07 | chr16:88716406-88716406 | c.7004G>A | R2335Q | CED | 24.9 | 0.736 | Benign | Pathogenic | 1.97E-05 | 0 | - | - | BP4, PP3, PP4 | VUS |

**Table S2:**
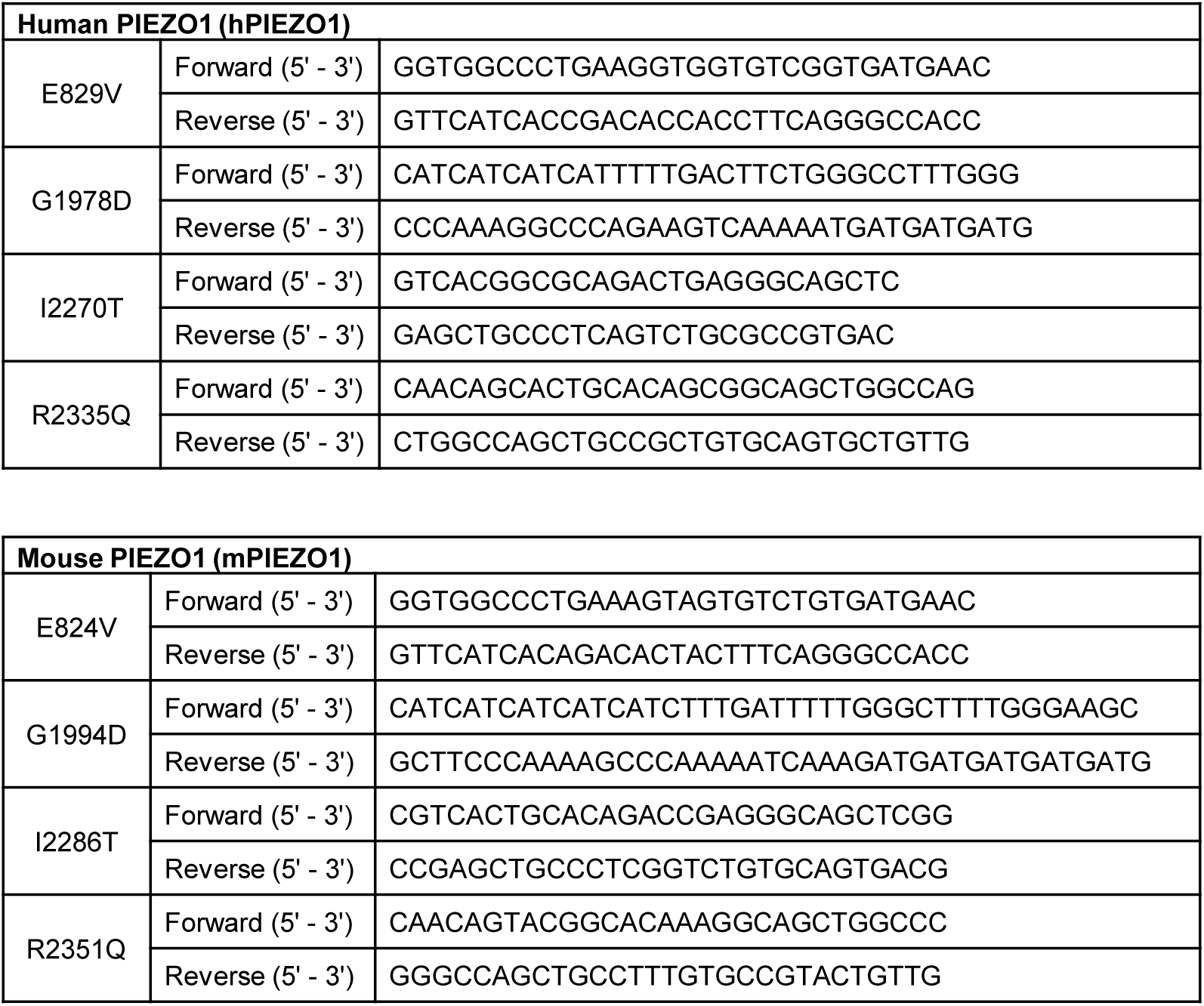
Primer sequences used to generate the PIEZO1 variants.

**Figure S1:**
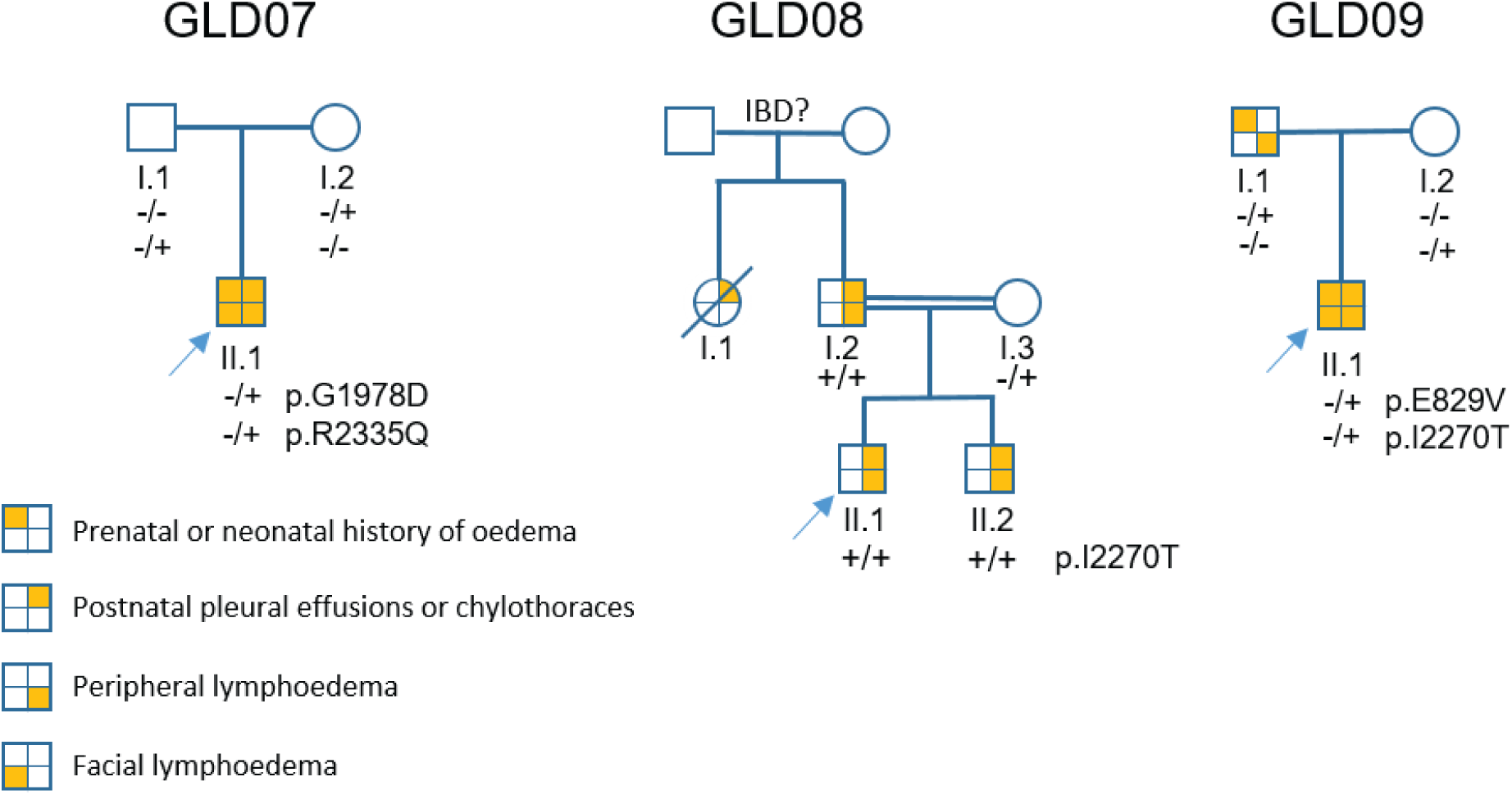
Pedigrees of the GLD families. Affected individuals are indicated with filled circles or squares. *PIEZO1* genotypes are indicated for individuals who underwent Sanger sequencing. The wildtype allele of the genotype is indicated by minus sign (-) and plus (+) represents the alternative allele. Arrows indicate proband. IBD?, unconfirmed identity by descent.

**Figure S2:**
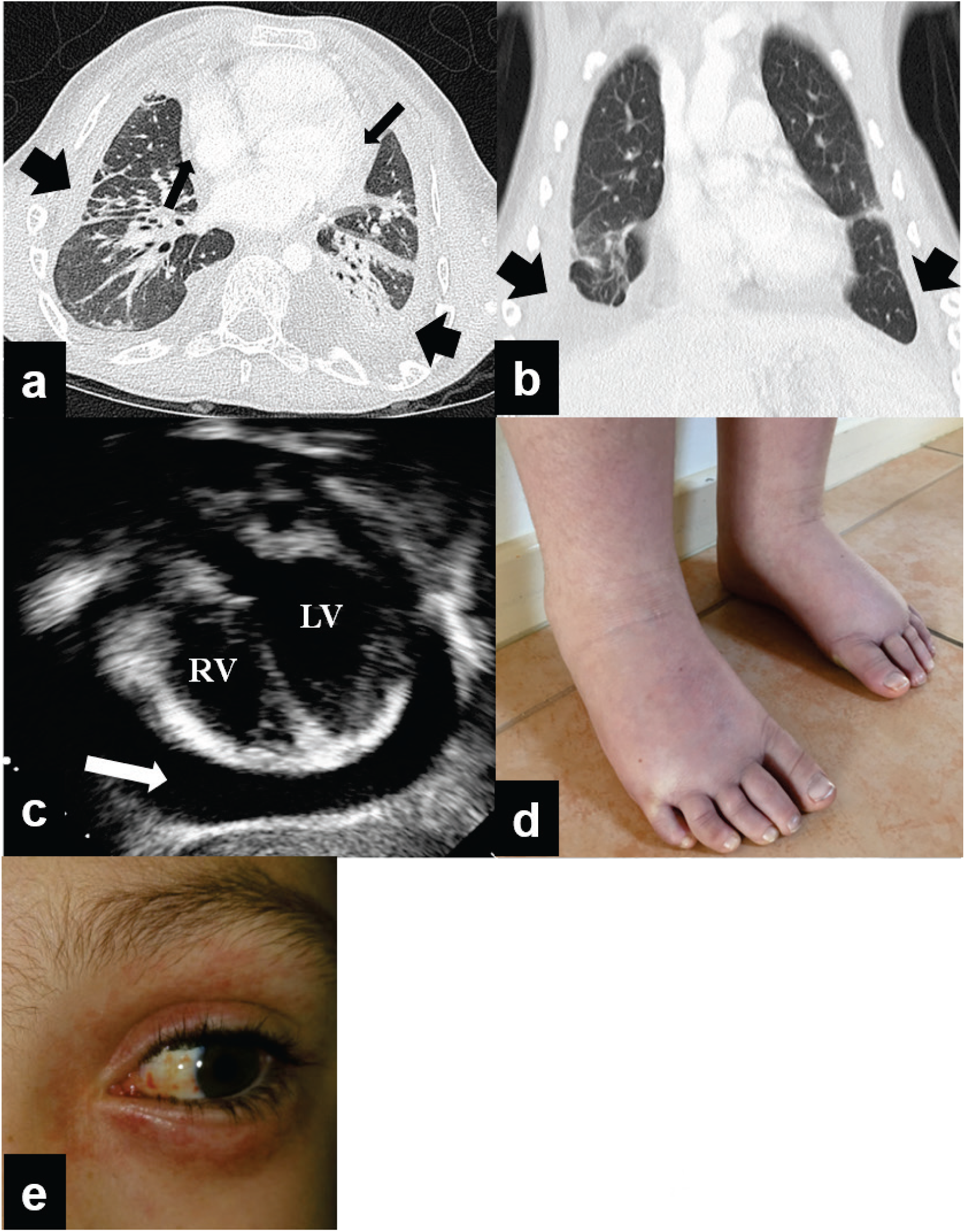
Clinical images of the GLD patients. (**a**) – (**c**) Proband (GLD08 II.1). In (**a**) and (**b**), computerised tomography (CT) of thorax demonstrating bilateral chylothorax (thick arrows) and a pericardial effusion (thin arrows) and in (**c**) a large pericardial effusion (white arrow) requiring percutaneous drainage twice for recurrence. RV, right ventricle. LV, left ventricle. (d) – (**e**) Proband (GLD09 II.1). In (**d**) showing bilateral lower limb lymphoedema in early childhood and (**e**) bilateral periorbital and conjunctival vascular changes with small punctate haemorrhages.

**Figure S3:**
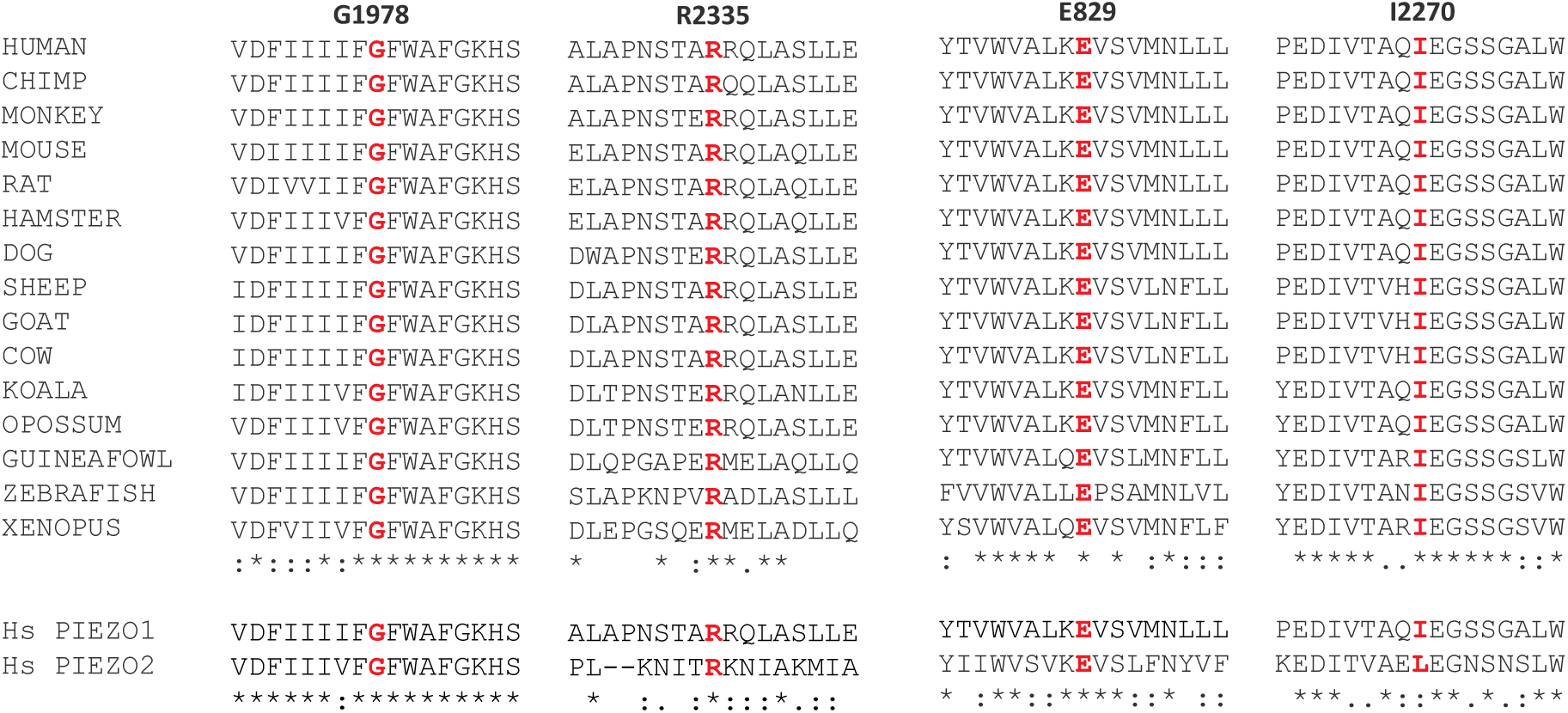
Amino acid sequence conservation at the affected sites. Upper panel: Single-letter amino acid code for sections of PIEZO1 amino acid sequences at and around the sites of the 4 GLD-associated variants after amino acid identity alignments of sequences of the 15 species indicated on the left. The wild-type residues are indicated across the top for the sites of the 4 variants in human PIEZO1 (hPIEZO1) and shown in red across the 15 species. Asterisks at the bottom indicate perfect sequence conservation across the species. Lower panel: Alignment results for human (Homo sapiens, Hs) PIEZO1 and PIEZO2.

**Figure S4:**
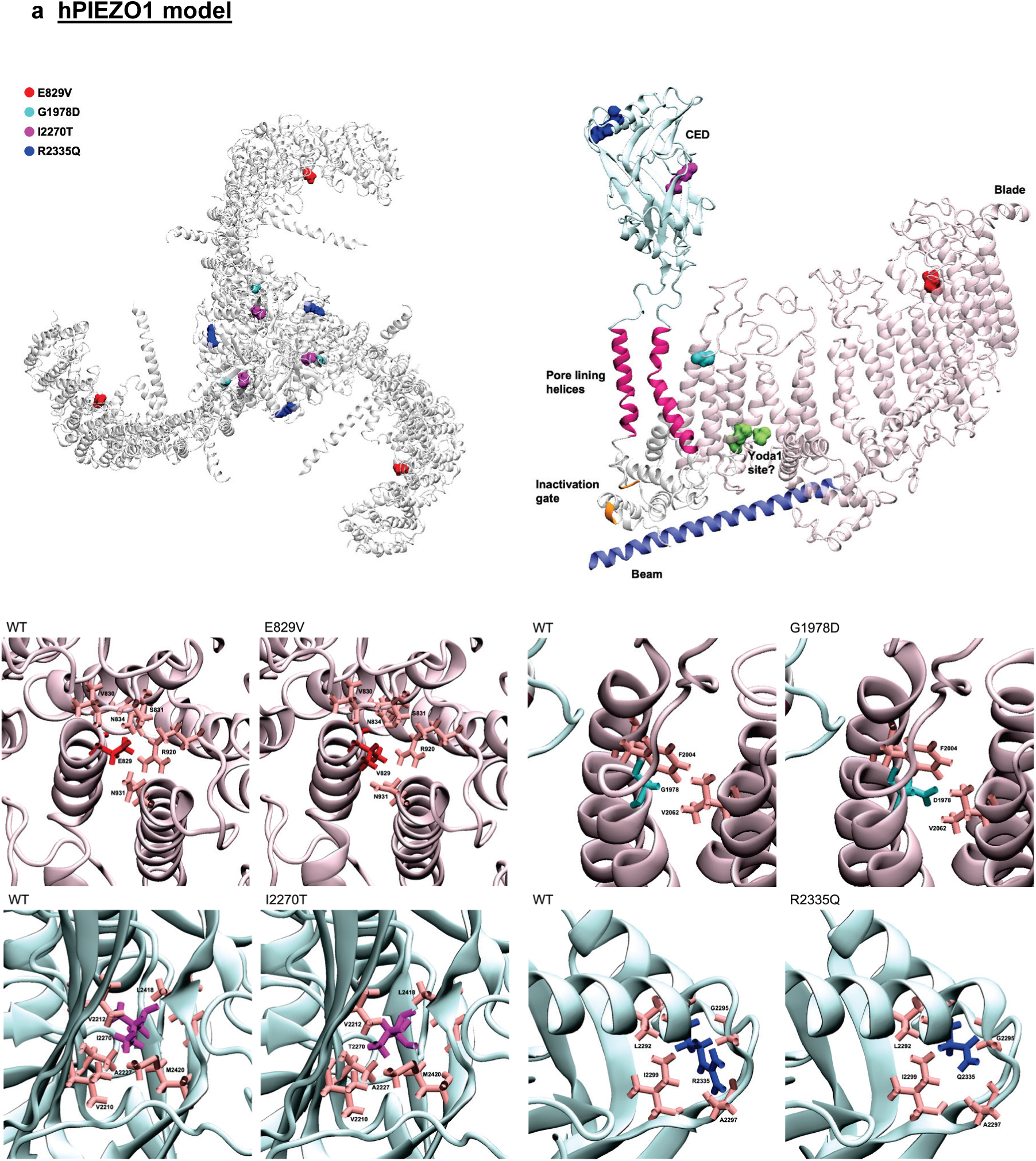
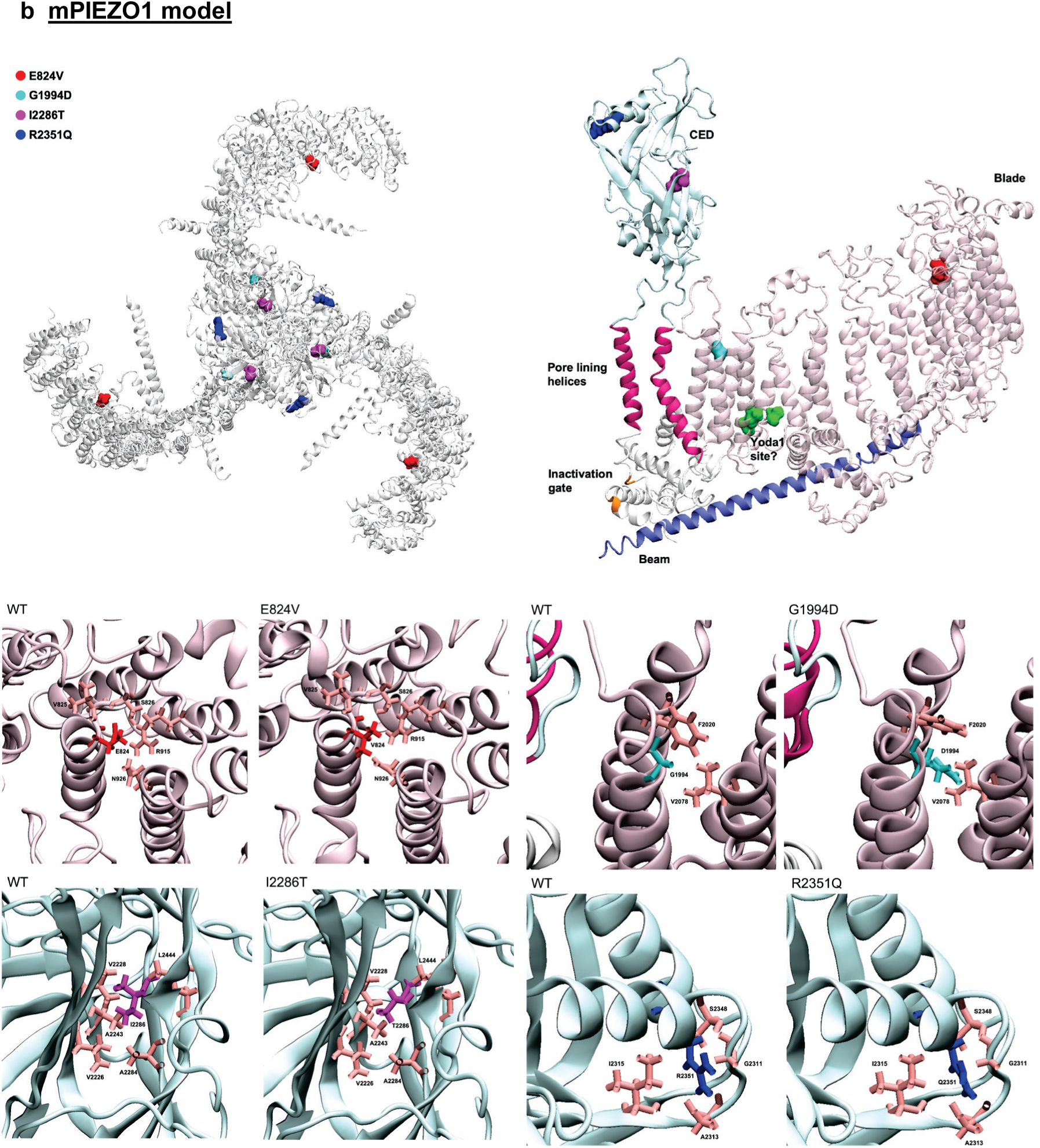
Molecular models of PIEZO1 channels. hPIEZO1 (**a**) and mPIEZO1 (**b**) models are shown using cartoon representation with mutated residues displayed as surface representation according to the colour-code indicated. Top left: trimer in helicopter view. Top right: one monomer in side view. Key features of the channel are indicated: C-terminal extracellular domain (CED, pale blue); pore lining helices (pink); inactivation gate (orange); beam (blue); blade (pale pink); and potential Yoda1 interaction amino acid residues (green) suggested by mutagenesis studies ^34^. Lower panels: close-ups of the channel at the mutated residues for variants and wildtype (WT) with neighbouring amino acid residues highlighted. Amino acid residues of interest are shown using dynamic bonds representation.

**Figure S5:**
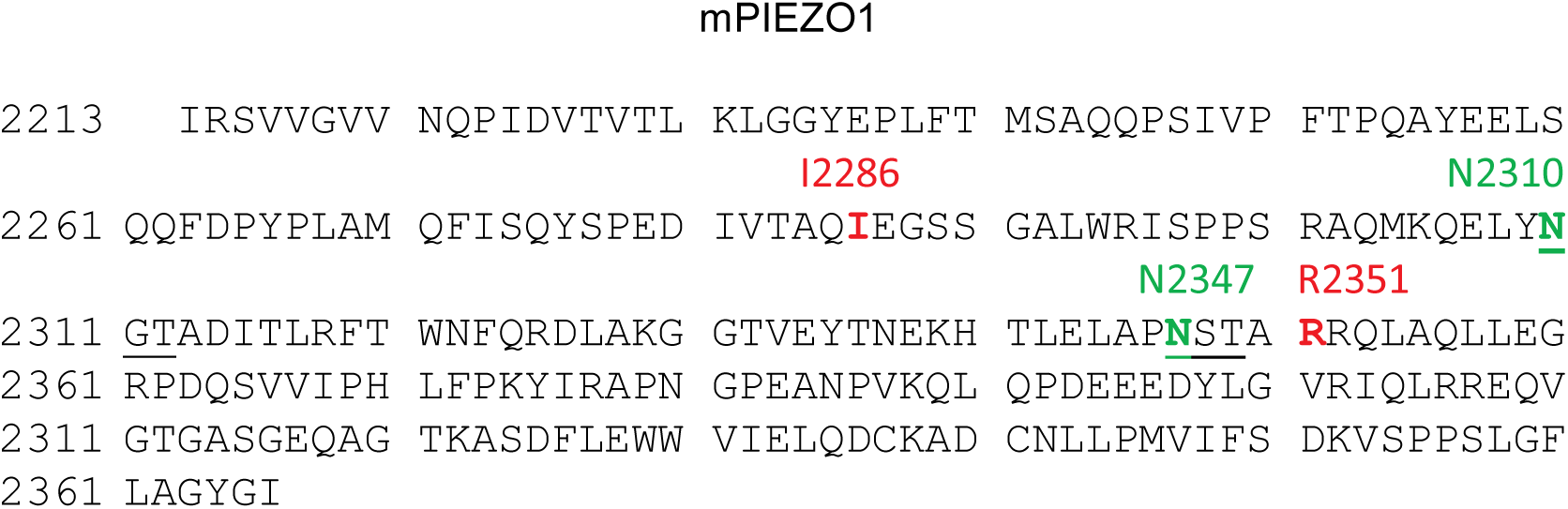
Proximity of two of the GLD-associated variants to N-linked glycosylation sites. Single-letter amino acid code with C-terminal extracellular domain (CED) of mPIEZO1 with predicted N-linked glycosylation sites in green and CED-located GLD-associated variants in red.

**Figure S6:**
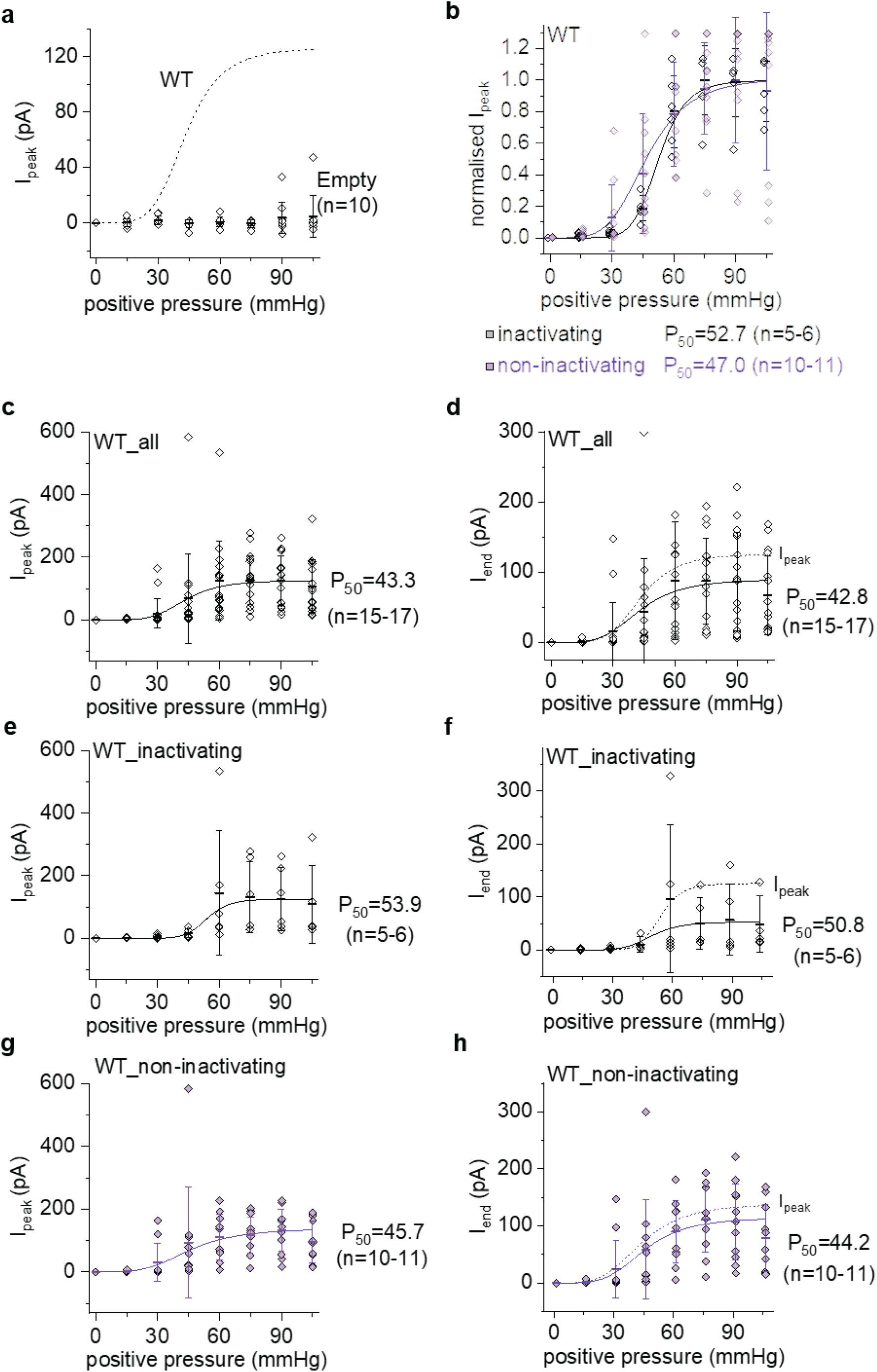
Additional data and analysis for Figure 3. Data for outside-out patch recordings from T-REx-293 cells transfected with empty vector or WT mPIEZO1 construct. The voltage across the patches during recordings was -80 mV. (a) As in Figure 3(d) but data for cells transfected with empty vector (n = 10). No Boltzmann function is fitted to these data because there is little or no channel activity. The WT Boltzmann curve from Figure 3(d) is superimposed for comparison as a dashed line. (b) As in Figure 3(d) but the WT data are split into inactivating kinetic and non-inactivating groups. (c) As in Figure 3(d) but with the data shown as absolute current (i.e., without normalisation). (d) As for (**c**) but current measured at the end of the 200-ms pressure step (I_end_). The Boltzmann curve from (**c**) is superimposed for comparison as a dashed line (I_peak_). (**e**, **f**) As for (**c**, **d**) but inactivating currents only. (**g**, **h**) As for (**c**, **d**) but non-inactivating currents only.

**Figure S7:**
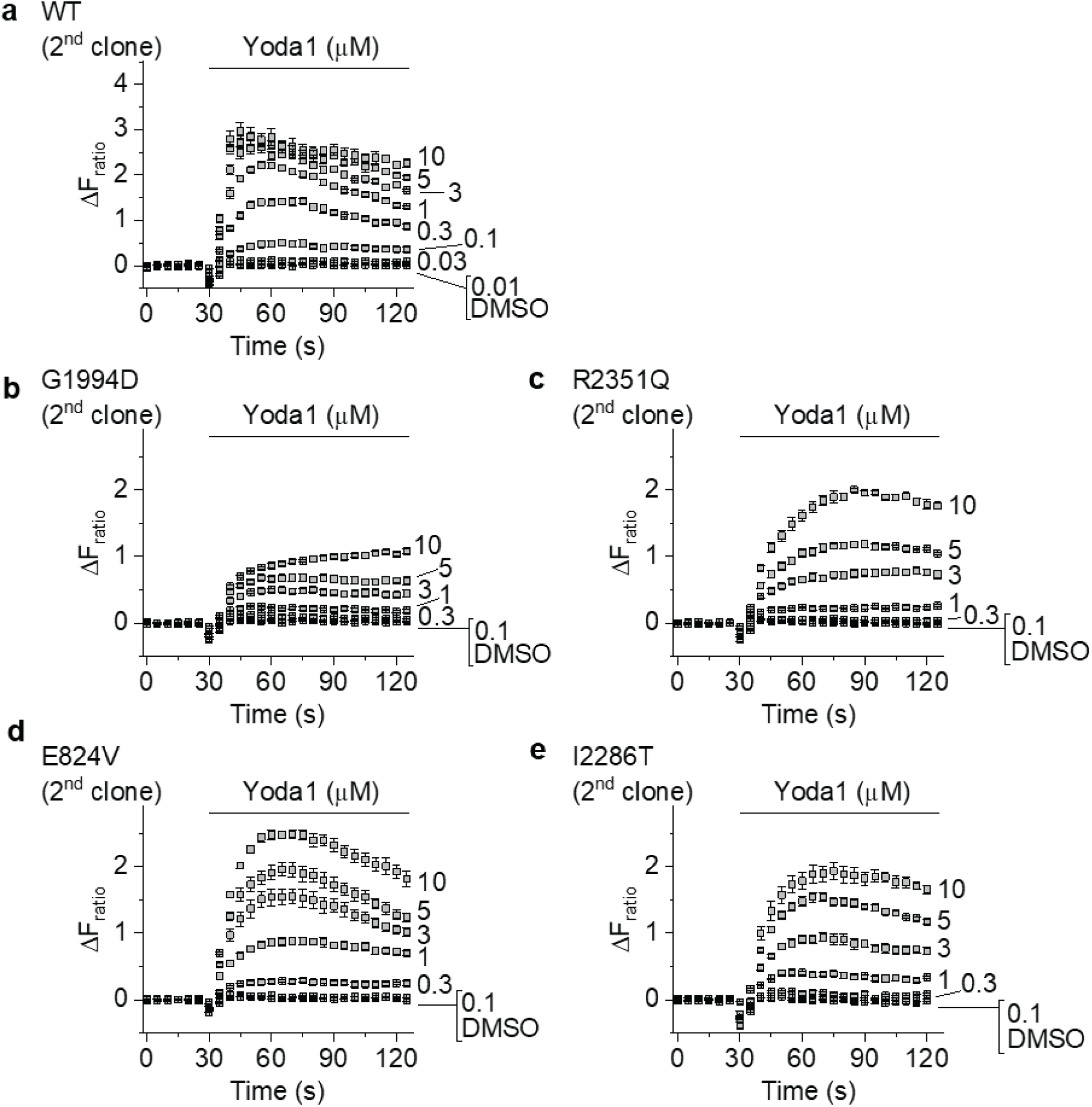
Further data in support of Figure 4. Experiments and data are comparable with those of Figure 4 (b), (d), (e), (f) and (g) but replicated in independent clonal cell lines for the same channels (2^nd^ clone).

**Figure S8:**
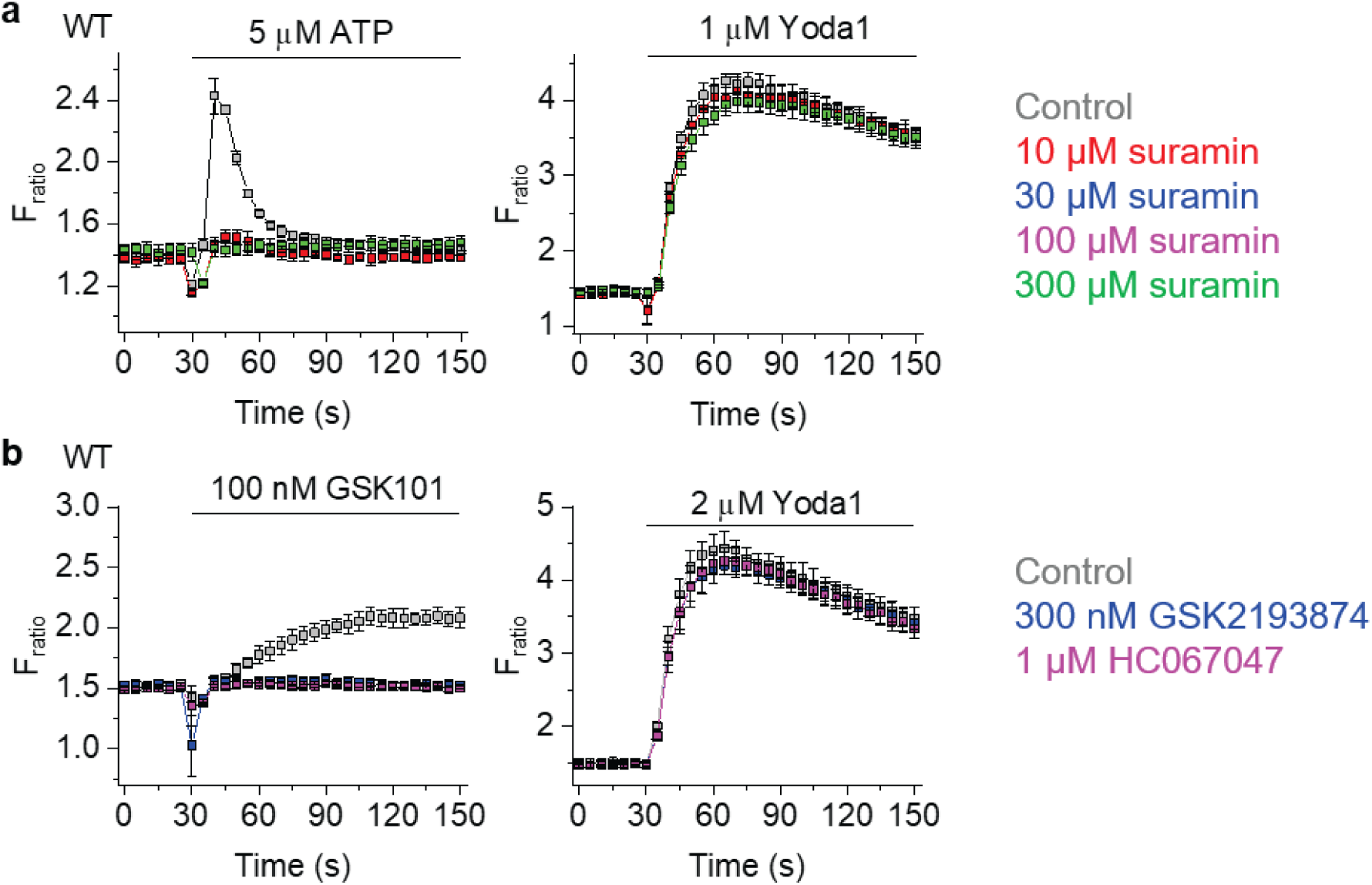
Lack of contribution from ATP receptors and TRPV4 channels. Data for Ca^2+^ measurements from T-REx-293 cells transfected with wild-type (WT) mPIEZO1. Data are displayed as the intracellular Ca^2+^ concentration indicated by absolute fura-2 fluorescence (F) ratio (F_ratio_), with increase in ratio being increase in Ca^2+^ concentration in the cytosol. Data are each from a single representative 96-well plate experiment with cells exposed to the indicated concentrations of ATP, Yoda1 or TRPV4 agonist GSK1016790A (GSK101) (mean ± s.e.m., N = 5 wells each) in the presence of: (**a**) P2Y2 antagonist (suramin) at the indicated concentrations or vehicle control; or (**b**) TRPV4 antagonists (GSK219384 or HC067047) at the indicated concentrations or vehicle control.

**Figure S9:**
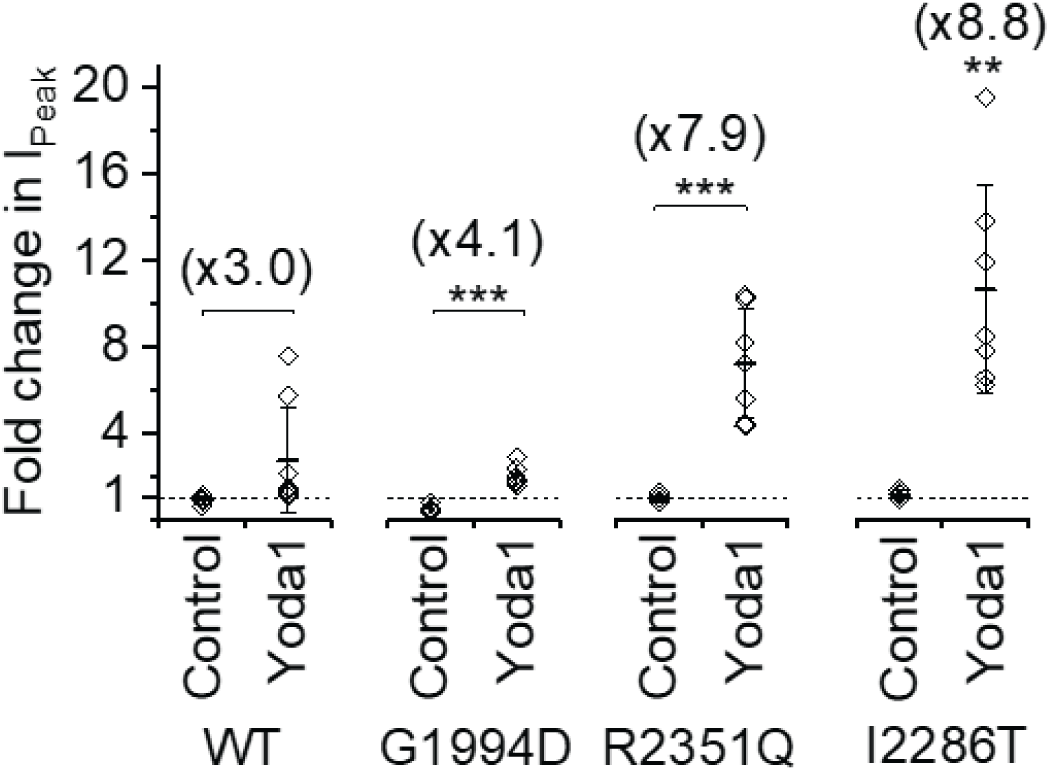
Further analysis of rescued mechanical activation by Yoda1 (supporting data of Figure 6) Analysis of data shown from Figure 6a, b and d, showing the effect of Yoda1 as a fold-change (increase) in inward current amplitude during the pressure step relative to the current before Yoda1 was applied. Data are shown as mean ± s.d. fold-change with each independent data point superimposed (n = 5-7). **P<0.01 and ***P<0.001. The mean fold-change is indicated numerically above each data set (e.g., x3.0 indicates 3.0-fold increase evoked by Yoda1).

**Figure S10:**
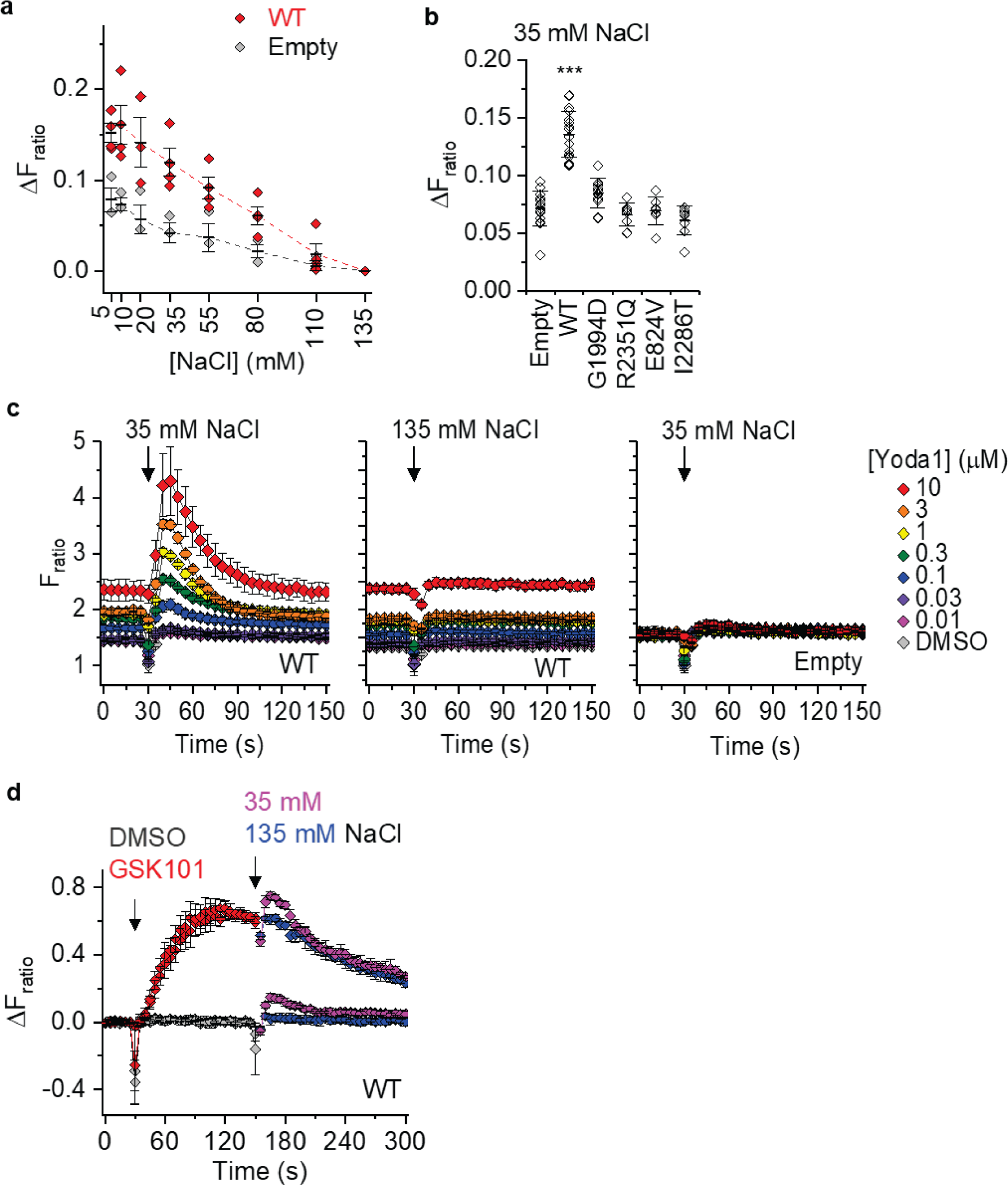
Further data for the effect of hypo-osmolality (supporting data for Figure 7) (a) Summary mean ± s.d. data (n = 3-4) from experiments of the type shown in Figure 7a, measured 10-40 s post-injection (indicated by arrows in Figure 7a). (b) Summarised mean ± s.d. data and individual data points superimposed comparing the responses to hypotonic challenge (35 mM NaCl) for empty vector (Empty) or wild-type (WT) or variant mPIEZO1 constructs (n = 8-13). Only WT channels showed a statistically significant increase compared with Empty (***P<0.001, one-way Anova with Bonferroni post-hoc test). (c) Typical 96-well plate data for changes in intracellular Ca^2+^ evoked by hypotonic (35 mM NaCl) or normotonic (135 mM NaCl) medium for WT- or Empty vector-transfected cells in increasing concentrations of Yoda1 as indicated and colour-coded on the right. Summary mean data for all experiments of this type are in Figure 7d. (d) For cells expressing WT mPIEZO1, responses to the TRPV4 agonist GSK1016790A (GSK101, 100 nM) or its vehicle control (DMSO) prior to challenge with hypotonic (35 mM NaCl) or normotonic (135 mM NaCl) buffer (mean ± s.e.m., N = 4-5 wells each). The response to hypo-osmolality is similar with and without GSK101, suggesting GSK101 does not mimic the action of Yoda1 and that elevation of intracellular Ca^2+^ itself (i.e., caused by TRPV4 activation in this case) is not sufficient to enhance the hypo-osmolality response.

**Figure S11:**
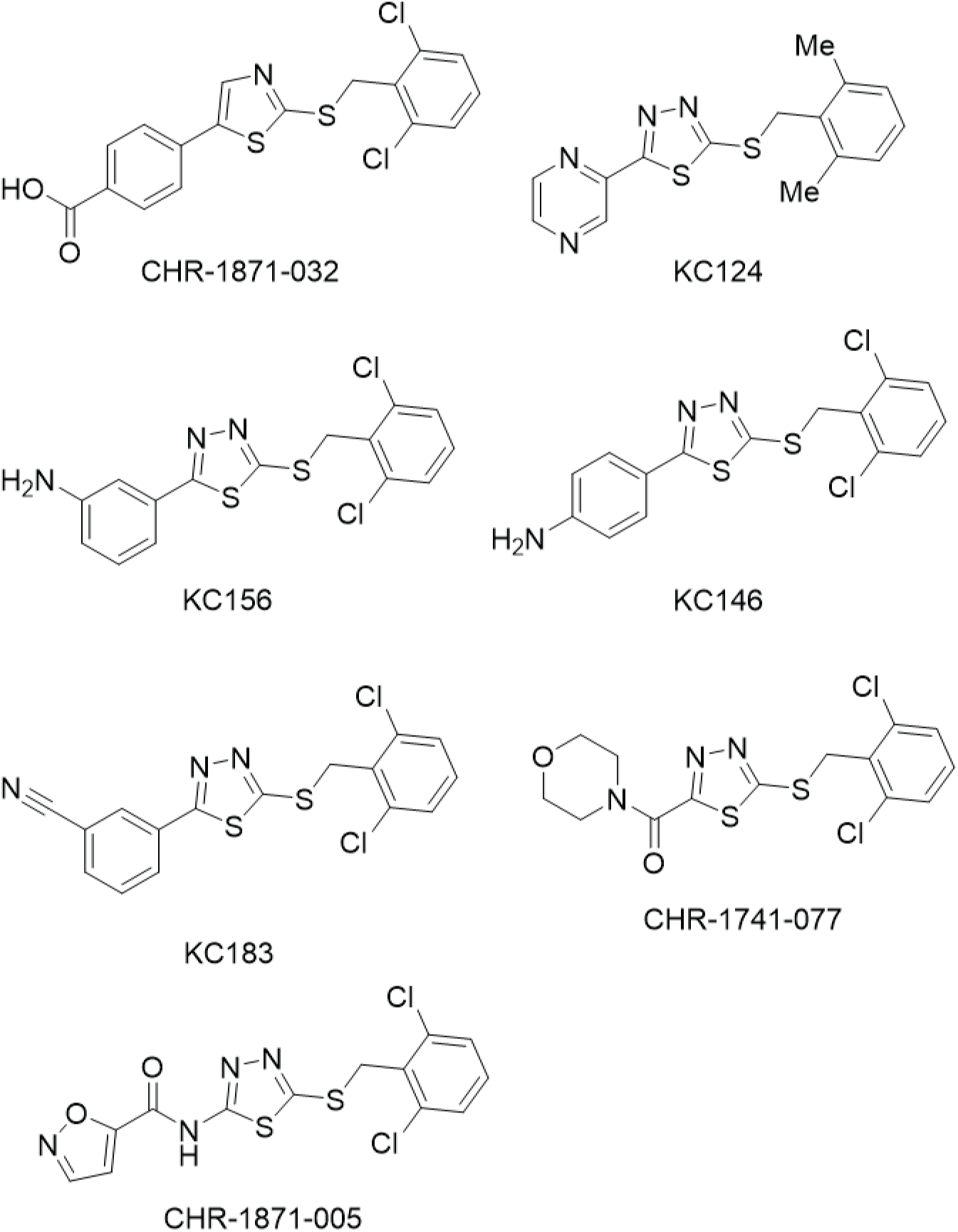
Chemical structures of the novel Yoda1 analogues.

**Figure S12:**
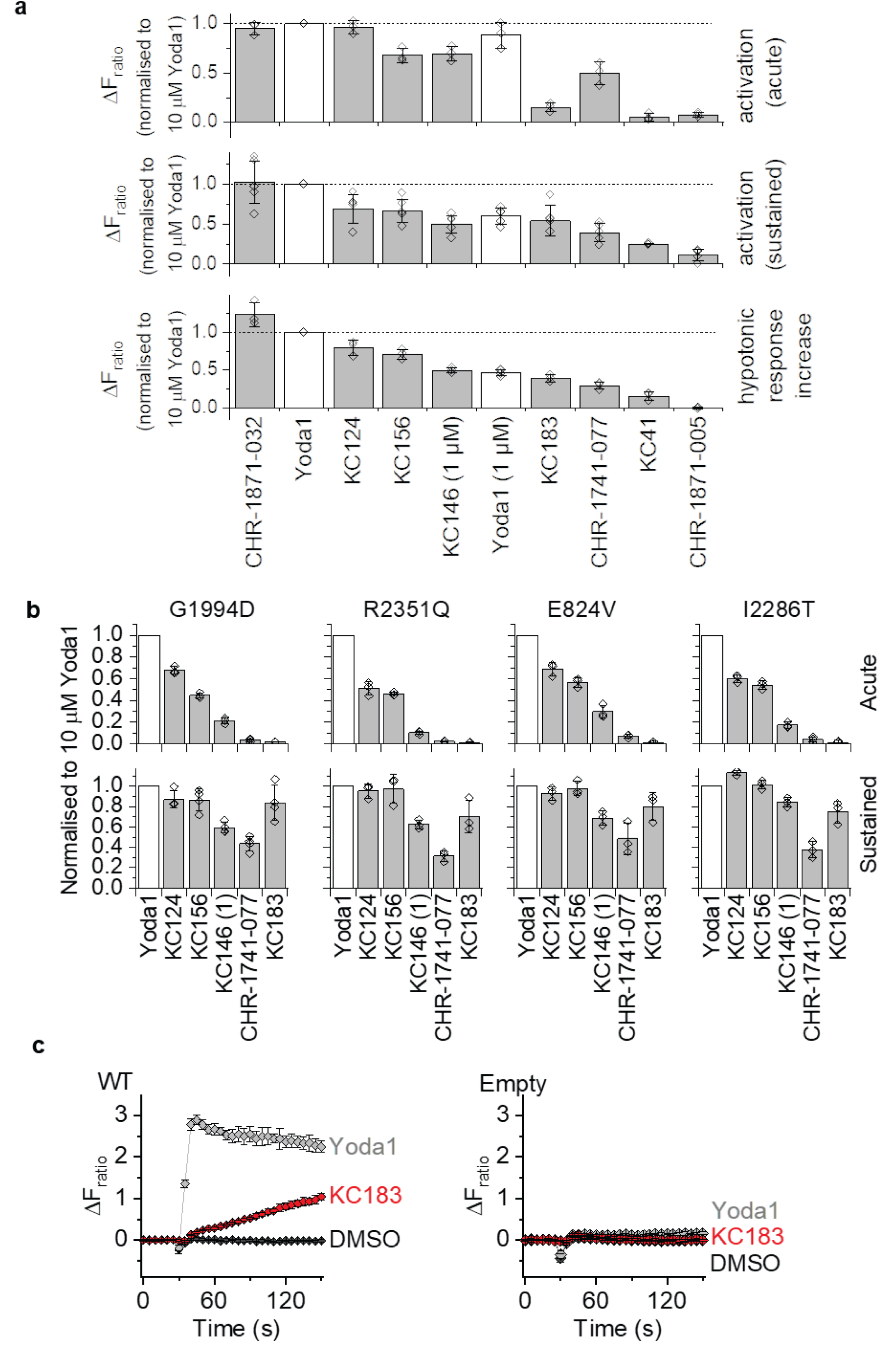
Yoda1 analogue screening data and further data for the effects of Yoda1 analogues. (a) Ca^2+^ measurement data for T-REx-293 cells transfected with wild-type (WT) mPIEZO1 comparing the effects of 7 Yoda1 analogues with Yoda1 at 10 μM concentration unless otherwise stated as 1 μM. Measurements are shown for elevation of Ca^2+^ (ΔF_ratio_) at 30-60 s (Acute activation) and ≥ 20 min (Sustained activation) after injecting the indicated compound (analogue) and amplification of the response to hypo-osmolality (35 mM NaCl). Compounds are ranked (left-to-right) according to their ability to amplify the hypo-osmolality response. Data are plotted as mean ± s.d. (n = 3-8). All effects are normalised to those of 10 μM Yoda1, as emphasized by the horizontal dashed lines at ΔF_ratio_ = 1.0. (b) As in (**a**) but for the indicated variant channels and acute and sustained elevation of Ca^2+^ only and 5 of the Yoda1 analogues as indicated. All analogues were tested at 10 μM except for KC146, which was tested at 1 μM because it caused a fluorescence artefact at 10 μM. (c) Example Ca^2+^ time-series data for KC183 compared with Yoda1 at 10 μM at WT channels and in empty vector-transfected cells (mean ± s.e.m., N = 4-5 wells each).

**Figure S13:**
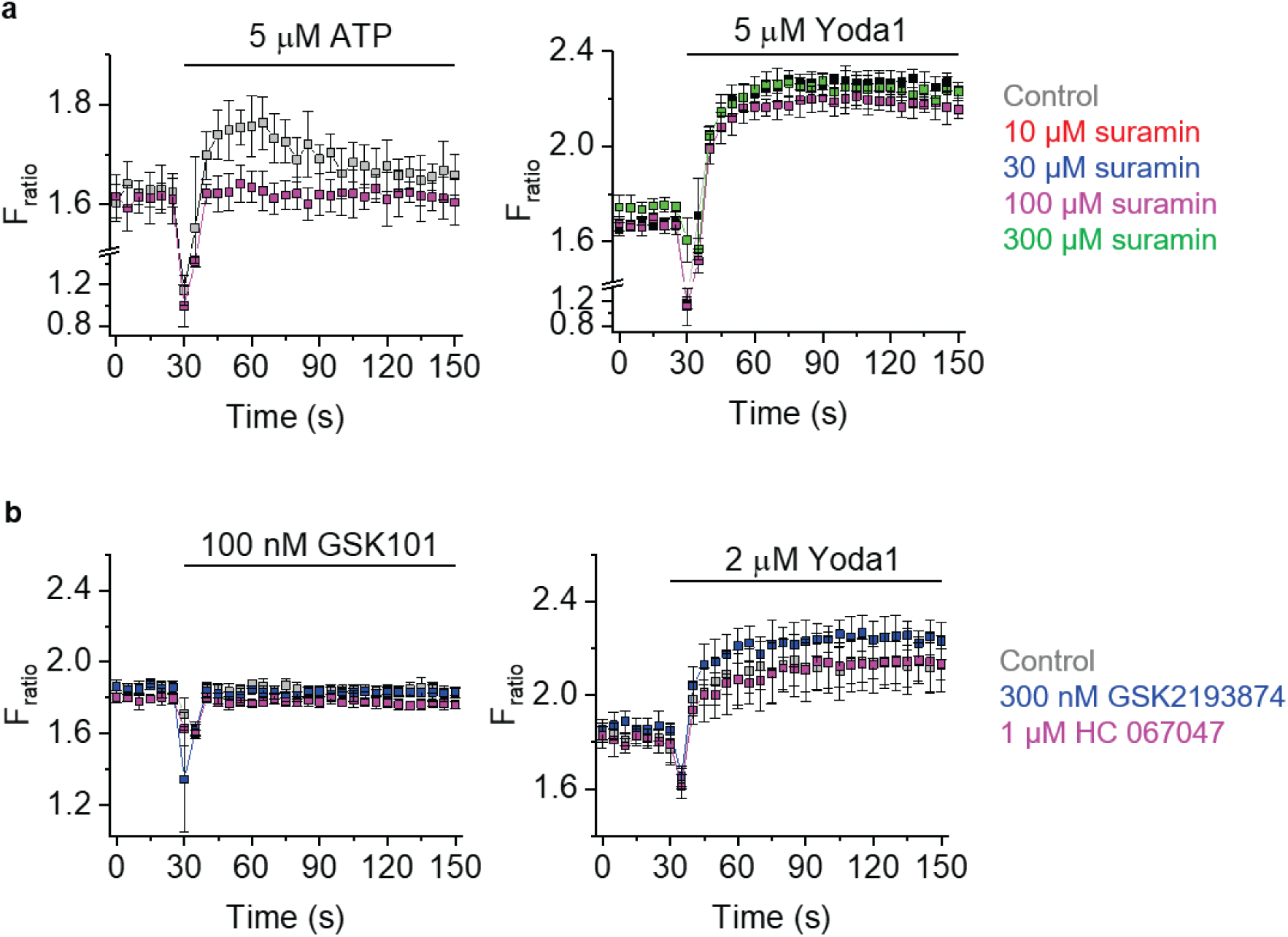
Contributions of ATP-release and TRPV4 channels in SVEC4-10 cells. Ca^2+^ measurement data for the endothelial cell line derived from lymph nodes, SVEC4-10 cells. Data are displayed as the intracellular Ca^2+^ concentration indicated by fura-2 fluorescence (F) ratio (F_ratio_), with increase in ratio indicating increase in Ca^2+^ concentration in the cytosol. Data shown are from single representative 96-well plate experiments showing effects of the indicated concentrations of ATP, Yoda1 and GSK1016790A (GSK101) (mean ± s.e.m., N = 5 wells each) in the presence of: (**a**) Non-selective P2Y2 antagonist (suramin) at the indicated concentrations or vehicle control; or (**b**) TRPV4 antagonists (GSK219384 or HC067047) at the indicated concentrations or vehicle control.

### SI FURTHER CLINICAL INFORMATION ON THE GLD PATIENTS

#### GLD07

Proband (GLD07 II.1) is a male in his teens born to non-consanguineous parents. First trimester nuchal translucency was normal. Antenatal imaging (gestational date not noted) identified NIFH and polyhydramnios. The baby was hydropic at birth, which resolved spontaneously. However, over a period of 6 – 12 months, he developed symmetrical lower limb lymphoedema with intermittent upper limb, facial (eyelid) and genital (scrotal) lymphoedema. At the age of 1-2 years his peripheral oedema had stabilised mainly affecting the feet, with persistent pleural effusions and minimal perihepatic ascites, diagnosed on MR imaging. Apart from a maldescended left testis, he has no other structural malformations. He has micrognathia and slight facial swelling but is otherwise non-dysmorphic. There are no concerns about his neurodevelopment. At the age of 3-4 years he developed cellulitis (followed by rheumatic fever) which was probably lymphoedema-related and he has been on penicillin prophylaxis since.

#### GLD08

The proband (GLD08 II.1) is a male in his 30s, born to consanguineous parents. His antenatal and neonatal period was uneventful. He was born at term with a normal birth weight. From age 10-14 years he developed progressive breathlessness and reduced exercise tolerance and was found to have bilateral chylous pleural effusions (triglycerides 19.42mmol/L with moderate lymphocytosis). Treatment has proved very difficult and has involved pleurodesis, ligation of the thoracic duct and pleuro-peritoneal shunting. He was also found to have a hydrocele (likely due to a patent ductus vaginalis) and ascites. At 15-19 years old he developed a pericardial effusion that rapidly progressed to cardiac tamponade. Pericardiocentesis confirmed chylous pericardial fluid, and this was repeated less than 3 weeks later due to recurrence of the pericardial effusion and tamponade (SI Figure S2). He was subsequently treated with octreotide and medium chain triglyceride diet for two years. He has residual, significant and symptomatic restrictive lung disease requiring nocturnal non-invasive ventilation. He developed bilateral lower limb lymphoedema with scrotal oedema in his 30s. Additional clinical features include mild microcephaly, short stature, and generalised osteopenia. He has a normal male karyotype and a 12-gene Ras-MAPK pathway disorders panel identified no causal variants. Lymphoscintigraphy has not been performed.

GLD08 I.2 is the proband’s father. His parents are not known to be consanguineous. He was diagnosed with spontaneous bilateral chylous effusions in his 30s. Despite pleurodesis, his effusions are persistent resulting in a restrictive lung defect and nocturnal hypoventilation requiring non-invasive ventilation (NIV). He also has a mild-moderate pericardial effusion, which has been stable and has not required pericardiocentesis. He has bilateral lower limb pitting oedema to the knees (onset at the age of 45-49 years) and varicose veins with varicose eczema at the ankles. He has no further complications and is developmentally normal. Venous duplex and lymphoscintigraphy not performed.

Recently, proband’s brother, GLD08 II.2, was found to have bilateral pleural effusions at the age of 10-14 years. A left drain was placed and chyle confirmed (triglycerides 21 mmol/L in pleural fluid). He also presented with mild peripheral oedema of the lower limbs, and tense bilateral hydrocoeles.

Proband’s maternal aunt (GLD08 I.1) had progressive pleural and pericardial effusions (with pleural and pericardial thickening) that first started aged 15-19 years. She died from chylothoraces and pleural disease in her 30s and was not formally assessed in clinic.

#### GLD09

GLD09 II.1 is the first child of non-consanguineous Caucasian parents, conceived by *in vitro* fertilisation. The first trimester nuchal translucency measurement was normal. At 19- weeks’ gestation the baby was noted to have bilateral hydrothoraces. Amniocentesis at this time revealed a 46,XY karyotype. The pregnancy was complicated by polyhydramnios requiring amnio-drainage thrice during early third trimester at 3-weekly intervals. A male infant was born at 34-weeks’ gestation with significant facial swelling and bilateral chylothoraces requiring bilateral chest drains. He was intubated and ventilated for one month and remained in neonatal intensive care for a total of three months. During this period, he had laryngomalacia and severe gastro-oesophageal reflux that required a fundoplication at 6-12 months of age. A gastrostomy was inserted owing to an oral aversion, with a low fat MCT diet instituted in the setting of the bilateral chylothoraces. A lymphoscintigram at 6-12 months demonstrated extensive subdermal flow of lymphatics in the lower limbs, chest wall and scrotum. Upon review at 1-2 years of age, he had oedema of the lower limbs to the thighs, and genitalia, as well as intermittent facial swelling (SI Figure S2). He had metopic synostosis and a facial haemangioma over the glabella. He had a circumcision at 1-2 years of age for significant scrotal/genital oedema. By 5-9 years of age he had contracted pneumonia on three occasions and was hospitalised with invasive group A Streptococcal septicaemia. He also began to exhibit bilateral periorbital and conjunctival vascular changes with small punctate haemorrhages. He had obstructive sleep apnoea from age 5-9 years treated with nocturnal CPAP. Orthodontic treatment with an upper jaw expander has resulted in clinical improvement of sleep apnoea symptoms. Early concerns regarding speech and language delay were resolved by the commencement of primary school. He was diagnosed with Asperger syndrome at 3-4 years of age. There is a bilineal family history of autism spectrum disorder.

GLD09 I.1 is the proband’s father. He has four limb lymphoedema, with swelling of the lower limbs at birth, followed by swelling of the hands from his 20s. The cause of this is unknown. He has had no episodes of cellulitis and no history of pleural effusions. He also has a diagnosis of Asperger syndrome. He only has one variant in *PIEZO1* [c.2486A>T; p.(E829V)].

### SI CHEMISTRY

**Scheme 1.**
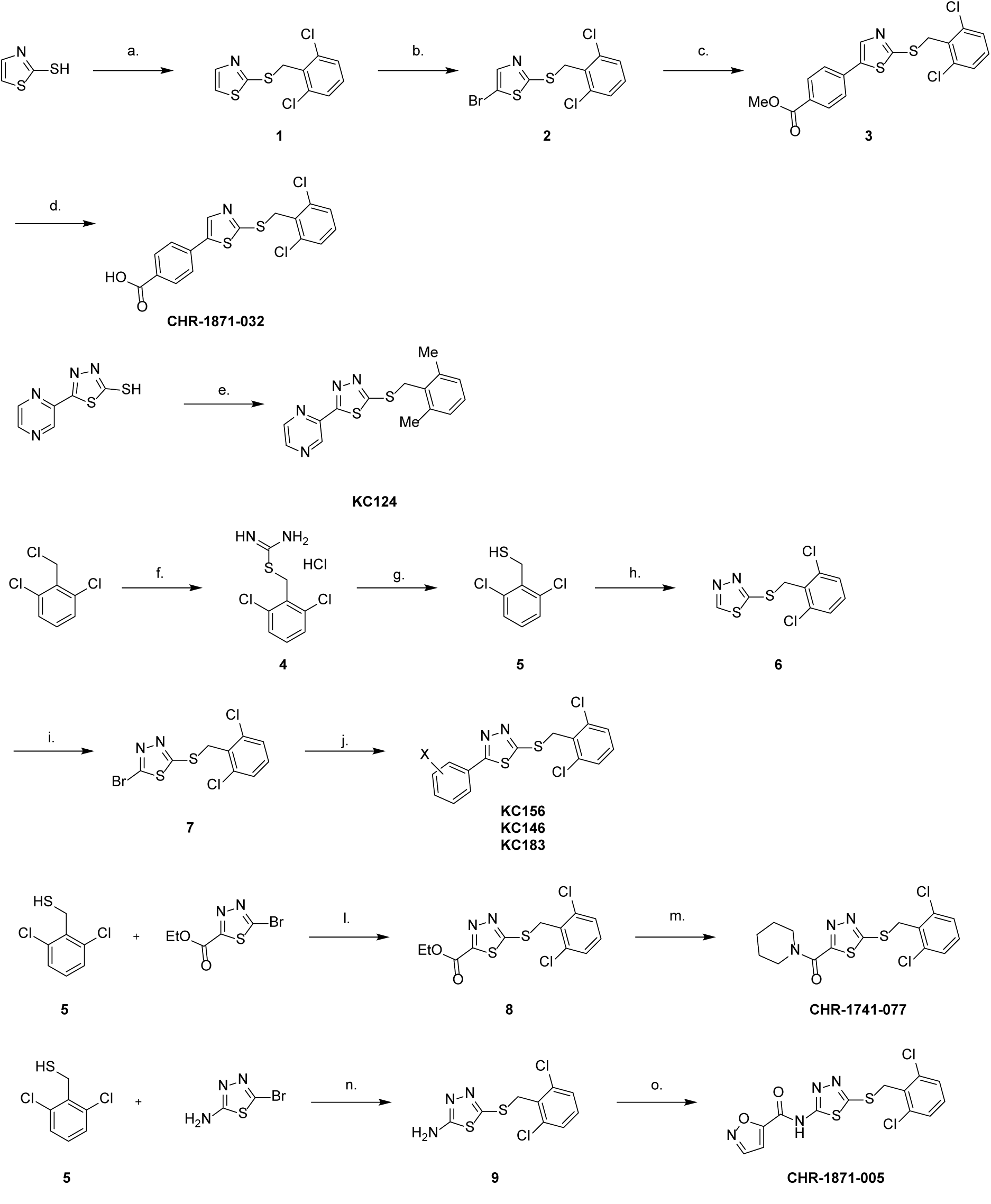
Reagents. a. 2,6-dichlorobenzyl chloride, K_2_CO_3_, DMF, 90 °C, N_2_, 1h, 83% b. NBS, DMF, 3h, 73% c. 4-methoxycarbonylphenyl boronic acid, 2M Na_2_CO_3 (aq)_, Pd(PPh_3_)_4_, 1,4- dioxane, 90 °C, N_2_, 18h, 38% d. 10M NaOH _(aq)_, THF, RT-60 °C, 16.5 h, 69% e. 2,6-dimethylbenzyl chloride, KOH, DMF, RT-80 °C, N_2_, 2h, 23% f. Thiourea, EtOH, reflux, 45 min, 99% g. 2M NaOH **_(aq_**_)_, EtOH, reflux, N_2_, 5h, then 1M HCl, RT, N_2_, 18 h, 71% h. 2-bromo-1,3,4- thiadiazole, K_2_CO_3_, DMF, 90 °C, N_2_, 18 h, 57% i. NBS, DCM, reflux, 48h, 78% j. Appropriate phenyl boronic acid, K_2_CO_3_, Pd(PPh_3_)_4_, 1,4-dioxane, H_2_O, 90 °C, N_2_, 2-24h, 18-25% k. Et_3_N, DMF, 90 °C, N_2_, 90°C, 21h, 80% l. piperidine, EtOH, reflux, N_2_, 21h, 39% m. Et_3_N, DMF, 90°C, N_2_, 20h, 74% n. isoxazole-5-carbonyl chloride, Et_3_N, DMAP, THF, RT, 20 h, 67%

#### General Procedure A

The desired aromatic halide (1.0 eq.) the desired boronic acid/ester (1-3.5 eq.) and K_2_CO_3_ (4.0 eq.) were dissolved in anhydrous 1,4-dioxane (2 mL) and H_2_O (2 mL) then degassed with N_2_ for 30 minutes. Pd(PPh_3_)_4_ (0.15 eq.) was then added and the reaction was then heated to 90 °C for 2-24 h. Upon completion, the reaction was diluted with H_2_O (20 mL), extracted with DCM (3 × 10 mL), dried over Na_2_SO_3_, filtered and reduced in vacuo to afford the crude or pure product.

2-([(2,6-dichlorophenyl)methyl]sulfanyl)-1,3-thiazole (**1**)

To a solution of 1,3-thiazole-2-thiol (134 mg, 1.14 mmol) and K_2_CO_3_ (170 mg, 1.05 mmol) in DMF (4 mL) under N_2_ was added 2,6-dichlorobenzyl chloride (240 mg, 1.05 mmol) and the reaction heated to 90 °C. After 1h the reaction was cooled to RT and diluted with H_2_O (70 mL). The aqueous suspension was extracted with EtOAc (3 x 70 mL) and the combined organic phases were washed with brine (50 mL) and 10% LiCl solution (30 mL), dried (Na_2_SO_4_) and concentrated *in vacuo*. Crude mixture purified by ACC (0-10% EtOAc in Petroleum ether (40- 60 °C)) to isolate the compound as a colourless liquid (260 mg, 0.94mmol, 83%). Rf 0.31 (19:1 Petroleum ether (40-60 °C):EtOAc (v/v)); δ_H_ (400 MHz, CDCl_3_): 7.76 (1H, d, thiazole H-4, *J* = 3.4 Hz), 7.31-7.29 (3H, m, thiazole H-5 and benzyl H-3/5), 7.16 (1H, ap. t, benzyl H-4, *J* = 6.6 and 8.4 Hz) 4.75 (2H, s, benzylic CH_2_); δ_C_ (100 MHz, CDCl_3_); 162.7 (thiazole C-2), 143.2 (thiazole C-4), 136.1 (benzyl C-2/6), 132.6 (benzyl C-1), 129.4 (benzyl C-4), 128.4 (benzyl C- 3/5) 120.6 (thiazole C-5), 35.4 (benzylic CH_2_); m/z ES+ Found MH^+^ 275.9462, C_10_H_8_Cl_2_NS_2_ requires MH^+^ 275.9469

5-Bromo-2-([(2,6-dichlorophenyl)methyl]sulfanyl)-1,3-thiazole (**2**)

To a solution of 2-([(2,6-dichlorophenyl)methyl]sulfanyl)-1,3-thiazole (**1**) (100 mg, 0.36 mmol) in DMF (3 mL) was added *N*-bromosuccinimide (97 mg, 0.54 mmol) and the reaction stirred at RT. After 3h the reaction was diluted with H_2_O (50 mL) and the aqueous solution extracted with EtOAc (3 x 30 mL). The combined organic phases were washed with brine (50 mL) and 10% LiCl solution (50 mL), dried (Na_2_SO_4_) and concentrated *in vacuo*. The crude product was purified by ACC (0-10% EtOAc in Petroleum ether (40-60 °C)) to isolate the compound as a colourless liquid (93 mg, 0.26mmol, 73%). Rf 0.43 (24:1 Petroleum ether (40-60 °C):EtOAc (v/v)); δ_H_ (400 MHz, CDCl_3_): 7.55 (1H, s, thiazole H-4), 7.24 (2H, d, benzyl H-3/5, *J* = 8.1Hz), 7.11-7.07 (1H, m, benzyl H-4), 4.62 (2H, s, benzylic CH_2_); δ_C_ (100 MHz, CDCl_3_); 163.6 (thiazole C-2), 144.4 (thiazole C-4), 136.0 (benzyl C-2/6), 132.4 (benzyl C-1), 129.5 (benzyl C-4), 128.5 (benzyl C-3/5), 109.0 (thiazole C-5), 35.4 (benzylic CH_2_); LC-MS m/z ES+ Found MH+ 355.78

Methyl 4-(2-([(2,6-dichlorophenyl)methyl]sulfanyl)-1,3-thiazol-5-yl)benzoate (**3**)

A solution of 5-bromo-2-([(2,6-dichlorophenyl)methyl]sulfanyl)-1,3-thiazole (**2**) (93 mg, 0.26 mmol), 4-methoxycarbonylphenyl boronic acid (57 mg, 0.31 mmol) and 2M Na_2_CO_3_ (0.39 mL, 0.79 mmol) in 1,4-dioxane (4 mL) under N_2_ was degassed for 20 min and Pd(PPh_3_)_4_ (15 mg, 0.01 mmol) added. The reaction was heated to 90 °C for 18 h, then cooled to RT and diluted with H_2_O (50 mL). The aqueous solution was extracted with EtOAc (2 x 40 mL) and the combined organic phases washed with brine (50 mL), dried (Na_2_CO_3_) and concentrated *in vacuo*. The crude product was purified using ACC (0-10% EtOAc in Petroleum ether (40-60 °C), then 0-25% EtOAc in Petroleum ether (40-60 °C)) to isolate the desired compound as a yellow solid (41 mg, 0.10 mmol, 38%). Rf 0.45 (4:1 Petroleum ether (40-60 °C):EtOAc (v/v)); δ_H_ (400 MHz, CDCl_3_): 7.98 (2H, d, C*H*CCOOMe, *J* = 8.4 Hz), 7.91 (1H, s, thiazole H-4), 7.49 (2H, d, C*H*C-thiazole, *J* = 8.4 Hz), 7.25 (2H, d, benzyl H-3/5, *J* = 8.1 Hz), 7.12-7.09 (1H, m, benzyl H-4), 4.72 (2H, s, benzylic CH_2_), 3.86 (3H, s, CO_2_*Me*); δ_C_ (100 MHz, CDCl_3_); 166.4 (*C*O_2_Me), 163.5 (thiazole C-2), 139.6 (thiazole C-5), 139.3 (thiazole C-4), 136.1 (benzyl C-2/6), 135.4 (C-thiazole), 132.4 (benzyl C-1), 130.5 (*C*CO_2_Me), 129.6 (*C*HCCO_2_Me), 129.5 (benzyl C-4), 128.5 (benzyl C-3/5), 126.2 (*C*HC-thiazole), 52.2 (CO_2_*Me*) 35.2 (benzylic CH_2_); m/z ES+ Found MH^+^ 409.9832, C_18_H_14_Cl_2_NO_2_S_2_ requires MH^+^ 409.9837

4-(2-([(2,6-Dichlorophenyl)methyl]sulfanyl)-1,3-thiazol-5-yl)benzoic acid **CHR-1871-032**

To a solution of methyl 4-(2-([(2,6-dichlorophenyl)methyl]sulfanyl)-1,3-thiazol-5-yl)benzoate (**3**) (34 mg, 0.08 mmol) in THF (4 mL) was added 10 M NaOH aq. solution (0.04 mL, 0.41 mmol) and the reaction stirred at RT for 1h then heated to 60 °C for 10 h. Further 10 M NaOH aq. solution (0.04 mL, 0.41 mmol) added and the reaction heated for a further 8 h before stirring at RT for 2 days. Further 10 M NaOH aq. solution (0.04 mL, 0.41 mmol) added and the reaction heated at 60 °C for a further 6.5h. The reaction was then cooled to RT and the volatile solvents removed *in vacuo*. The crude mixture was diluted with H_2_O (20 mL) and extracted with EtOAc (3 x 20 mL). The aqueous solution was acidified with 1 M HCl to pH1 and the resulting precipitate collected by filtration isolating 4-(2-([(2,6-dichlorophenyl)methyl]sulfanyl)- 1,3-thiazol-5-yl)benzoic acid as a pale yellow solid (22 mg, 0.06 mmol, 69%). δ_H_ (500 MHz, d_6_-DMSO): 13.09 (1H, br. s, CO_2_H), 8.38 (1H, s, thiazole H-4), 7.99 (2H, d, *J* = 8.4 Hz, C*H*CCO_2_H), 7.77 (2H, d, *J* = 8.4 Hz, phenyl H-2/6), 7.55 (2H, d, *J* = 8.1 Hz, benzyl H-3/5), 7.42-7.39 (1H, m, benzyl H-4), 4.76 (2H, s, benzylic CH_2_); δ_C_ (125 MHz, d_6_-DMSO): 167.2 (thiazole C-2), 162.3 (CO_2_H), 141.1 (thiazole C-4), 139.5 (thiazole C-5), 135.5 (phenyl C-1), 134.9 (benzyl C-2/6), 132.4 (phenyl C-4), 131.1 (benzylic C-4), 130.7 (phenyl C-3/5), 129.3 (benzylic C-3/5), 126.7 (phenyl C-2/6), 35.2 (benzylic CH_2_); m/z ES+ Found MH^+^ 395.9677, C_17_H_12_Cl_2_NO_2_S_2_ requires MH^+^ 395.9681

2-((2,6-Dimethylbenzyl)thio)-5-(pyrazin-2-yl)-1,3,4-thiadiazole **KC124**

5-(2-Pyrazine)-1,3,4-thiadiazole-2-thiol (25 mg, 0.13 mmol) and KOH (8 mg, 0.15 mmol) were added to degassed DMF (5 mL) under a flow of nitrogen. This solution was stirred for 1 h followed by dropwise addition of 2,6-dimethylbenzyl chloride (22 mg, 0.14 mmol) in degassed DMF (2 mL) over 25 minutes The solution was heated to 80 ⁰C and allowed to stir for 2 h. The reaction mixture was then diluted with H_2_O (20 mL) and extracted with EtOAc (3 × 15 mL), the organic layers were combined and washed with sat. NH_4_Cl solution (2 × 15 mL), sat. NaHCO_3_ solution (2 × 15 mL), sat. NaCl solution (2 ×15 mL), 10% LiCl solution (w/w) (2 × 15 mL), dried over Na_2_SO_4_, filtered and evaporated to dryness *in vacuo*. This was purified by ACC (0-80% EtOAc in Petroleum ether (40-60 °C)) to afford a white powder (10 mg, 0.03 mmol, 23%). Rf 0.45 (7:3 Petroleum ether (40-60 °C):EtOAc (v/v)); δ_H_ (400 MHz, CDCl_3_): 9.48 (1H, d, J =1.5 Hz, pyrazinyl 3-H), 8.58 (1H, d, *J* = 2.5 Hz, pyrazinyl 6-H), 8.53-8.52 (1H, m, pyrazinyl 5-H), 7.06 (1H, dd, J =8.5 & 6.5 Hz, benzyl 4-H), 6.99 (2H, d, *J* = 7.5 Hz, benzyl 3-H), 4.70 (2H, s, benzyl CH_2_), 2.39 (6H, S, Me); δ_C_ (100 MHz, CDCl_3_); 169.1 (thiadiazole 5-C), 167.4 (thiadiazole 2-C), 145.7 (pyrazinyl 6-C), 144.8 (pyrazinyl 2-C), 144.2 (pyrazinyl 5-C), 142.4 (pyrazinyl 3-C), 137.9 (benzyl 4-C), 130.9 (benzyl 1-C), 128.5 (benzyl 3-C), 128.3 (benzyl 3- C), 33.6 (benzyl CH_2_), 19.7 (Me); m/z ES+ Found MH^+^ 337.0525, C_15_H_14_N_4_S_2_ requires MH^+^ 337.0558

2,6-dichlorophenyl)methyl carbamimidothioate hydrochloride (**4**)

A solution of 2,6-dichlorobenzyl chloride (921 mg, 4.71 mmol) and thiourea (362 mg, 4.76 mmol) in EtOH (15 mL) was heated to reflux for 45 min. The reaction mixture was cooled to RT and concentrated *in vacuo* to give the title compound as a white solid (1.27 g, 4.68 mmol, 99%). Used without further purification. Compound can be stored for several months. δ_H_ (400 MHz, d_6_-DMSO): 9.52 (4H, s, 2 x NH_2_), 7.58 (2H, d, benzyl H-3/5, *J* = 8.2 Hz,) 7.47-7.43 (1H, m, benzyl H-4), 4.70 (2H, s, benzylic CH_2_); δ_C_ (100 MHz, d_6_-DMSO): 169.7 (S*C*(NH_2_)NH_2_), 135.6 (benzyl C-2/6), 131.8 (benzyl C-1), 130.2 (benzyl C-4), 129.4 (benzyl C-3/5), 32.0 (benzylic CH_2_); m/z ES+ Found MH^+^ 234.9848, C_8_H_9_Cl_2_N_2_S requires MH^+^ 234.9858

2,6-Dichlorobenzyl thiol (**5**)

A solution of (2,6-dichlorophenyl)methyl carbamimidothioate hydrochloride (**4**) (797mg, 3.93 mmol) in EtOH (15 mL) under N_2_ was treated with 2 M aq. NaOH (5.9 mL, 11.8 mmol) and heated to reflux. After 5 h the reaction was cooled to RT and 1 M HCl (15.7 mL, 15.7 mmol) added. The reaction was stirred for 18h, then diluted with H_2_O(80 mL). The aqueous solution was extracted with EtOAc (3 x 70 mL) and the combined organic phases washed with brine (70 mL), dried over MgSO_4_ and concentrated *in vacuo* to give 2,6-dibenzyl thiol as a colourless oil which forms a white solid on standing (535 mg, 2.77 mmol, 71%. Compound must be used within weeks of synthesis. δ_H_ (400 MHz, CDCl_3_): 7.23 (2H, d, benzyl H-3/5, *J* = 8.04 Hz), 7.07-7.03 (1H, m, benzyl H-4), 3.92 (2H, d, benzyl CH_2_, *J* = 8.4 Hz), 2.02 (1H, t, SH, *J* = 8.4 Hz); δ_c_ (100 MHz, CDCl_3_): 137.3 (C-2/6), 134.6 (C-1), 129.0 (C-4), 128.4 (C-3/5), 24.4 (benzyl CH_2_)

2-((2,6-Dichlorobenzyl)thio)-1,3,4-thiadiazole (**6**)

2,6-dichlorobenzyl thiol (**5**) (2.85 g, 14.77 mmol), 2-bromo-1,3,4-thiadiazole (2.43 g, 14.77 mmol), & K_2_CO_3_ (2.38 g, 17.72 mmol) were dissolved in DMF (10 mL) and heated to 90 °C for 18 h. The reaction was diluted with H_2_O (100 mL), extracted with EtOAc (3 × 40 mL) and the organic layers combined. These were washed with brine (3 × 40 mL), 10% LiCl (3 × 40 mL), dried over MgSO_4_, filtered and concentrated *in vacuo* to give brown residue (4.33 g). This was purified by ACC (0- 30% EtOAc in petroleum ether (40-60 °C)) to afford white crystalline solid (2.33 g, 8.43 mmol, 57%) Rf 0.60 (7:3 Petroleum ether (40-60 °C):EtOAc (v/v)); δ_H_ (400 MHz, CDCl_3_): 8.99 (1H, s, thiadiazole 5-H), 7.27 (2H, d, *J* = 8.5 Hz, benzyl 3-H), 7.13 (1H, t, *J* = 8.5 H, benzyl 4-H), 4.89 (2H, s, benzyl CH_2_); δ_C_ (100 MHz, CDCl_3_): 164.7 (thiadiazole 2-C), 152.1 (thiadiazole 5-C), 136.3 (benzyl 2-C), 131.7 (benzyl 1-C), 129.8 (benzyl 4-C), 128.5 (benzyl 3- C), 34.73 (benzyl CH_2_); m/z ES+ Found MNa^+^ 298.9235, C_9_H_6_Cl_2_N_2_S_2_ requires MNa^+^ 298.9242

2-Bromo-5-((2,6-dichlorobenzyl)thio)-1,3,4-thiadiazole (**7**)

2-((2,6-dichlorobenzyl)thio)-1,3,4-thiadiazole (**6**) (2.33 g, 8.43 mmol), & *N*-bromosuccinimide (2.10 g, 11.80 mmol) were dissolved in DCM (10 mL) and refluxed for 48 h. The reaction was then cooled, quenched with sat. aq. Na_2_S_2_O_3_ (20 mL), partitioned and the aqueous extracted with DCM (2 × 15 mL). The organic layers were combined, dried over MgSO_4_, filtered and concentrated *in vacuo* to give an orange, oily crystals (3.15 g). This was purified by ACC (0- 30% EtOAc in Petroleum ether (40-60 °C)) to afford a white crystalline solid (2.80 g, 7.87 mmol, 78 %) Rf 0.80 (7:3 Petroleum ether (40-60 °C):EtOAc (v/v)); δ_H_ (400 MHz, CDCl_3_): 7.37 (2H, d, *J* = 8 Hz, benzyl 3-H), 7.24 (1H, dd, *J* = 8.5 & 7.5 Hz, benzyl 4-H), 4.92 (2H, s benzyl CH_2_); δ_C_ (100 MHz, CDCl_3_): 168.1 (thiadiazole 5-C), 138.1 (thiadiazole 2-C), 136.3 (benzyl 2- C), 131.5 (benzyl 1-C), 129.9 (benzyl 4-C), 128.6 (benzyl 3-C), 34.6 (benzyl CH_2_); m/z ES+ Found MH^+^ 356.8462, C_9_H_5_BrCl_2_N_4_S_2_ requires MH^+^ 356.8512

3-(5-((2,6-dichlorobenzyl)thio)-1,3,4-thiadiazol-2-yl)aniline **KC156**

General procedure A was followed using 2-bromo-5-((2,6-dichlorobenzyl)thio)-1,3,4- thiadiazole (**7**) (100 mg, 0.28 mmol), 3-aminophenyl boronic acid (40 mg, 0.28 mmol), K_2_CO_3_ (155 mg, 1.12 mmol), Pd(PPh_3_)_4_ (35 mg, 0.03 mmol), 1,4-dioxane (2 mL) and water (2 mL) to afford a crude brown oil (165 mg). This was purified by ACC (0- 60% EtOAc in Petroleum ether (40-60 °C)) followed by HPLC (50-95% MeCN in H_2_O with a 0.1% formic acid additive) to afford a white solid (26 mg, 0.70 mmol, 25%). Rf 0.3 (7:3 Petroleum ether (40-60 °C):EtOAc (v/v)); δ_H_ (400 MHz, D_6_-Acetone): 7.37 (2H, d, *J* = 7.5 Hz, benzyl 3-H), 7.27 (1H, dd, *J* = 9.0 & 7.0 Hz, benzyl 4-H), 7.16 (1H, ap. t, *J* =2 Hz, anilinyl 2- H), 7.07 (1H, ap. t, *J* = 8.0 Hz, anilinyl 5-H), 7.00 (1H, ddd, *J* = 8.0, 2.0 & 1.0 Hz, 6-H), 4.79 (2H, s, benzyl CH_2_); δ_C_ (100 MHz, D_6_-Acetone): 170.0 (thiadiazolyl 2-C), 162.5 (thiadiazolyl 5- C), 149.4 (aniline 1-C), 135.8 (benzyl 2-C), 132.1 (benzyl 1-C), 130.5 (benzyl 4-C), 130.0 (aniline 5-C), 128.8 (benzyl 3-C), 118.2 (aniline 3-C), 117.1 (aniline 4-C), 116.0 (aniline 6-C), 112.5 (aniline 2-C), 34.5 (benzyl CH_2_); m/z ES+ Found MH^+^ 367.9841, C_15_H_11_Cl_2_N_3_S_s_ requires MH^+^ 367.9844

4-(5-((2,6-dichlorobenzyl)thio)-1,3,4-thiadiazol-2-yl)aniline **KC146**

General procedure A was followed using 2-bromo-5-((2,6-dichlorobenzyl)thio)- 1,3,4- thiadiazole (**7**) (100 mg, 0.28 mmol), 4-aminophenyl boronic acid (39 mg, 0.28 mmol), K_2_CO_3_ (155 mg, 1.12 mmol), Pd(PPh_3_)_4_ (35 mg, 0.03 mmol), 1,4-dioxane (2 mL) and H_2_O (2 mL) to afford a crude brown oil (146 mg). This was purified by ACC (20-70% EtOAc in Petroleum ether (40-60 °C)) to afford a yellow solid (22 mg, 0.06 mmol, 21%). Rf 0.30 (7:3 Petroleum ether (40-60 °C):EtOAc (v/v)); δ_H_ (400 MHz, CDCl_3_): 7.63 (1H, d, *J* = 8.5 Hz, phenyl 2-H), 7.26 (2H, d, *J* = 8 Hz, benzyl 3-H), 7.12 (1H, dd, *J* = 8.5 & 7.5 Hz, benzyl 4-H), 6.64 (d, *J* = 8.5 Hz, phenyl 3-H), 4.84 (2H, s, benzyl CH_2_); δ_C_ (100 MHz, CDCl_3_): 169.8 (thiadiazole 2-C), 167.8 (thiadiazole 5-C), 149.3 (phenyl 4-C), 136.3 (benzyl 2-C), 132.2 (benzyl 2-C), 129.6 (benzyl 4-C), 129.4 (phenyl 2-C), 128.5 (benzyl 3-C), 120.1 (phenyl 4-C), 114.9 (phenyl 3-C), 34.8 (benzyl CH_2_); m/z ES+ Found MH^+^ 367.9893, C_15_H_11_Cl_2_N_3_S_2_ requires MH^+^ 367.9850

3-(5-((2,6-Dichlorobenzyl)thio)-1,3,4-thiadiazol-2-yl)benzonitrile **KC183**

General procedure A was followed using 2-bromo-5-((2,6-dichlorobenzyl)thio)- 1,3,4- thiadiazole (**7**) (100 mg, 0.28 mmol), (3-cyanophenyl)boronic acid (37 mg, 0.28 mmol), K_2_CO_3_ (155 mg, 1.12 mmol), Pd(PPh_3_)_4_ (35 mg, 0.03 mmol), 1,4-dioxane (2 mL) and H_2_O (2 mL) to afford a crude brown solid (390 mg). This was purified by ACC (0-60% ethyl acetate in petroleum ether (40-60 °C)) to afford a yellow solid (25 mg, 0.05 mmol, 18%). δ_H_ (400 MHz, CDCl_3_): 8.12 (1H, s, benzonitrilyl 2-H), 8.07 (1H, d, *J* = 8.0 Hz, benzonitrilyl 4-H), 7.70 (1H, d, *J* = 8.0 Hz, benzonitrilyl 6-H), 7.55 (1H, ap. *t*, J = 8.0 Hz, benzonitrilyl 5-H), 7.29 (2H, d, *J* = 8.0 Hz, benzyl 3-H), 7.15 (1H, t, *J* = 8.0 Hz, benzyl 4-H), 4.92 (2H, s, benzyl CH_2_); δ_C_ (100 MHz, CDCl_3_): 166.4 (thiadiazolyl 2/5-C), 165.6 (thiadiazolyl 2/5-C), 136.3 (benzyl 2-C), 134.1 (benzonitrilyl 6-C), 131.6 (benzonitrilyl 4-C), 131.6 (benzyl 1-C), 131.2 (benzonitrilyl 1-C), 131.1 (benzonitrilyl 2-C), 130.2 (benzonitrilyl 5-C), 129.9 (benzyl 4-C), 128.6 (benzyl 3-C), 117.7 (nitrile), 113.8 (benzonitrilyl 3-C), 34.7 (benzyl CH_2_); m/z ES+ Found MH^+^ 377.9685, C_16_H_9_Cl_2_N_3_S_2_ requires MH^+^ 377.9688

Ethyl 2-([(2,6-dichlorophenyl)methyl]sulfanyl)-1,3,4-thiadiazole-5-carboxylate (**8**)

To a solution of 2,6-dichlorobenzyl thiol (**5**) (502 mg, 2.60 mmol) and ethyl 5-bromo-1,3,4- thiadiazole-2-carboxylate (570 mg, 2.41 mmol) in an. DMF (8 mL) under N_2_ was added Et_3_N (0.40 mL, 2.89 mmol) and the reaction heated to 90 °C for 21h. The reaction was then cooled to RT and diluted with H_2_O (100 mL). The aqueous solution was extracted with EtOAc (2 x 70 mL) and then combined organic phases washed with brine (70 mL) and 10% LiCl solution (70 mL), dried (Na_2_SO_4_) and concentrated *in vacuo*. ACC (0-20% EtOAc in petroleum ether (40- 60 °C)) to give the title compound as a white solid (670 mg, 1.92 mmol, 80%). Rf 0.66 (8:2 Petroleum ether (40-60 °C):EtOAc (v/v));δ_H_ (400 MHz, CDCl_3_): 7.28 (2H, d, *J* = 8.1 Hz, benzyl H-3/5), 7.17-7.13 (1H, m, benzyl H-4), 4.94 (2H, s, benzylic CH_2_), 4.43 (2H, q, *J* = 7.1 Hz, C*H*_2_CH_3_), 1.38 (3H, t, *J* = 7.12 Hz, CH_2_C*H*_3_); δ_C_ (100 MHz, CDCl_3_): 170.4 (thiadiazole C-2), 160.2 (C=O), 158.5 (thiadiazole C-5), 136.3 (benzyl C-2/6), 131.3 (benzyl C-1), 130.0 (benzyl C-4), 128.6 (benzyl C-3/5), 63.3 (*C*H_2_CH_3_), 34.5 (benzylic CH_2_), 14.2 (CH_2_*C*H_3_); m/z ES+Found MH^+^ 348.9631, C_12_H_11_Cl_2_N_2_O_2_S_2_ requires MH^+^ 348.9633 (2-([(2,6-Dichlorophenyl)methyl]sulfanyl)-1,3,4-thiadiazol-5-yl)(piperidin-1-yl)methanone

### CHR-1741-077

To a solution of ethyl 2-([(2,6-dichlorophenyl)methyl]sulfanyl)-1,3,4-thiadiazole-5-carboxylate (**8**) (55 mg, 0.16 mmol) in EtOH (4 mL) under N_2_ was added piperidine (0.08 mL, 0.79 mmol) and the reaction heated to reflux. After 3h the reaction was cooled to RT and volatile solvents removed *in vacuo*. The crude mixture was dissolved in H_2_O (30 mL) and extracted with EtOAc (3 x 40 mL), combined organic phases were washed with brine (30 mL), dried (Na_2_SO_4_) and concentrated *in vacuo*. ACC (0-30% EtOAc in petroleum ether (40-60 °C)) to give the title compound as a white solid (24 mg, 0.062 mmol, 39%). Rf 0.48 (8:2 Petroleum ether (40-60 °C):EtOAc (v/v)); δ_H_ (400 MHz, CDCl_3_): 7.28 (2H, d, *J* = 8.04 Hz, benzyl H-3/5), 7.17-7.13 (1H, m, benzyl H-4), 4.90 (2H, s, benzylic CH_2_), 4.15-4-13 (2H, m, piperidine), 3.67-3.65 (2H, m, piperidine), 1.70-1.65 (6H, m, piperidine); δ_C_ (100 MHz, CDCl_3_): 169.0 (thiadiazole C-2), 167.0 (thiadiazole C-5), 156.8 (C=O), 136.3 (benzyl C-2/6), 131.3 (benzyl C-1), 129.9 (benzyl C-4), 128.6 (benzyl C-3/5), 47.8 (piperidine *C*HN), 44.8 (piperidine *C*HN), 34.2 (benzylic CH_2_), 26.7 (piperidine), 25.8 (piperidine), 24.5 (piperidine); m/z ES+ Found MNa^+^ 409.9930, C_15_H_15_Cl_2_N_3_NaOS_2_ requires MNa^+^ 409.9925

2-([(2,6-Dichlorophenyl)methyl]sulfanyl)-1,3,4-thiadiazol-5-amine (**9**)

To a solution of 2-amino-5-bromo-1,3,4-thiadiazole (557 mg, 3.09 mmol) and 2,6- dichlorobenzyl thiol (**5**) (717 mg, 3.71 mmol) in an. DMF (15 mL) under N_2_ was added Et_3_N (0.52 mmol, 3.71 mmol) and the reaction heated to 90 °C. After heating for 20h reaction was cooled to RT and diluted with H_2_O (100 mL). The resulting precipitate was collected by filtration. ACC (30-70% EtOAc in petroleum ether (40-60 °C)) to give the title compound as a white solid (665 mg, 2.28 mmol, 74%). Rf 0.28 (1:1 Petroleum ether (40-60 °C):EtOAc (v/v)); δ_H_ (400 MHz, d_6_-DMSO): 7.51 (2H, d, *J* = 7.8 Hz, benzyl H-3/5), 7.42-7.35 (3H, m, benzyl H-4 and NH_2_), 4.46 (2H, s, benzylic CH_2_); δ_C_ (100 MHz, d_6_-DMSO): 171.8 (thiadiazole C-5), 147.6 (thiadiazole C-5), 135.4 (benzyl C-2/6), 133.2 (benzyl C-1), 130.8 (benzyl C-4), 129.2 (benzyl C-3/5), 36.0 (benzylic CH_2_); m/z ES+ Found MH^+^ 291.9529, C_9_H_8_Cl_2_N_3_S_2_ requires MH^+^ 291.9531 *N*-(5-([(2,6-Dichlorophenyl)methyl]sulfanyl)-1,3,4-thiadiazol-2-yl)-1,2-oxazole-5-carboxamide

### CHR-1871-005

To a suspension of 2-([(2,6-dichlorophenyl)methyl]sulfanyl)-1,3,4-thiadiazol-5-amine (**9**) (59 mg, 0.20 mmol) in anhydrous THF (4 mL) under N_2_ was added DMAP (1 mg, 0.01 mmol) Et_3_N (0.03 mL, 0.22 mmol) followed by isoxazole-5-carbonyl chloride (0.03 mL, 0.30 mmol). The reaction was stirred at room temperature for 20 h, then diluted with water (30 mL). The aqueous solution was extracted with EtOAc (3 x 20 mL), the combined organic phases were washed with brine (20 mL), dried (Na_2_SO_4_) and concentrated *in vacuo*. ACC (50-100% EtOAc in Petroleum ether (40-60 °C)) to afford a white residue (52 mg, 0.13 mmol, 67 %). Rf 0.56 (3:7 Petroleum ether (40 - 60 °C):EtOAc (v/v)); δ_H_ (400 MHz, d_6_-DMSO): 13.87 (1H, br. s, NH), 8.88 (1H, s, isoxazole H-3), 7.55-7.53 (3H, m, isoxazole H-4 and benzyl H-3/5), 7.43-7.39 (1H, m, benzyl H-4), 4.70 (2H, s, benzylic CH_2_); δ_C_ (125 MHz, d_6_-DMSO): 160.9 (thiadiazole C-5), 157.7 (thiadiazole C-2 or isoxazole C-5), 155.2 (C=O), 152.5 (isoxazole C-3). 135.6 (benzyl C-2/6), 132.5 (benzyl C-1), 131.1 (benzyl C-4), 129.3 (benzyl C-3/5), 108.8 (isoxazole C-4), 35.1 (benzylic CH_2_); m/z ES+ Found MH^+^ 386. 9534, C_13_H_8_Cl_2_N_4_O_2_S_2_ requires 386.9538 MH^+^

